# Machine Learning for Real-Time Aggregated Prediction of Hospital Admission for Emergency Patients

**DOI:** 10.1101/2022.03.07.22271999

**Authors:** Zella King, Joseph Farrington, Martin Utley, Enoch Kung, Samer Elkhodair, Steve Harris, Richard Sekula, Jonathan Gillham, Kezhi Li, Sonya Crowe

## Abstract

Machine learning for hospital operations is under-studied. We present a prediction pipeline that uses live electronic health-records for patients in a UK teaching hospital’s emergency department (ED) to generate short-term, probabilistic forecasts of emergency admissions. A set of XGBoost classifiers applied to 109,465 ED visits yielded AUROCs from 0.82 to 0.90 depending on elapsed visit-time at the point of prediction. Patient-level probabilities of admission were aggregated to forecast the number of admissions among current ED patients and, incorporating patients yet to arrive, total emergency admissions within specified time-windows. The pipeline gave a mean absolute error (MAE) of 4.0 admissions (mean percentage error of 17%) versus 6.5 (32%) for a benchmark metric. Models developed with 104,504 later visits during the Covid-19 pandemic gave AUROCs of 0.68-0.90 and MAE of 4.2 (30%) versus a 4.9 (33%) benchmark. We discuss how we surmounted challenges of designing and implementing models for real-time use, including temporal framing, data preparation, and changing operational conditions.

## Introduction

To date, most applications of Artificial Intelligence (AI) to healthcare have been applied to address clinical questions at the level of individual patients^1^. Now that many hospitals have electronic health records (EHRs) and data warehouse capabilities, there is the potential to exploit the promise of AI for operational purposes^2^. Hospitals are highly connected systems in which capacity constraints in one area (for example, lack of ward beds) impede the flow of patients from other locations, such as the emergency department (ED)^3^ or those ready for discharge from intensive care^4^. Arrivals to the ED show diurnal and seasonal variations, with predicted peaks in the morning and early evening, but workflows elsewhere in a hospital mean that discharges from the hospital happen late in the day, creating flow problems^5^. This mismatch of cadence between different parts of the hospital results in patients boarding in ED, or being admitted to inappropriate wards, with adverse consequences including longer stays^6^, greater risk of medical errors^7^ and worse long-term outcomes in elderly patients^8^.

Hospital services can be managed more efficiently if accurate short-term forecasts for emergency demand are available^9,10^. Currently, most hospitals use simple heuristics to make short-term forecasts of numbers of emergency admissions, which are based on rolling averages for each day of the week^11^. Scholars have suggested improvements using Bayesian approaches or auto-regressive inductive moving averages with meteorological, public health and geographic data^9,12,13^. However, such methods do not take account of stochastic nature of ED arrivals^14^ and cannot be adapted to reflect the case mix of people in the ED at a given point in time. In hospitals with EHRs, where staff are recording patient data at the point of care, there is an opportunity to use EHR data to generate short-horizon predictions of bed demand. These would help the teams responsible for allocating beds make best use of available capacity and reduce cancellations of elective admissions.

ML is attractive for such predictions because its aggregation of weak predictors may create a strong prediction model^2^. Emergency medicine scholars have compared predictions made by ML algorithms against conventional approaches like linear regression and naïve Bayes^10,15^. It is common for such studies to use arrival characteristics (e.g. arrival by ambulance or on foot), triage data and prior visit history^16–18^ to make predictions, although recent studies have included a wider variety of data captured by EHRs, including medical history, presenting condition and pathology data^10,19–21^. Hong et al^10^ showed that ML algorithms like gradient-boosted trees and deep neural networks, applied to a large EHR dataset of 972 variables, improved predictive performance. By including data on lab test results and procedures, El-Bouri et al^21^ were able to predict which medical specialty patients would be admitted to. Barak-Corren et al’s study^19^ is one of few in emergency medicine to address the challenges of making predictions during a patient’s visit to ED. They built progressive datasets from historical data, each intended to reflect the data usually available at 10, 60 and 120 minutes after presentation to the ED. Notwithstanding their use of chief complaint data that was entered by ED receptionists as free text and retrospectively coded by the researchers, they were able to show that the later datasets offered better predictions than at 10 minutes. Their study demonstrates the potential that EHRs offer for improving on approaches that use triage data only.

Although these studies demonstrate the predictive utility of ML, they do not unlock its potential to generate predictions in real-time to help managers address problems of patient flow. Building a model for implementation involves several additional challenges to those encountered when simply optimising the technical performance of a prediction model. These include preparing training examples of incomplete visits from historic data in which visits have been completed^22^, making decisions about the temporal framing of the model (for example, at what point in the visit to check if the outcome of interest has occurred)^23^, and planning for a drift in model performance over time^24^. Models for real-time prediction have been trained in clinical contexts such as circulatory failure in critical care^25^ and post-operative complications^26,27^. These are contexts where patient observations are taken with high frequency whereas the frequency of data collection and the type of data collected varies greatly from patient to patient in the ED. A patient in the resuscitation area of an ED may have frequent observations, while a patient in the waiting room has no data collected. These heterogeneous data profiles are themselves indicative of likelihood of admission.

From the bed planners’ point of view, knowing the probability that a particular patient will be admitted is less valuable than knowing in aggregate how many patients to plan for. In this respect a prediction tool that can provide a probability distribution for the number of admissions in a given time frame is more useful than one that solely estimates probability of admission at the patient level. One study in emergency medicine derived an expected number of admissions among a roomful of patients in ED by summing their individual probabilities of admission^28^, but there was no presentation of the uncertainty of their point estimates. Also, when making predictions for admissions within a time-window after the prediction is made, projections must allow for the number of patients not on the ED at the prediction time who will arrive and be admitted within the window^29^.

If models are to be used operationally, their performance needs to be sustained over time as care provision, patient characteristics and the systems used to capture data evolve^24^. Real-time operational models also need to cover the ‘last mile’ of AI deployment; this means that the applications that generate predictions can run end-to-end without human intervention. This last mile is the most neglected^30^, leading to calls for a delivery science for AI, in which AI is viewed as an enabling component within an operational workflow, rather than an end in itself^31^.

This research aimed to harness the heterogenous stream of real-time data coming from patients in the ED of a UK hospital to make predictions of aggregate admissions in a short time horizon. Bed planners at the hospital were closely involved with the research team to specify their requirements. They requested predictions for bed requirements in the next four and eight hours to be sent at four times daily, to coincide with their own capacity reporting. As part of the project, we developed an application that formats and sends an email to the bed planners at the four report times. See Supplementary Note 9 for details of the bed planners’ workflow and the application we created. In this paper, we explain how the predictions are generated, evaluate their performance and compare them with standard benchmarks.

The contributions of the research are: the development and deployment of a ML-based information product in use in hospital operations; the demonstration of a method to train ML models for real-time use when patient-level data is variable between patients and over the course of individual visits; the incorporation of a method to aggregate individual-level predictions for operational planning purposes; and an exposition of some of the challenges associated with developing models for real-time implementation.

## Results

Figure 1 illustrates a real example of predictions generated at 16:00 on 11 May 2021 using the seven-step pipeline built through this work. As noted above, the bed planners wanted these predictions at four times daily (06:00, 12:00, 16:00 and 22:00). The following paragraphs present an evaluation of the predictions made at the four prediction times on a test set of 97 days from 13 December 2019 to 18 March 2020.

**Figure 1:**
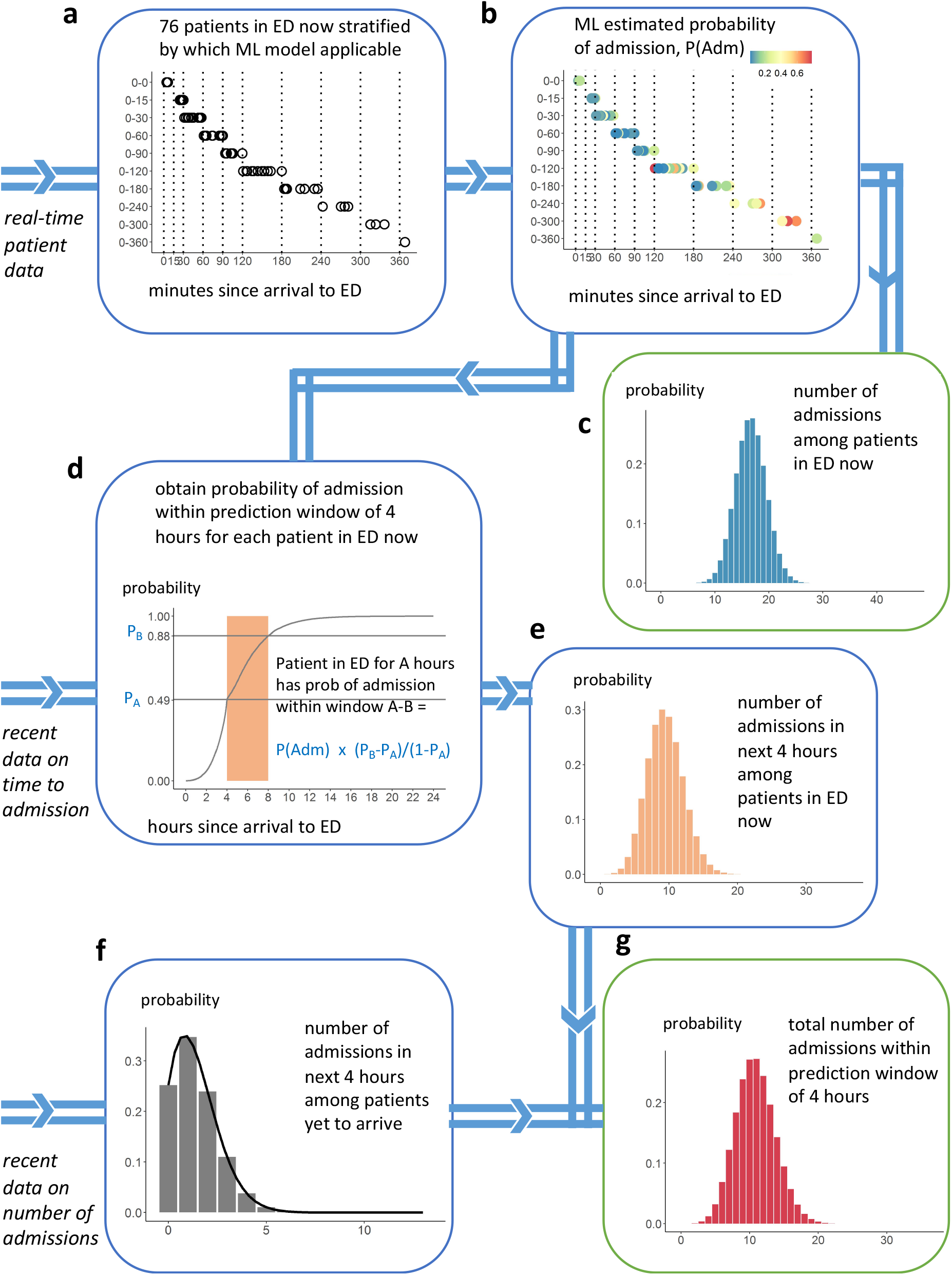
Example of the seven-step prediction pipeline using a real example, predicting the number of admissions within 4 hours after 16:00 on 11 May 2021. **a** illustrates the roomful of patients in the ED at the prediction time on the day of interest, grouped according to how long they had been in the ED since arrival. **b** shows each patient’s probability of admission, generated using a set of ML models. These are combined in **c** into a probability distribution for the number of admissions among this roomful of patients. **d** shows the probability of admission within 4 hours calculated from recent data on time to admission, taking into account the time the patient has been in ED up to the prediction time **e** shows a distribution over the number of admissions among the roomful of patients in the prediction window of 4 hours. **f** shows a probability distribution over the number of patients who have not yet arrived, who will be admitted in the prediction window, generated by a Poisson equation. **g** shows the final probability distribution for the number of admissions within the prediction window

At each prediction time, EHR data on the set of patients in ED was retrieved (Step 1). A ML prediction was made for each about their probability of admission at Step 2. At Step 3, the individual probabilities were combined to give a probability distribution for the number of admissions from the patients currently in ED. At Step 4, the individual probability of admission for each patient was combined with survival analysis to give for each patient the probability that they would be admitted within the prediction window, accounting for when they arrived and the number of patients in ED when they arrived. At Step 5 the individual probabilities from Step 4 were combined to give a probability distribution for the number of admissions within the prediction window from patients currently in the ED. At Step 6 Poisson regression was used to give a probability distribution for the number of additional patients that would arrive and be admitted within the prediction window. Finally, at Step 7, the distributions obtained at Steps 5 and 6 were convoluted to give a probability distribution for the total number of admissions within the prediction window by patients currently in the ED and others yet to arrive.

### Results for steps 1-2: Machine learning to estimate individual probability of admission

The most important features for admission prediction selected by the XGBoost classifier are shown in Figure *2*a for 12 distinct models developed for use with patients that have been on the ED for increasing periods of time, where model T15 is based on data available within the first 15 minutes of a visit and T240 based on data available within the first 240 minutes etc. See Supplementary Table S2 for a glossary of features.

**Figure 2:**
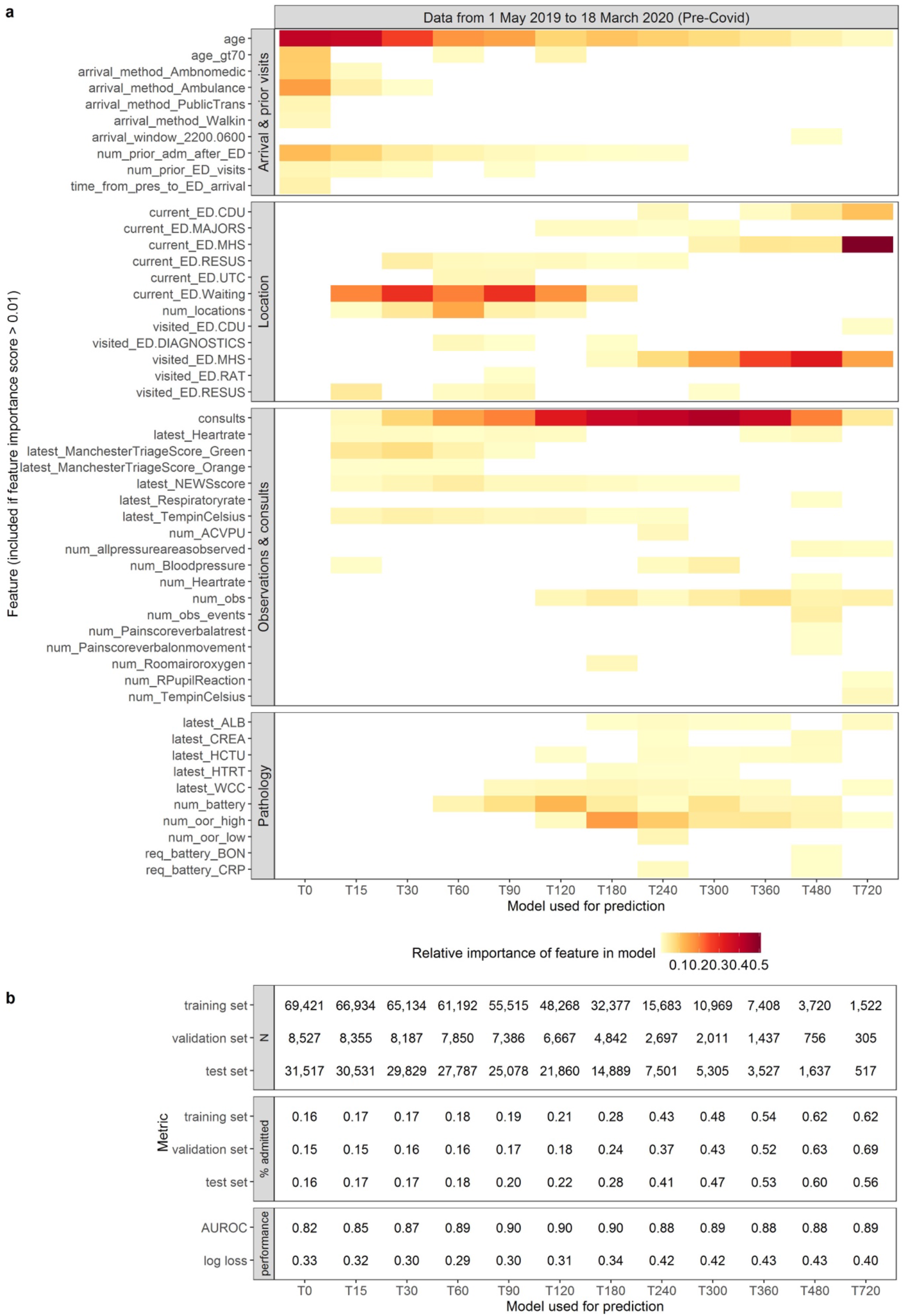
Feature importance and performance for each model on test set. **a** shows the feature importances, presented for ease of interpretation in four groups (visit data, location history, observations and consults, and pathology). The colour intensity reflects the relative importance of each feature within each model. For simplicity of presentation, a feature is excluded from the figure if it had a raw importance of less than 0.01 in all models. **b** shows the number of visits, admission proportion and performance of each model. See Supplementary Table S2 for a glossary of features and Supplementary Note 6 for equivalent analysis of later visits during the Covid-19 pandemic

Among the **visit features**, age, arrival method and previous admission are initially important but these diminish as elapsed time passes and signals from other features become stronger. Among the **location features**, being in a waiting area, or being in the resuscitation area (Resus), are important between 15 and 120 minutes. (See Supplementary Note 1 for more information on the locations within the ED). After 180 minutes, having visited or being in the Mental Health Stream (MHS) is important; this location is used for people with mental health disorders who are usually discharged to a specialist facility but who often stay in MHS for a long time. That explains why MHS is so important in the T720 model, whose training examples include a higher proportion of MHS visits. After 240 min, being in the Clinical Decision Unit (CDU) becomes important; this location is for people waiting for test results or being observed, prior to discharge.

Among the **observation and consults features**, the number of consults with inpatient specialists, signalling likely admission, is important in all models, especially between 180 and 360 minutes. Triage scores are important up to 60 minutes, and the National Early Warning Score (NEWS) remains important up to 240 minutes. The cumulative number of measurements taken, and the number of times certain indicators are recorded, like nurse checks of body pressure areas, are more important later in the ED stay, presumably reflecting sustained attention by staff to more unwell patients. Among the **pathology features**, test result values and the number of out-of-range results become important from 60 min onwards, as lab tests results start to be returned to the ED. Requests for certain sets of lab tests (bone profile and C-reactive protein) are important for longer-staying patients.

The performance metrics for each model are shown in Figure *2*b. The models achieved lowest log loss when presented with patients with elapsed times of between 30 and 120 minutes, and the best Area Under the Receiving Operating Curve (AUROC) of 0.90 between 90 and 180 minutes. Up to this point, few patients have departed, so the models can differentiate well between likely admissions and discharges. As time goes on, the more straightforward discharges and admissions are made, the number of training examples diminishes (see Figure 2b) and the case mix includes a higher proportion of more clinically complex cases which are harder to predict.

Calibration plots for each ML model are shown in Figure 3, applied to all visits in the test set. All models are well calibrated, up to the final two models which related to a very small subset of visits where patients remain in the ED after 8 hours.

**Figure 3:**
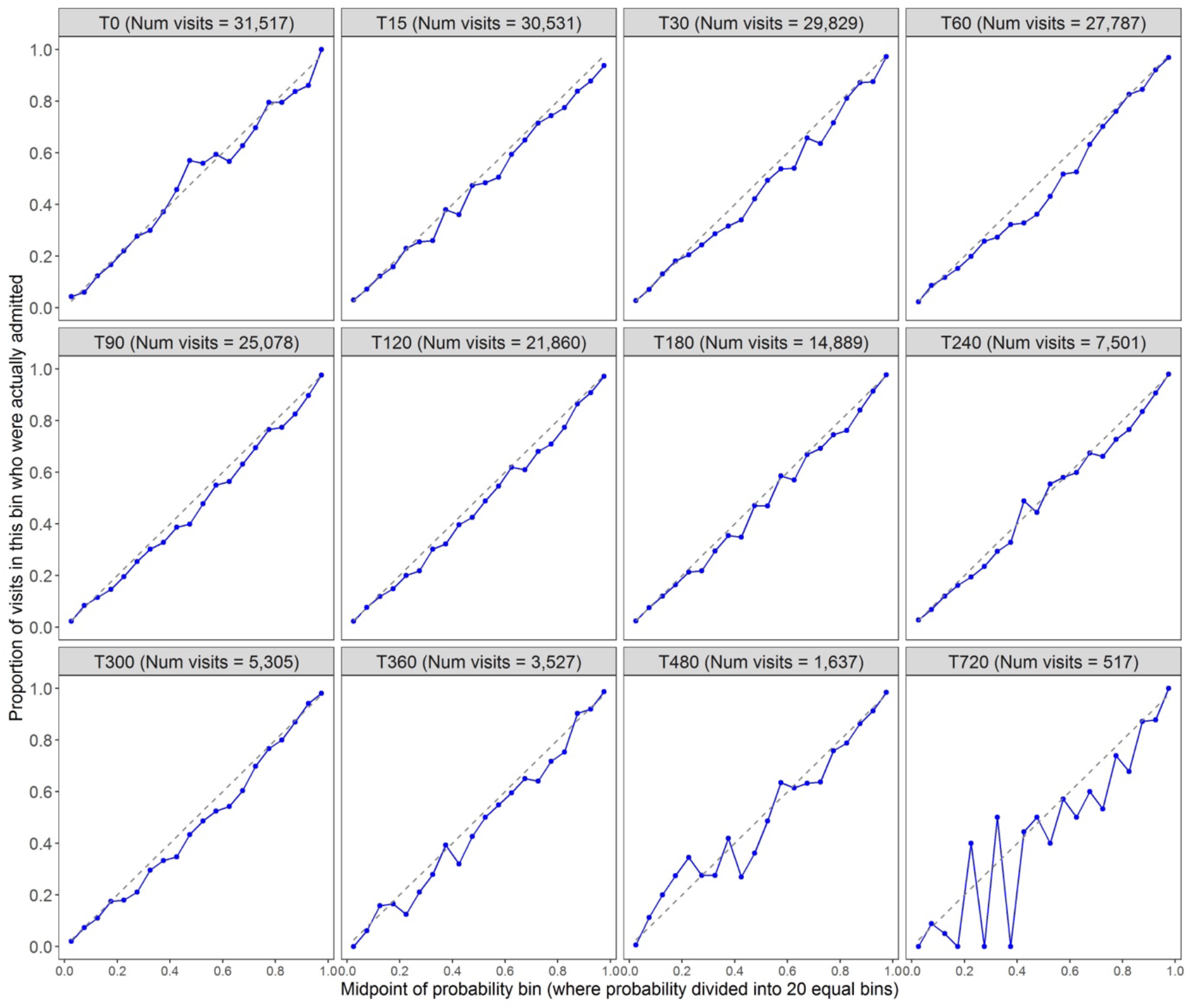
Calibration plots for each of the ML models applied to the test set.

In additional analyses we found XGBoost to outperform Random Forest (RF) in terms of log loss on all 12 models (Figure 4). Logistic Regression with Lasso regularisation gave performance almost as good as XGBoost, with increased regularisation (and as a consequence sparser regression models) giving less good performance (Figure 4). Both Random Forrest and Logistic Regression models drew on similar feature sets to XGBoost (Supplementary Note 8).

**Figure 4:**
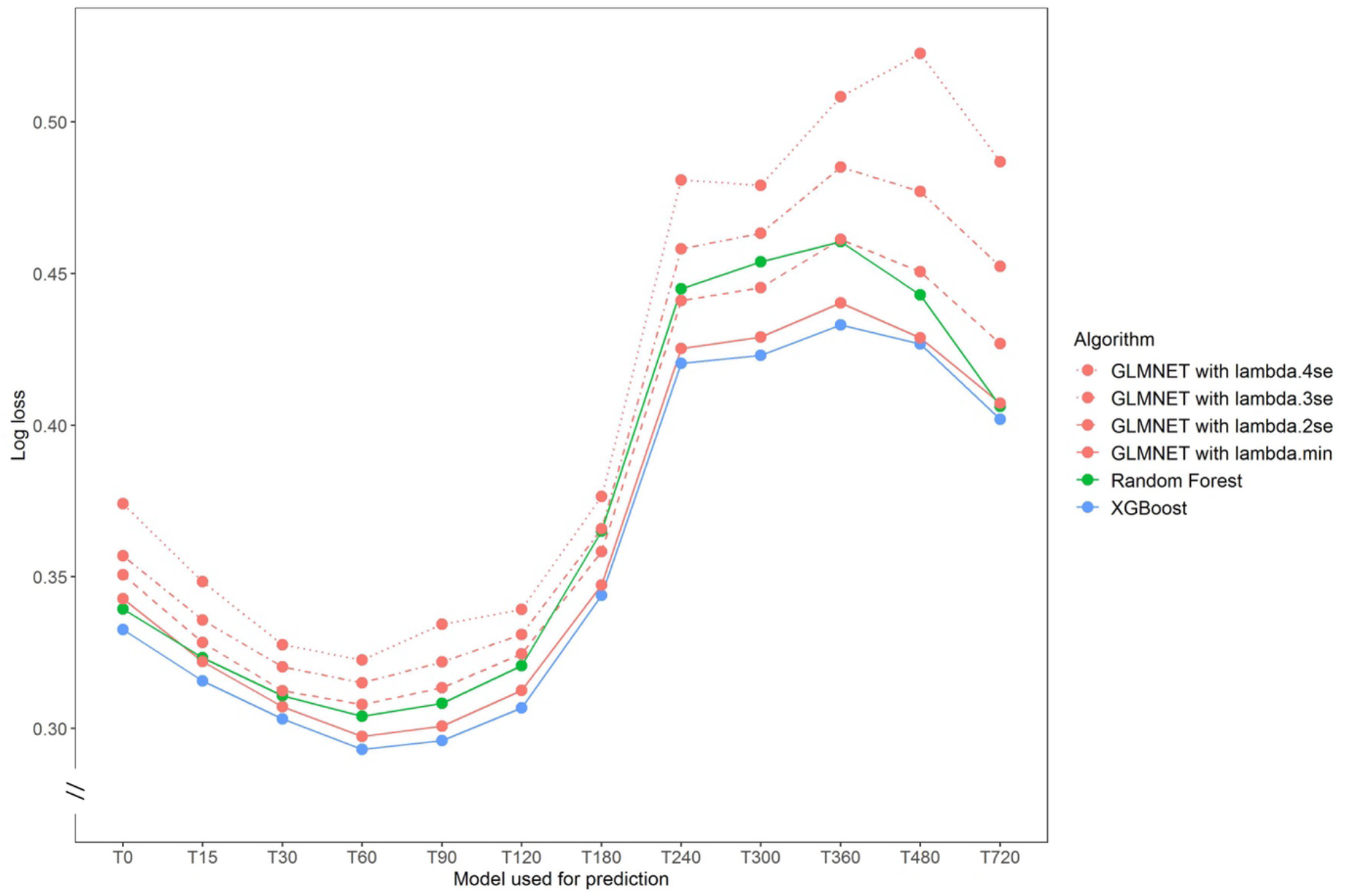
Comparison of model performance of XGBoost, Random Forest and Logistic Regression on test set. For each set of 12 models (T0 to T720), the log loss achieved by each algorithm is shown as a circle with a line to link circles representing the same algorithm. Logistic regression (LR) results are in red with diminishing line solidity as lambda, the penalty value, is pushed towards greater regularisation. Lamda.min is the value of lambda with minimum cross-validated error, which is the optimal model. Lambda.2se is the value that gives the most regularised model such that the cross-validated error is within 2 standard errors of the minimum. Lambda.3se and Lambda.4se are within 3 and 4 standard errors respectively.

### Results for steps 3-7: Aggregated predictions of number of emergency admissions

In the remaining steps, individual probabilities at the patient level were aggregated into probability distributions over the number of admissions at each prediction time. Figure 5 uses QQ plots to evaluate the concordance between observations and the predicted distributions created at Steps 3, 5 and 7 of the prediction pipeline (see Methods for more about the intervening steps). From visual inspection of the QQ plots, there is very good concurrence between the predicted distributions and observations after Step 3. (Concurrence at Step 3 was worse when using only 3 rather than 12 models at Step 2, as explained in Supplementary Note 7 and shown in Figure S17.) After Step 5, concurrence remains good, although (especially for an eight-hour prediction window) the predicted distributions underestimate slightly the number of admissions within the prediction window, suggesting that patients were taking less time to be admitted than predicted. Similar concurrence is observed after step 7.

**Figure 5:**
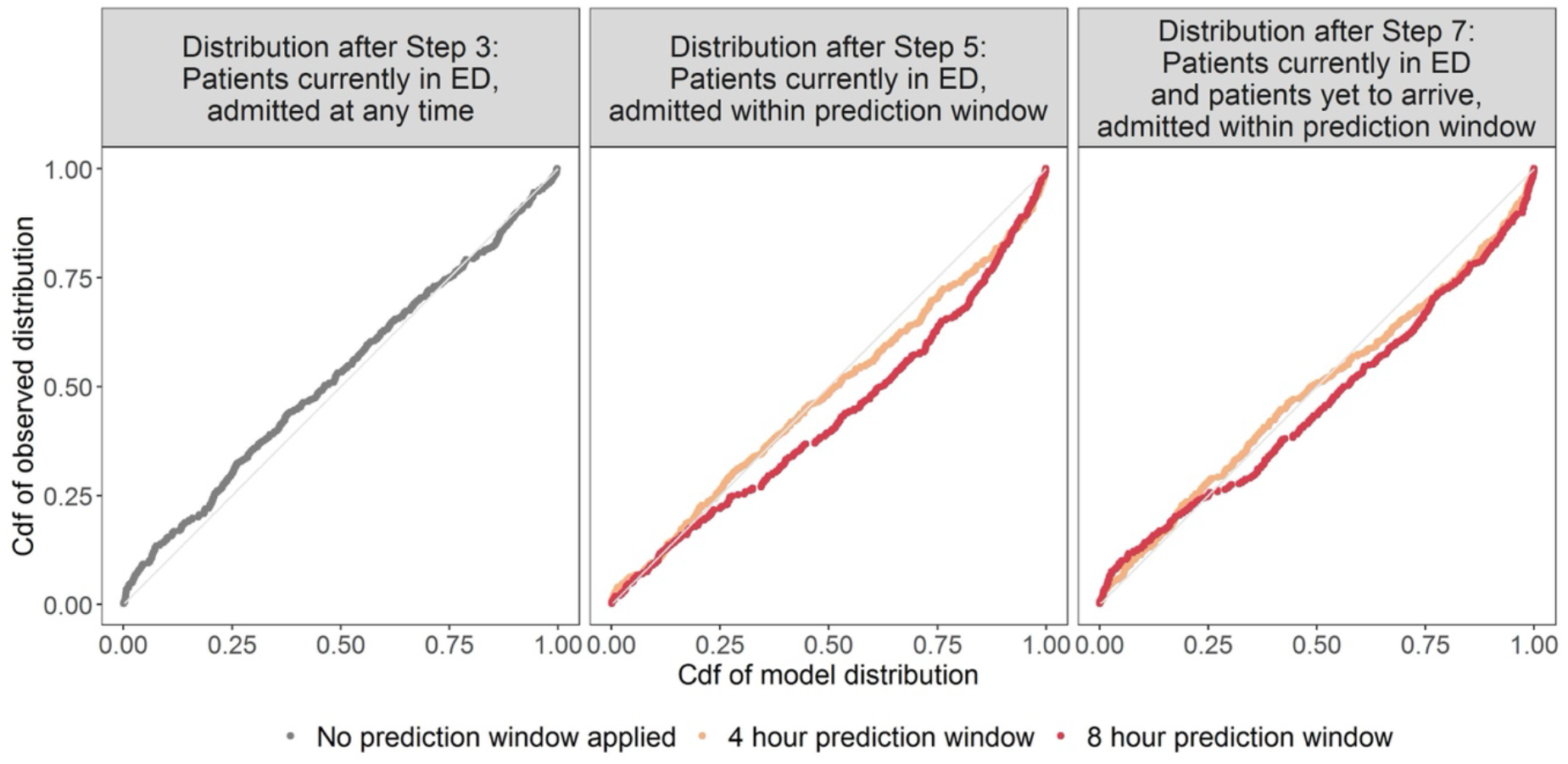
QQ plots evaluating the predicted distributions of the number of admissions after Steps 3, 5 and 7. Abbreviations: cdf (cumulative distribution function). The first column of plots evaluates the distributions for all patients currently in the ED, without applying a prediction window. The second column evaluates the distributions for all patients currently in the ED, with a prediction window of 4 and 8 hours. The third column evaluates the distributions for all patients currently in ED with a prediction window and including patients yet to arrive.

Figure 6 compares the model predictions with the conventional six week rolling-average benchmark for daily admissions, adjusted for use at 16:00 hours (see Methods for how this was derived). Mean Absolute Error (MAE) was used to compare the approaches, as this avoids positive and negative deviations cancelling each other out, and the error was also expressed as a percentage of observed admissions to derive a mean percentage error (MPE). The prediction pipeline underestimated the number of admissions within the 8-hour window but performed better than the benchmark (MAE of 4.0 admissions with MPE of 17%, compared with 6.5 and 32% for the benchmark).

**Figure 6:**
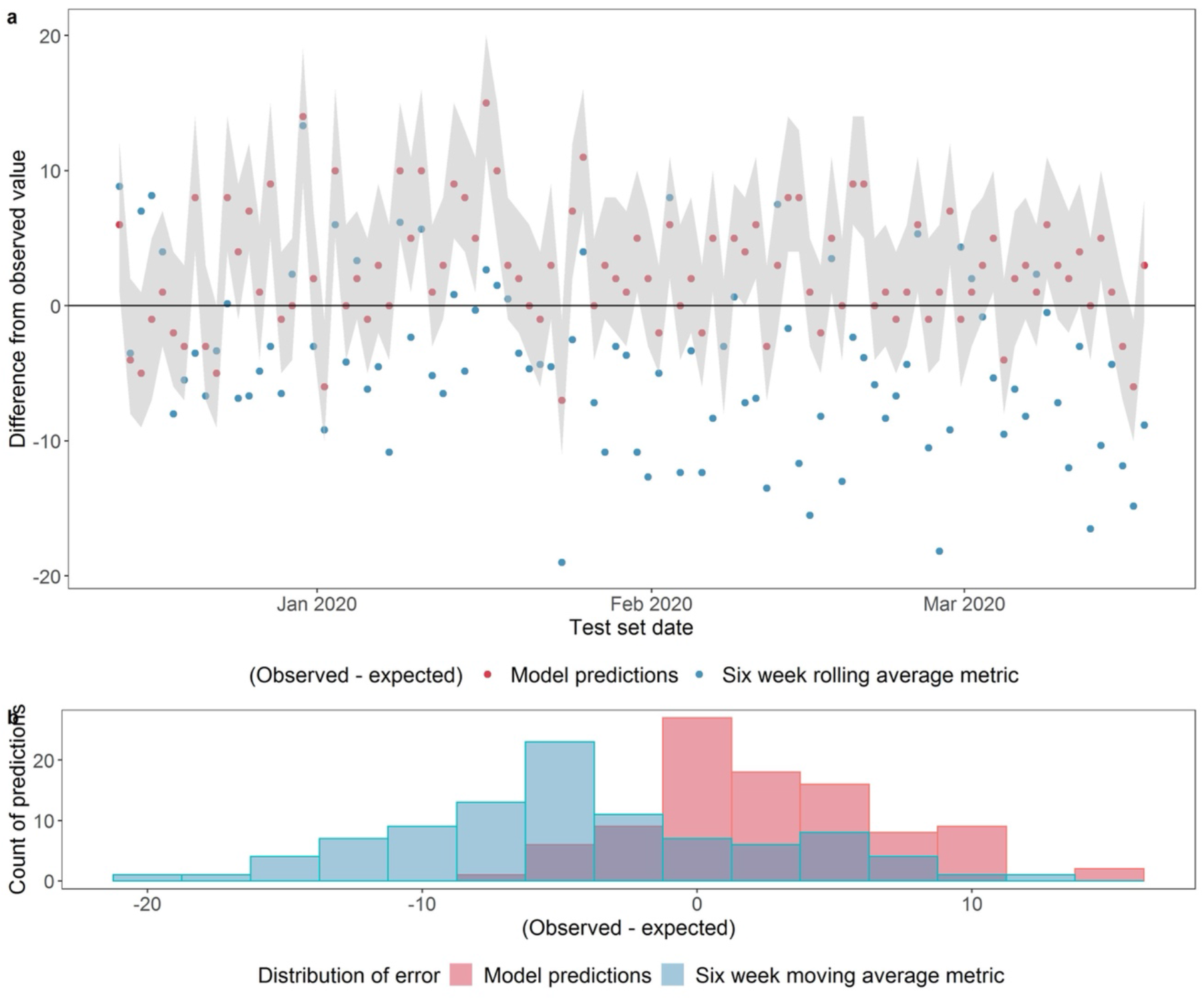
Comparing model predictions with six week rolling average benchmark for number of admissions within a prediction window of 8 hours after 16:00, including patients who are yet to arrive. **a** shows the difference between the observed number of admissions and the expected value from the probability distribution for the model predictions (the red dots) and between the observed number of admissions and expected value from the benchmark (the blue dots). Where the expected value equals the observed value, the dots fall on the x axis (y = 0). The grey shaded band represents the range of probability between the 10^th^ and the 90^th^ centile of the cumulative probability distribution of the model. **b** shows the distribution of errors (difference between observed and expected). See Methods for a rationale for conducting the evaluation at 16:00 with an 8-hour prediction window.

The results were achieved using data from May 2019 to March 2020, before the Covid-19 pandemic took hold in the UK. In Supplementary Note 6 we present the results of modelling two alternative datasets; one which starts at the point of the Covid outbreak, and one trained only on the period after SDEC was introduced. Patterns of feature importance (Supplementary Figure S11) and model performance (Supplementary Figure S12) were largely consistent across the three datasets. The seven-step pipeline needed one adjustment to accommodate the greater variation in both patients and operational conditions after the outbreak of Covid; this was to refine the survival curve used in Step 4 to draw only on rolling data from the past six weeks.

## Discussion

Our work is one of the first examples of a ML-based modelling approach that is designed and fit for the purpose of informing real-time operational management of emergency admissions. The predictions outperformed the conventional six-week rolling average benchmark. Moreover, the prediction pipeline improved on the benchmark (which only projects up to midnight) by enabling predictions for short time-horizons of 4 and 8 hours at various times throughout the day. When adapted to take account of operational variations in how long it takes a patient to be admitted during the state of flux introduced by the pandemic, the predictions made using models trained on data following the Covid outbreak were also able to improve on the benchmark. The results were achieved using only data that are available for inference in real-time.

The predictions were based on ML models that have equivalent or better performance to other studies. Using logistic regression (LR), Barak-Corren et al^19^ achieved an AUROC of 0.82 in their 10 minute model (augmented by free-text chief complaint data that was retrospectively encoded), compared with 0.85 at 15 minutes here without chief complaint; they achieved an AUROC of 0.83 after 120 minutes, compared with 0.90 here. Using XGBoost, Barak-Corren et al^33^ achieved AUROC of 0.87–0.93 with a 60 minute model, compared with 0.89 here, and Hong et al ^10^ achieved 0.87 with triage data. The challenge for predictions using ED data is the sparse and heterogenous nature of the recorded data. This study created complex models with many features, without dropping visits with missing data as other scholars have done^21^, and included real-time location features that proved to be important for the long-staying patients who are more difficult to predict for. To exploit the evolving predictive power of features, including data-completeness during patients’ visits to ED, 12 temporally framed models were used; Supplementary Note 7 shows the value added by the use of 12 models compared to 3, (the number chosen by Barak-Corren et al^33^), confirming that additional models contribute to superior performance. We found that XGBoost outperformed Random Forest and that LR optimised with Lasso regularisation was a close second. More heavily regularised LR models also performed well, suggesting that sparser models could potentially achieve acceptable performance in data-poor settings. That said, we note that that the full set of variables was available for inclusion in the regression models. In addition to good predictive performance, the XGBoost and other classifiers used here have the benefit of being more interpretable than other algorithms such as neural networks, giving some insight into how the models derive their individual-level predictions. Patterns of feature importance and model performance were largely consistent across three different datasets, confirming that the ML models are relatively robust to changes brought about by the pandemic.

This study is also, to our knowledge, one of few to aggregate patient-level probabilities into predictions of bed demand. Peck et al’s^28^ proposed summation of probabilities to calculate running bed need does not quantify uncertainty, and England et al’s^34^ simulations do not benefit from real-time data. The prediction pipeline generated output that could be evaluated at each step, which means that, when applied to data from later time periods, it is possible to identify (and potentially address), any weak step(s) in the pipeline. For example, a post-hoc change to the step that uses survival curves was made for the period following the Covid outbreak to address fluctuations in the flow of patients through the ED after the pandemic began. Other changes also required adjustments. Same Day Emergency Care (SDEC) was introduced late in 2020 and significantly changed how the ED is run. So, for the real-time information product currently in use, a training set starting in November 2020 was used to train the models, and the survival curves are updated nightly to use only time-to-admission data from the last six weeks.

The need for a post-hoc adjustment to models is reflective of wider difficulties with drift for all predictive applications that learn from past data, including but not limited to those using ML. Methods have been proposed for dealing with non-stationary learning problems^35,36^. Here a sliding window was applied to one step of the model, but changing patterns of ED presentations and/or changing operational practice may introduce drift or sudden changes in model performance, necessitating continuous monitoring. Here, the modular nature of the aggregation made it easy to swap out the bit that changed and retrain. No model of an evolving system can be expected to predict accurately in perpetuity, so some human action to monitor and revise models is acceptable and indeed expected by patients^37^. It is also important to alert users of the application if prediction quality deteriorates and we intend to do this in a future release. Nonetheless there is evidence for the robustness of the models. The ML models draw on similar features before and during Covid (see Supplementary Note 6) suggesting that the signals of likelihood of admission remain somewhat consistent.

The hospital where this work was conducted is urban with a student and commuter case mix and no major trauma centre. As ED organisation varies between hospitals (especially location and pathways) the cross-site generalisability of the findings may be restricted to similar hospitals. But aside from those based on location, the risk factors for admission found here are familiar in ED practice and therefore likely to be common to other sites^33^. There are limitations to this study. Some features used in other studies^19,21^ are not available here, particularly chief complaint and imaging. Because presentations at ED are seasonal, ideally models have more than one complete annual cycle to learn patterns from^12^; in fact the two years in this study were very different, affected by organisation changes and the impact of Covid-19 on health services. Future research in this setting could benefit from longer training periods, and focus on predictions of onward destinations, following El-Bouri^21^, to distinguish between demand for medical versus surgical admissions, and perhaps to differentiate demand according to infection status with respect to Covid-19 or other infectious diseases.

This study was novel in two respects. From a research point of view, the use of real-time EHR data is new in the literature on emergency admissions, as is the aggregation of patient-level predictions into data for operational use. In future studies, researchers may find it useful to deploy the prediction pipeline proposed here in healthcare settings with sparse and heterogeneous flows of patient data, such as outpatient clinics. Second, it presented an information product in use, co-designed with bed managers with continued, and ongoing, iterations to meet their needs. Few published studies complete the ‘last mile’ of AI deployment by reporting on models in production^38^. Here we demonstrate an application of ML in that last mile, providing an example of how ML for healthcare will need to be delivered if it is to become a dependable and reliable tool. This work draws attention to some challenges that make technically high-performing systems perform poorly for their intended use, including model drift, and considers how to address them.

In a resource-constrained environment, the benefits of ML implementations must be weighed against the costs. Our study was designed under the assumption that better information would improve bed planners’ ability to manage the complexities of patient flow, but its benefits are hard to isolate from the contribution of other workflows and initiatives in a complex and changing environment. The costs of implementation were small as the hospital had already invested in the infrastructure to utilise data from the EHR, and no data entry was required by clinical staff in real-time. Other settings without the upfront investment in infrastructure might be able to achieve similar results with fewer data points and simpler models, as suggested by our LR results, and it may be possible to implement some predictive models within an EHR. Ultimately of course, crowding in EDs is an outcome of system-wide issues downstream such as bottlenecks and capacity constraints^7,39^. AI does not do anything on its own; to succeed, it must be connected to real-world processes^40^.

## Methods

### Data source

The source of the data is HL7 messages generated by Epic, the hospital’s EHR system. These are captured as they are issued, and stored in EMAP, a PostgresSQL relational database that is kept up to date with latency of less than 5 minutes. The database records a subset of the full patient record, including observations, pathology orders and results, location of patients, consult requests, and a summary of prior visit history. Data were analysed with R version 4.0.0 using MLR3 packages^41^ to manage the ML pipeline. The real-time application is run on a security-enhanced Linux machine within the hospital network.

The study was deemed exempt from NHS Research Ethics Committee review as there is no change to treatment or services or any study randomisation of patients into different treatment groups. It was considered a Service Evaluation according to the NHS Health Research Authority decision tool (http://www.hra-decisiontools.org.uk/research/).

### Study population and datasets

The data include all inpatient and emergency visits involving an ED location from 1 May 2019 to 19 July 2021. Pre-processing steps are shown in Supplementary Figure S1. Patients under 18 on the day of admission were excluded, as were the very few who died in ED and those who self-discharged. After filtering, there were 213,985 visits. In total 36,225 (20.7%) visits ended in admission. Supplementary Note 2 explains how the outcome of admission was derived. The distribution of the outcome variable at different report times, and in periods before and during Covid, is shown in Supplementary Figure S2.

From March 2020 onward, three notable changes happened that could be material to the modelling process. First, as Covid-19 case numbers have fluctuated through various surges, EDs have had to deal with the uncertainties of a new disease and changing clinical case mix. Second, operational conditions, in terms of levels of busy-ness, have been highly variable. Numbers of presentations fell dramatically in late March 2020 (see Supplementary Figure S3) and patients were processed quickly. After a quiet period in April to June 2020, presentations gradually climbed, reaching pre-pandemic levels by mid-2021 and EDs have become progressively busier and slower to admit patients. Third, structural changes have led to various reconfigurations of care pathways, including: the introduction of Same Day Emergency Care (SDEC) in December 2020 intended to avoid unnecessary admissions^42^; the resumption of primary care and urgent pediatric services; the accelerated vaccination campaign and; the establishment of ambulatory services for the treatment of mild to moderate Covid cases. We therefore modelled three datasets; a pre-Covid dataset up to the point of the Covid outbreak in the UK on 19 March 2020, a Covid dataset running from then to 19 July 2021, and one trained only on the period after SDEC was introduced, which runs from 17 November 2020 to 19 July 2021.

Visits were added to training, validation and test sets chronologically with the test set containing the most recent 20% of days, and the validation set the 10% before that, as shown in Supplementary Figure S3. This temporal split avoids any leak of future information into the past, and allows for a fairer test for problems with temporal drift that are commonly in real-time implementation^24^. See Supplementary Table S1 for a detailed explanation of how the training, validation and test sets were used to prepare models and regression equations, and to evaluate the predictions, at each step of the pipeline.

### Design of prediction pipeline

Our pipeline was designed to generate bed-level predictions from real-time patient-level data streams. We have four *prediction times* in the day and use data from an *observation window* to make predictions about the number of admissions in *prediction windows* of 4 and 8 hours after each prediction time (*italics* refer to the terminology of Lauritsen et al^23^). We constructed the aggregate predictions in a series of seven steps (see Figure 1). Figure 7 shows the temporal detail for each step at a hypothetical moment when four patients were in the ED at the prediction time, and an unknown number of patients could be expected to arrive after the prediction time and be admitted within the prediction window.

**Figure 7:**
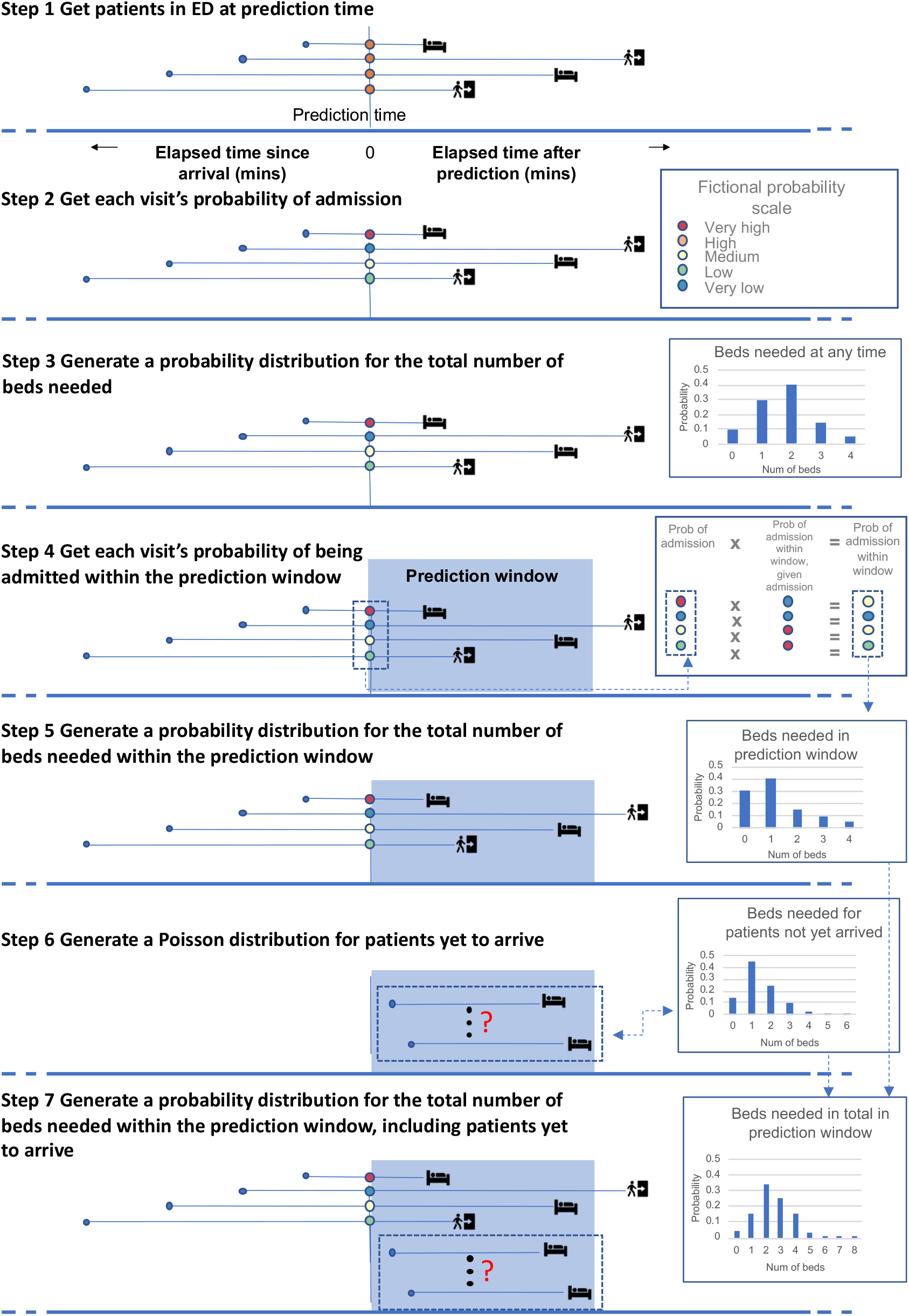
Temporal framing of each step in the process of making aggregated predictions.

#### Methods for Steps 1-2: Machine learning for predictions of individual probability of admission

Using Lauritsen et al.’s ^23^ terminology, samples were left-aligned for training and right-aligned for prediction, as shown in Figure 8. A series of 12 models, each trained on successively longer elapsed times in ED, was created. For example, model T60 was trained on all visits lasting more than 60 minutes and any data timestamped as recorded within 60 min of the patient’s arrival. For ease of interpretation, features are described here in four groups. The *visit features* included demographics, prior visit history and the nature and timing of the patients’ arrival. *Location features* included the patient’s location within the ED at the time of the prediction, and locations visited previously. *Observation and consults features* included triage scores, vital signs, the use of respiratory support and other data recorded by staff. Counts of each type of observation and latest values were used as features. Observations were also summarised by counting: how many observations were recorded; how many different types of observations were recorded; how many recording events took place; and how many specialist consultations were requested. *Pathology features* included requests for sets of lab tests, and latest results on a selection of tests clinically associated with acute illness. Pathology features were summarised by counting the number of out-of-range results that were higher, and the number that were lower, than the relevant target range for the patient. A glossary and descriptive statistics for features used T0, T90 and T240 models for the pre-Covid period are in Supplementary Table S2. Supplementary Table S3 has descriptive statistics for the T90 model in the various periods analysed (pre-Covid, during Covid and post-SDEC). Supplementary Figure S4 shows how a Covid surge feature was derived for the periods after the Covid outbreak.

**Figure 8:**
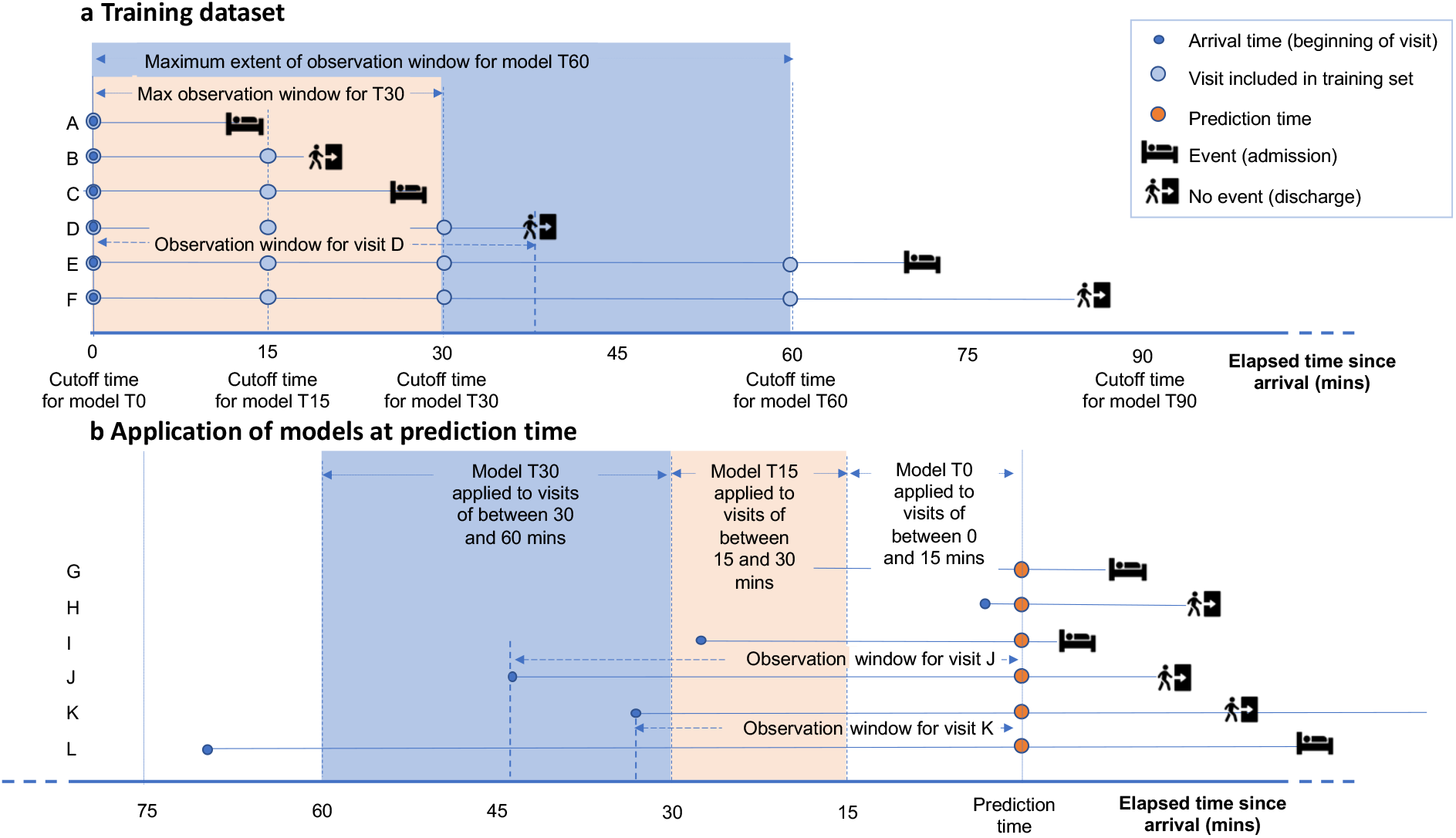
Temporal framing of ML models. A series of left-aligned datasets was constructed for training as shown in **a**. In each, the observation window began at the patient’s arrival time in the ED. Successive models were trained on longer observation windows. Model T0 had a zero minute cutoff, ie was given only the data known at the arrival time. Model T15 was trained on any data known up to 15 mins. Thus visit A, which lasted less than 15 min, only appears in T0. Model T30 was trained on any data known up to 30 min, so would only include data from visits D, E and F. To make predictions, right-aligned datasets of all patients in the ED at the time of prediction, as shown in **b**, were created. For each patient, the elapsed time of their visit determined which model was used to predict their probability of admission. Visit I began just less than 30 minutes before the prediction time, so this would be predicted by model T15. Visit K began just more than 30 minutes before the prediction time, so model T30 would be used for this visit.

The size and class balance of the dataset used for each model are shown in Supplementary Figure S5. As elapsed time increases, fewer patients remain and the proportion of admissions increases. An extreme gradient boosting (XGBoost) classifier^43^ was trained on each of the 12 datasets using a binary logistic loss function to generate a probability of admission. XGBoost was chosen due to its efficiency and capability of processing large datasets, its ability to handle missing data and imbalanced datasets, and (relative to other ML algorithms) its interpretability. Ten-fold cross-validation was used to find hyperparameters for each model, optimising for log loss, and the validation set used to assess held-out performance of individual-level models and the whole pipeline during development^44^. See Supplementary Note 3 for details on class balance, tuning and feature generation. To evaluate the individual-level probabilities, predictions were generated for all visits in the test set period and scored using log loss and AUROC as shown in the Results. Log loss was considered the most important metric, because, for input into the aggregation steps, accurate probabilities are more important than classification.

Predictions from XGBoost were compared with Random Forest (RF) and from Logistic Regression (LR). For RF and LR, missing values were imputed at the median. Lasso regularisation was applied to LR using the *Lasso and Elastic-Net Regularized Generalized Linear Models* (GLMNET) algorithm developed by Friedman et al ^32^. A series of Logistic Regression equations were created, using the lambda parameter – ranging from lambda.min, the optimal value, to lambda.4se, which is the most regularised model with a mean loss that was 4 standard errors away from that of the lambda.min model – to force greater model sparsity. More details are provided in Supplementary Note 8.

A post-hoc analysis evaluated the use of 3 models at Step 2, whereby visits were stratified as less than 90 min, less than 240 min and the remainder, with the T0, T90 and T240 models applied respectively (see Supplementary Note 7 for details).

#### Methods for Step 3: Aggregation into a probability distribution

At each prediction time, the probabilities of admission for every individual in the ED estimated at Step 2 were combined to give a predicted cumulative distribution function (cdf) for the aggregate number of admissions among this group (see for instance Utley et al^29^). The observed number of admissions associated with each prediction time was mapped to the midpoint of the relevant portion of its respective predicted cdf^45^. A plot of the cumulative distribution of these mapped observations against the predicted cdf was constructed to give a visual guide to the concurrence between the predicted distributions and the observations analogous to a QQ plot^44^.

#### Methods for Step 4-5: Survival analysis for time to admission and aggregation

Survival analysis applied to ED visit durations among admitted patients was used to estimate the probability of a patient that had been on the ED for a given time being admitted within the prediction window conditional on them being admitted eventually, with Cox regression used to adjust such probabilities to account for the time of the patient’s arrival (time of day, weekday or weekend, quarter of year) and the occupancy of the ED at that time. (See Supplementary Note 4 for more details and Supplementary Table S4 for regression coefficients.) This analysis was combined with the probabilities of admission estimated at Step 2 to give a probability for each patient in the ED at the prediction time of being admitted within the prediction window. These probabilities were then combined to give a predicted cumulative distribution function for the aggregate number of admissions within the prediction window among this group (as per Step 3).

#### Methods for Step 6-7: Poisson analysis for patients who have not yet arrived and aggregation

At each prediction time during the training set periods, a count was made of the number of patients not on the ED at the prediction time who were admitted via ED within the prediction window. A Poisson regression was fitted to the count data, with coefficients for the prediction time of day (06:00, 12:00, 16:00 and 22:00), quarter of year, and weekday or weekend. (See Supplementary Note 5 for more details and Supplementary Table S5 for regression coefficients.) The resulting coefficients were used to generate a probability distribution for the relevant prediction time of patients who have not yet arrived, and this was convoluted with the output from Step 5 to generate the final aggregated predictions of number of admissions within the prediction window, which was evaluated using QQ plots as in Step 3.

### Methods for final evaluation: comparison with benchmarks

Comparison with the commonly used six week rolling average was not straightforward, as this metric is for a 24 hour prediction window from midnight. A direct comparison with it is only possible for a prediction window that ends at midnight. Following practice in the hospital, the observed number of admissions up to 16:00 was subtracted from the daily rolling average to derive a prediction for the period from 16:00 to midnight for the benchmark. This was compared with the models’ 8 hour predictions at 16:00 for all report days in the test set, illustrated in Figure 6.

## Data Availability

The datasets analysed during the current study are not publicly available: due to reasonable privacy and security concerns, the underlying EHR data are not easily redistributable to researchers other than those engaged in approved research collaborations with the hospital.

## Code availability

The code used in this project is available at https://github.com/xxxxxx/real-time-admissions

## Acknowledgements

We acknowledge Lorraine Walton and Craig Wood for help with user requirements, Mohamed Abdalla, Jennifer Hunter, Dan Stein and Rebecca Wong for clinical advice, Nel Swanepoel for help building the real-time application, Sarah Keating, Tom Keen, Roma Klapaukh and Stefan Piatek for assistance with EMAP and Tim Roberts for modelling advice.

The work was funded by grants from the Wellcome Institutional Strategic Support Fund (ISSF) UCL and Partner Hospitals: AI in Healthcare Funding Call 2019, with an extension funded by NIHR UCLH Biomedical Research Centre HIGODS Theme (award number 182851). The contribution of MU and JG was funded under the National Institute for Health Research (Artificial Intelligence, Digitally adapted, hyper-local realtime bed forecasting to manage flow for NHS wards, AI_AWARD01786) and NHSX. The views expressed in this publication are those of the authors and not necessarily those of the National Institute for Health Research, NHSX or the Department of Health and Social Care.

## Author contributions

ZK performed the literature search, conducted the modelling and evaluation, contributed to the modelling design, generated the figures, and co-wrote the paper. SC and MU contributed to the modelling design and model evaluation, and co-wrote the paper. KL and JF contributed to the data interpretation and model evaluation and drafting. EK contributed to model evaluation. SC conceived the study with RS and SE, and secured funding. SH secured further project funding and contributed to model evaluation and drafting. JG contributed to the software engineering of the application. RS and SE contributed to drafting.

