## Supplementary material for "Machine Learning for Real-Time Aggregated Prediction of Hospital Admission for Emergency Patients": All supplemental material (from another version of this preprint: version 2, 2022-05-25)

Table of contents

**Supplementary Note 1: About patient flow in the ED department 3**

**Supplementary Note 2: Data pre-processing 4**

Figure S 1: Filtering steps for data pre-processing 5
Figure S 2: Histograms showing distribution of target variable 6
Figure S 3: Number of admissions and discharges by day, and allocation of visits into training, validation and test sets 7
Table S 1: Use of training, validation and test sets at each stage in the prediction pipeline 8
Figure S 4: Seven day rolling average deaths from Covid in the UK, derived from ONS data, with suggested cut-off for surge period 9
Table S 2: Glossary and descriptive statistics for variables used in three of the Pre-Covid models, with mean, standard deviation and number of missing values for each variable 10
Table S 3: Mean and standard deviations for features included in each dataset for Pre-Covid, Covid and Post-SDEC datasets for the T90 models only 14

**Supplementary Note 3: Machine Learning 16**

Figure S 5: Class balance in each dataset 17
Figure S 6: Calibration plots: Pre-Covid test set 18
Figure S 7: Calibration plots: Covid test set 18
Figure S 8: Calibration plots: Post-SDEC test set 19

**Supplementary Note 4: Survival analysis 20**

Figure S 9: QQ plots evaluating survival curves used for predicting probability of admission within prediction windows, for Pre-Covid, Covid and Post-SDEC datasets 21
Table S 4: Regression coefficients for Cox regression equations 22

**Supplementary Note 5: Poisson regression 23**

Figure S 10: QQ plots evaluating Poisson distributions used for predicting numbers of patients yet-to-arrive for Pre-Covid, Covid and Post-SDEC datasets 23
Table S 5: Exponentiated coefficients from Poisson regression 24

**Supplementary Note 6: Comparing results across the Pre-Covid, Covid and Post-SDEC datasets 25**

Figure S 11: Feature importance and performance for each model on test set 26

Figure S 12: Performance for each model on test sets of Pre-Covid, Covid and Post-SDEC datasets 27
Figure S 13: QQ plots evaluating distribution of models predicting number of beds needed for Pre-Covid, Covid and Post-SDEC datasets 28
Figure S 14: Survival curves for Pre-Covid, Covid and Post-SDEC training, validation and test sets 29
Figure S 15: Comparing model predictions with six week rolling average benchmark for number of admissions 8 hours after 16:00, including patients who yet to arrive, for Pre-Covid, Covid and Post-SDEC datasets 30

**Supplementary Note 7: Comparing 3 and 12 models 31**

Figure S 16: Calibration plots when using 3 models at Step 2 32
Figure S 17: Comparing 3 and 12 models at Step 3 33

**Supplementary Note 8: Examining alternatives to XGBoost 34**

Figure S 18: Number of non-zero coefficients (excluding the intercept) in each regularised LR model 35
Figure S 19: Feature importance for each model on test set using Random Forest 36
Figure S 20: Logistic regression with optimal penalty value, lambda.min: standardised regression coefficients with absolute value > 0.05 37
Figure S 21: Logistic regression with penalty value lambda.2se: standardised regression coefficients (all) 38
Figure S 22: Logistic regression with penalty value lambda.3se: standardised regression coefficients (all) 39
Figure S 23: Logistic regression with penalty value lambda.4se: standardised regression coefficients (all) 40

**Supplementary Note 9: Details of the application in use 41**

Figure S 24: Screenshots of the application in use 42

**Supplementary Note 10: TRIPOD Checklist for Prediction Model Development and Validation 43**

### Supplementary Note 1: About patient flow in the ED department

A typical patient is registered at the front desk, or by ambulance crew. Within 15 minutes or arrival they should be triaged, and have basic vitals documented, unless the nature of the presenting condition does not warrant taking vitals (eg a thumb injury). Ambulance arrivals and other urgent cases are assessed in RAT (Rapid Assessment and Triage), where they may have pathology orders issued, and requests for imaging and consultations with inpatient specialists. Patients are then assigned to Majors for further investigation and to await results. Patients typically stay in Majors until a treatment plan has been made. Alternatively, for minor illness or immediate treatment they are sent to the Urgent Treatment Centre (also called Minors). Patients requiring resuscitation or other intensive monitoring are sent to Resus. Paediatric patients have their own area of the ED. Up to March 2020, a location called Clinical Decision Unit (CDU) was used for patients who needed further observation or were waiting for results before discharge.

From November 2020 a Same-Day Emergency Care (SDEC) area was introduced, staffed with acute medicine specialists, with the intention of avoiding full hospital admission for patients who only need short-term care. This receives patients from the ED and those directly referred from the community. SDEC has taken over the operation of the previous Ambulatory Care Unit. Some patients who are ultimately admitted to a ward are routed to SDEC to manage ED flow at busy times. The Mental Health Stream (MHS) is a specialised location for patients with mental health issues.

Records on EMAP, the data source for this project, capture arrival and departure for each location. These timestamps are labelled as admission and discharge, and each admission time should equal the previous location discharge time. If a patient is required to wait, their location is recorded either as null or as Triage Waiting Area. Movements to imaging locations are not recorded in EMAP; the patient remains in their previously recorded location while imaging takes place.

### Supplementary Note 2: Data pre-processing

To identify visits involving the ED, various filtering steps were followed as shown in Figure S 2. These included steps to remove inconsistent data (such as data with equal admission and discharge timestamps, implying zero duration), visits that did not include any ED location, and visits with chronological mismatches in admission and discharge from locations. Patients under 18, self-discharges and patients who died in the ED were excluded. Visits with implausible lengths of stay (more than 48 hours) in one ED location were excluded, except where a patient stayed in the Mental Health Stream (for patients with mental health issues) for more than 48 hours, since this can happen.

The outcome variable is binary: whether a hospital visit ends in admission or not. There was no single variable identifying whether a patient was admitted. EMAP includes a patient class variable, which has categories of emergency and inpatient. However, a change from emergency to inpatient class was not suitable as the outcome variable; exploratory analysis showed that some patients arrived in ED with a class of inpatient, and others who were assigned a class of inpatient never visited a ward outside ED. Visits were classified as ending in admission if the patient went to a location other than ED, after being in the ED. Patients who were in a non-ED location prior to ED were only counted as admissions if they went to another non-ED location after their visit to ED. In total, of the 213,985 visits, 36,225 (20.7%) ended in admission by this definition. The distribution of the outcome variable at different report times, and in periods before and during Covid, is shown in Supplementary Figure S2.

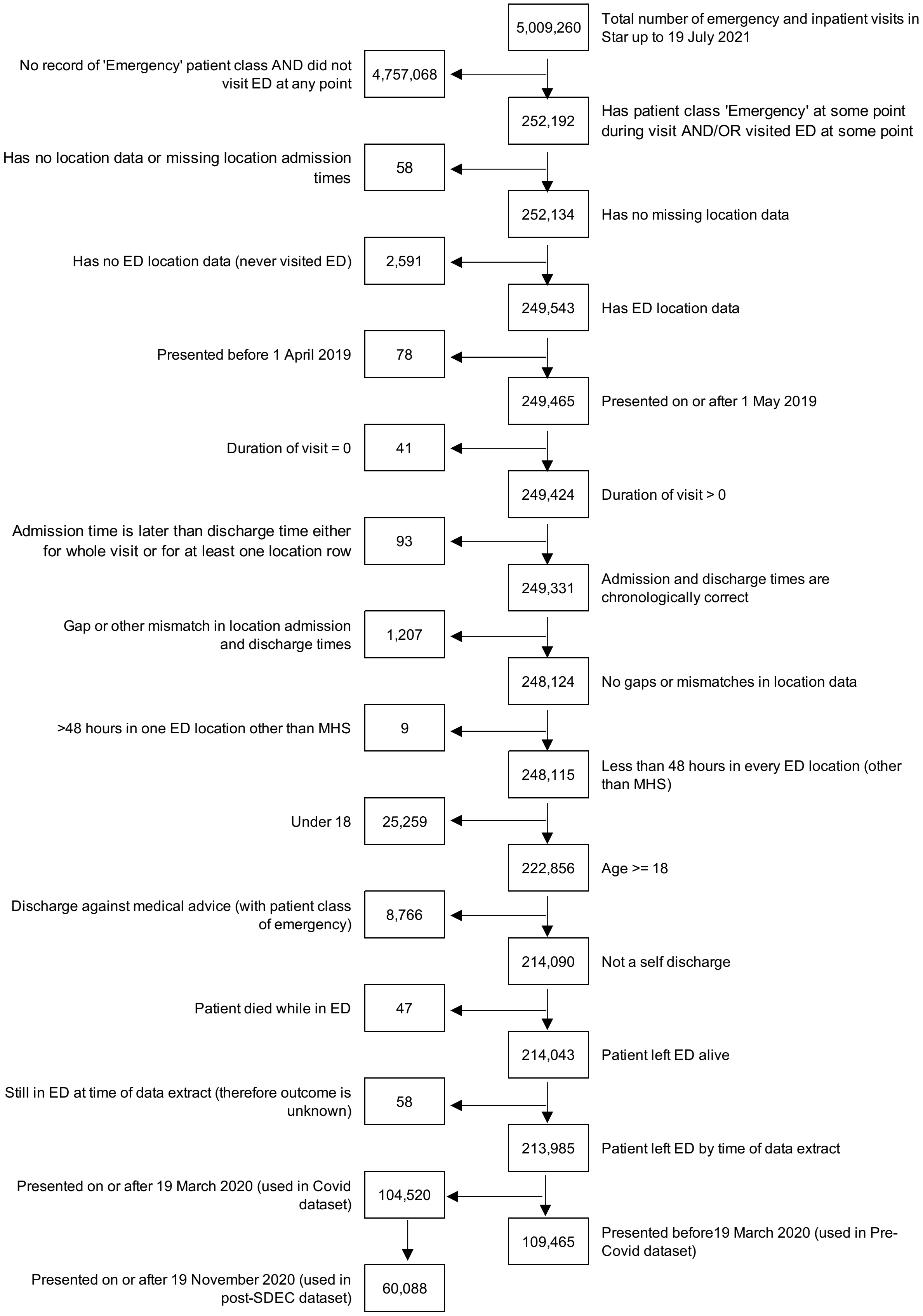


*Figure S 1: Filtering steps for data pre-processing*

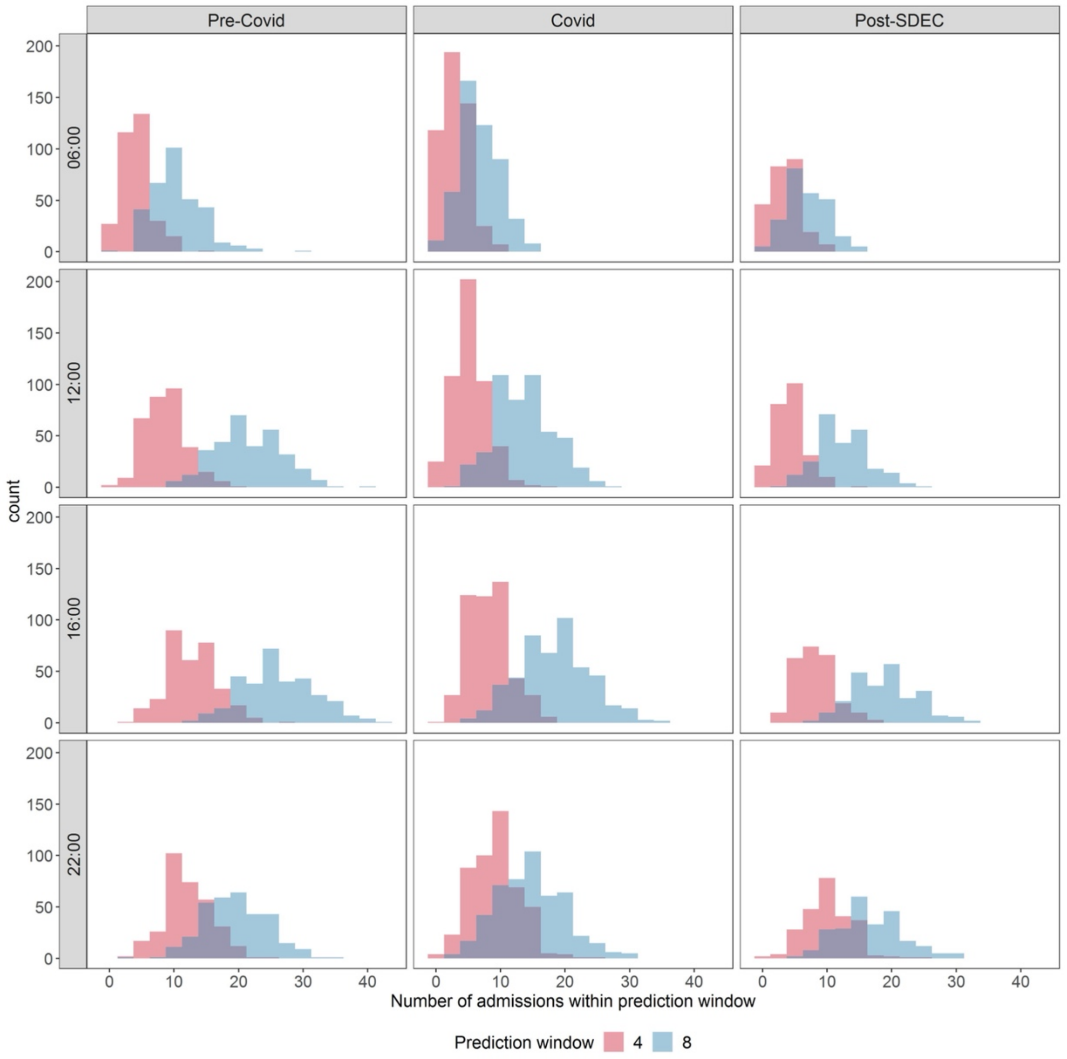


*Figure S 2: Histograms showing distribution of target variable*
Predictions were made at four points in the day (by row) and for three different datasets (by column). Predictions were made over two prediction windows (4 and 8 hours) shown in colour. The number of admissions within the prediction window increases later in the day, as a typical pattern of late discharges mean beds become available later.

The entire dataset of visits began on 1 May 2019 and ended on 20 July 2021. Patterns of presentation and care provision changed significantly over this period due to the Covid-19 pandemic. The dataset used for the main study runs up to 23:59 on 18 March 2020, prior to the point when the pandemic took hold in the UK. This end point was chosen by looking at patterns of presentations to the ED. From 19 March onward, there were fewer presentations, and a larger proportion were admitted, as shown in Figure S 3. As described in the main paper, two alternative datasets; a Covid dataset, which starts on 19 March 2019, and a post-SDEC dataset which is trained only on the period after SDEC was introduced (see Supplementary Note 1), were also presented for modelling.

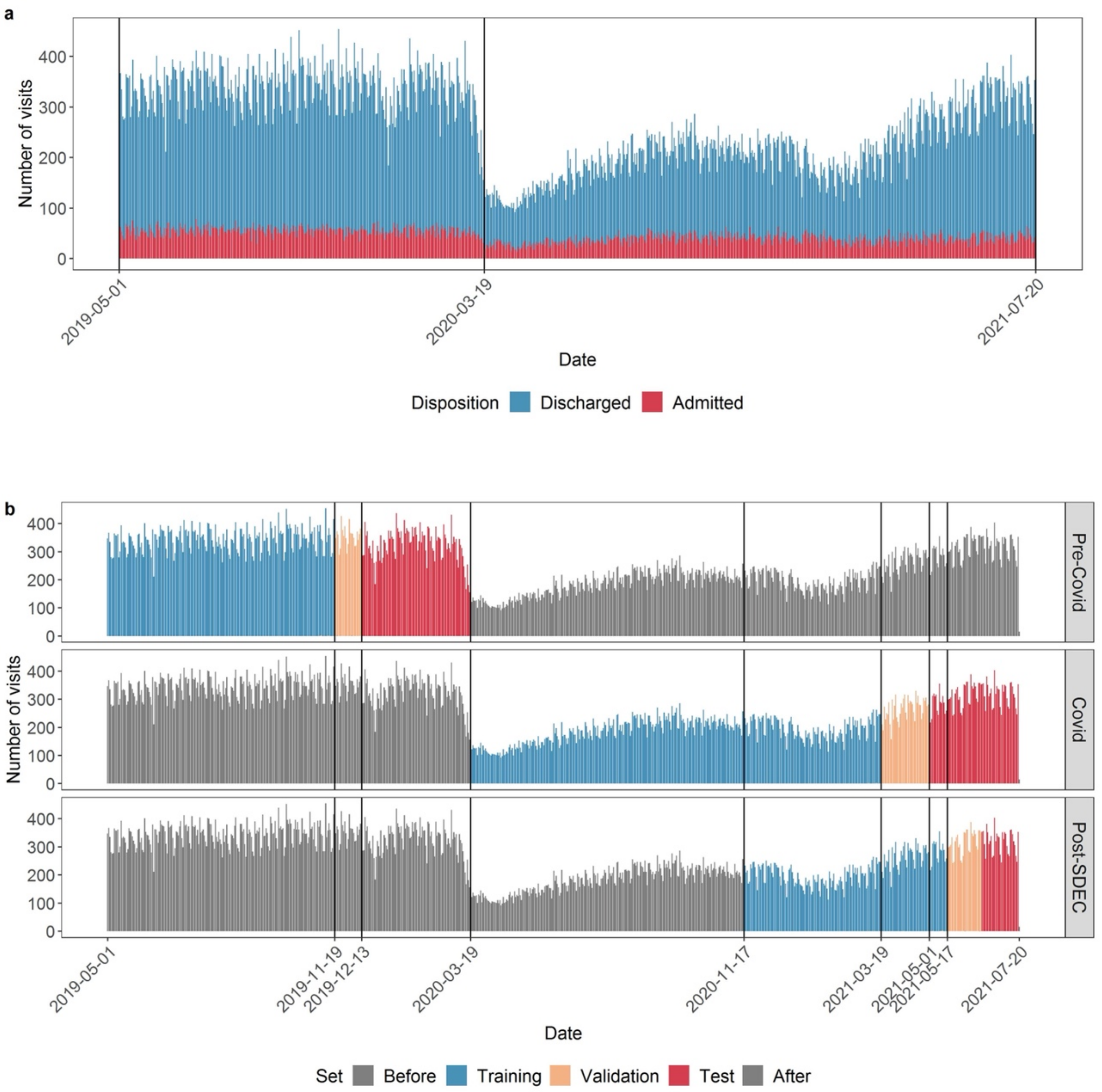


*Figure S 3: Number of admissions and discharges by day, and allocation of visits into training, validation and test sets*
**a** shows the number of patients presenting at the ED each day during the study period, coloured according to the number admitted and discharged. **b** shows how these visits were assigned chronologically within the pre-Covid, Covid and post-SDEC datasets, and within these into training, validation and test sets.

Visits were allocated to training, validation and test sets chronologically, with the test set containing approximately the most recent 20% of days in the period covered by the dataset, as shown in Figure S 3 above. Table S1 explains how the data were used in each of the seven steps of the prediction pipeline.

*Table S 1: Use of training, validation and test sets at each stage in the prediction pipeline*
In the table below, model preparation, prediction and model evaluation refer to stages of the work. Prediction was done using the validation set to assess model performance while the models were in development, and only done using the test set once the models were deemed to be complete. Model evaluation was done on the test set only.

| **Step in pipeline** | **Training set**<br>**All visits where date and time of ED arrival was within training set date range** | **Validation set**<br>**All visits where date and time of ED arrival was within validation set date range** | **Test set**<br>**All visits where date and time of ED arrival was within test set date range** |
|---|---|---|---|
| Step 1 Get patients in ED at prediction time | | **Prediction:** Retrieve each patient in the ED at each of the four report times (06:00, 12:00, 16:00, 22:00) at each day in the validation set period and create a right-aligned dataset (see Figure 7 in main paper for explanation of right- and left-alignment) | **Prediction:** As for Step 1 validation set |
| Step 2 Get each visit's probability of admission | **Model preparation:** All visits in training set period form a left-aligned dataset of training examples for the ML models. Multiple datasets are created, one for each model; eg every visit of least 15 min is included in dataset on which T15 model is trained. Hyperparameters tuned using 10-fold cross validation. | **Model preparation:** All visits in validation set period form multiple left-aligned datasets, used to assess ML model calibration while models under development<br>**Prediction:** Trained models applied to all validation set visits in right-aligned dataset from Step 1 to generate probabilities for next step. | **Prediction:** Trained models applied to all visits in right-aligned dataset from Step 1 to generate probabilities for next step<br>**Model evaluation:** All visits in test set form a left-aligned dataset of examples used to assess calibration of ML models (see Figure 3 in main paper and Figures S 6-8) and to report performance metrics in Figure 2 of main paper and Supplementary Note 6 |
| Step 3 Generate a probability distribution for the total number of beds needed | | **Prediction:** probabilities from Step 2 used to assess performance of aggregate predictions while models under development | **Prediction:** probabilities from Step 2 used to create an aggregate probability distribution<br>**Model evaluation:** these aggregate distributions are evaluated using QQ plots in Figure 5 of main paper and Supplementary Figure 11 (left column of each) |
| Step 4 Get each visit's probability of being admitted within the prediction window | **Model preparation:** All visits in training set that ended in admission used to generate survival curves and fit Cox regression equations as described in Supplementary Note 4 | **Model preparation:** Survival curve and Cox regression coefficients from training set applied to validation set (right-aligned) to calculate each visit's probability, given patient will be admitted, of being admitted within time window | **Prediction:** Survival curve and Cox regression coefficients from training set applied to test set (right-aligned) to calculate each visit's probability, given patient will be admitted, of being admitted within time window [1] |
| Step 5 Generate a probability distribution for the total number of beds needed within the prediction window | | **Prediction:** probabilities from Step 4 used to assess performance of aggregate predictions while models under development | **Prediction:** probabilities from Step 4 used to create an aggregate probability distribution<br>**Model evaluation:** these aggregate distributions are evaluated using QQ plots in Figure 5 of main paper and Supplementary Figure S13 (second column of each) |

[1] In a post-hoc analysis of the Covid and post-SDEC datasets, this step was modified. In addition to using survival curves from the training set to generate predictions of being admitted within the prediction window, a sliding observation window, over six weeks ending on the day of the prediction, was used to generate the survival curve and Cox regression coefficients.

| Step in pipeline | Training set<br>All visits where date and time of ED arrival was within training set date range | Validation set<br>All visits where date and time of ED arrival was within validation set date range | Test set<br>All visits where date and time of ED arrival was within test set date range |
|---|---|---|---|
| Step 6 Generate a Poisson distribution for patients who have not yet arrived | **Model preparation:** Training set used to generate frequencies of patients yet to arrive at each prediction time who were admitted within the prediction window. A Poisson equation was fitted to this dataset as described in Supplementary Note 6 | **Prediction:** Poisson regression coefficients from training set applied to each report time in the validation set to get a probability distribution for number of patients who have not yet arrived | **Prediction:** As for Step 6 validation set |
| Step 7 Generate a probability distribution for the total number of beds needed within the prediction window, including patients yet to arrive | | **Prediction:** probabilities generated by Steps 5 and 6 used to assess performance of aggregate predictions while models under development | **Prediction:** probabilities generated by Steps 5 and 6 used to create an aggregate probability distribution<br>**Model evaluation:** these aggregate distributions are evaluated using QQ plots in Figure 5 of main paper and Supplementary Figure S13 (right column(s) of each) |

To identify surges during the Covid and post-SDEC periods, data from the Office for National Statistics was used. Any visits which began on a day where the 7 day rolling average death rate was greater than 75 was considered a surge period, as shown in Figure S 4.

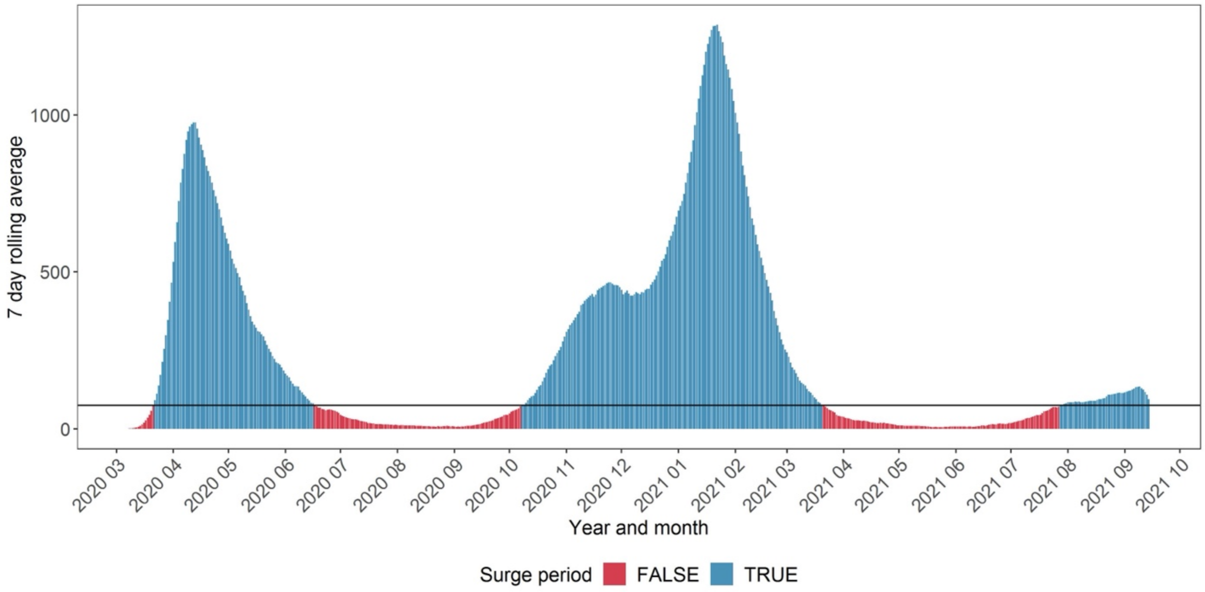


*Figure S 4: Seven day rolling average deaths from Covid in the UK, derived from ONS data, with suggested cut-off for surge period*
Source: https://coronavirus.data.gov.uk/details/deaths

*Table S 2: Glossary and descriptive statistics for variables used in three of the Pre-Covid models, with mean, standard deviation and number of missing values for each variable*

Mean, standard deviations and number of missing values are shown for three models. T0 includes all patients in the dataset. Since at 0 minutes after arrival, not much information is known about patients, data for T90 and T240 have also been included to show how the mean values change. By T240 (four hours after the visit began) only a quarter of the patients remain, with a higher proportion admitted. This table includes all features with an importance of > 0.01 in at least one of the Pre-Covid ML models, plus two other variables: whether patient is female (which was not important in any ML model but is important to include for comparison with other studies) and total visit duration (which was not included in the ML models because this would constitute data leakage). As would be expected, mean duration increases from T0 to T90 to T240 because the shorter visits drop out of the later models.

| Description | T0 | | T90 | | T240 | |
|---|---|---|---|---|---|---|
| | N | Proportion admitted | N | Proportion admitted | N | Proportion admitted |
| Number of visits whose duration was long enough to be included in model (proportion admitted) | 109465 | 0.16 | 87979 | 0.19 | 25881 | 0.43 |

| Feature / variable | Description | Data type (units) | T0 | | T90 | | T240 | |
|---|---|---|---|---|---|---|---|---|
| | | | Num missing | Mean (SD) | Num missing | Mean (SD) | Num missing | Mean (SD) |
| age | Age | Integer (years) | 0 | 43.14 (19.19) | 0 | 44.17 (19.57) | 0 | 50.44 (21.01) |
| age_gt70 | Age greater than 70 | Boolean | 0 | 0.12 | 0 | 0.14 | 0 | 0.23 |
| arrival_method_Ambnomedic | Patient arrived in an ambulance without a medical doctor | Integer (0 or 1) | 0 | 0.06 (0.23) | 0 | 0.06 (0.24) | 0 | 0.10 (0.3) |
| arrival_method_Ambulance | Patient arrived in an ambulance with a medical doctor | Integer (0 or 1) | 0 | 0.15 (0.36) | 0 | 0.17 (0.38) | 0 | 0.28 (0.45) |
| arrival_method_PublicTrans | Patient arrived by public transport | Integer (0 or 1) | 0 | 0.29 (0.45) | 0 | 0.28 (0.45) | 0 | 0.22 (0.42) |
| arrival_method_Walkin | Patient walked in | Integer (0 or 1) | 0 | 0.46 (0.5) | 0 | 0.45 (0.5) | 0 | 0.37 (0.48) |
| arrival_method_Other | Patient arrived by another method | Integer (0 or 1) | 0 | 0.05 (0.22) | 0 | 0.04 (0.19) | 0 | 0.03 (0.18) |
| arrival_window_0600.1200 | Arrival after 6 am and before or at 12 pm | Integer (0 or 1) | 0 | 0.25 (0.43) | 0 | 0.23 (0.42) | 0 | 0.17 (0.37) |
| arrival_window_1200.1600 | Arrival after 12 pm and before or at 4 pm | Integer (0 or 1) | 0 | 0.25 (0.43) | 0 | 0.27 (0.44) | 0 | 0.31 (0.46) |
| arrival_window_1600.2200 | Arrival after 4 pm and before or at 22 pm | Integer (0 or 1) | 0 | 0.34 (0.47) | 0 | 0.33 (0.47) | 0 | 0.30 (0.46) |
| arrival_window_2200.0600 | Arrival after 22 pm and before or at 6 am | Integer (0 or 1) | 0 | 0.16 (0.37) | 0 | 0.17 (0.38) | 0 | 0.22 (0.42) |
| female | Patient is female | Boolean | 0 | 0.52 | 0 | 0.52 | 0 | 0.52 |
| inpatient_prior_to_ED | Patient was an inpatient during the current hospital visit, prior to coming to ED (a tiny minority of vists) | Boolean | 0 | 0.00 | 0 | 0.00 | 0 | 0.00 |
| num_consults | Number of consultations from inpatient specialists requested | Integer | 0 | 0.00 (0.01) | 0 | 0.12 (0.34) | 0 | 0.52 (0.56) |
| num_prior_ED_visits | Number of prior visits to the ED | Integer | 0 | 0.69 (3.63) | 0 | 0.73 (3.82) | 0 | 0.99 (4.77) |
| num_prior_adm_after_ED | Number of prior admissions after a previous visit to the ED | Integer | 0 | 0.14 (0.63) | 0 | 0.15 (0.67) | 0 | 0.28 (0.91) |
| quarter_of_year_2 | Visit in the second quarter of the year (Apr to Jun) | Boolean | 0 | 0.19 (0.39) | 0 | 0.19 (0.39) | 0 | 0.18 (0.38) |
| quarter_of_year_3 | Visit in the third quarter of the year (Jul to Sep) | Boolean | 0 | 0.29 (0.45) | 0 | 0.28 (0.45) | 0 | 0.27 (0.44) |
| quarter_of_year_4 | Visit in the fourth quarter of the year (Oct to Dec) | Boolean | 0 | 0.29 (0.45) | 0 | 0.30 (0.46) | 0 | 0.32 (0.47) |

| Feature / variable | Description | Data type (units) | T0 | | T90 | | T240 | |
|---|---|---|---|---|---|---|---|---|
| | | | Num missing | Mean (SD) | Num missing | Mean (SD) | Num missing | Mean (SD) |
| time_from_pres_to_ED_arrival | Time difference between patient arrival in the hospital, and first recorded in a ED location | Float (mins) | 0 | 10.75 (817.04) | 0 | 11.77 (895.74) | 0 | 16.75 (823.52) |
| weekend | Whether weekday (0) or weekend (1) | Integer (0 or 1) | 0 | 0.25 | 0 | 0.24 | 0 | 0.21 |
| duration | Time difference between first arrival in a ED location, and first departure to a non-ED location (either to discharge or to a hospital ward) | Float (mins) | 0 | 216.41 (388.37) | 0 | 257.45 (422.96) | 0 | 472.50 (733.57) |
| current_ED.CDU | Current location is CDU (Clinical Decision Unit) | Integer (0 or 1) | NA | NA | 0 | 0.00 (0.03) | 0 | 0.06 (0.23) |
| current_ED.DIAGNOSTICS | Current location is Diagnostics | Integer (0 or 1) | 0 | 0.00 (0.01) | 0 | 0.01 (0.11) | 0 | 0.00 (0.02) |
| current_ED.MAJORS | Current location is Majors | Integer (0 or 1) | 0 | 0.00 (0.02) | 0 | 0.27 (0.45) | 0 | 0.56 (0.5) |
| current_ED.RAT | Current location is RAT (Rapid Assessment and Triage) | Integer (0 or 1) | 0 | 0.00 (0.05) | 0 | 0.05 (0.22) | 0 | 0.01 (0.09) |
| current_ED.RESUS | Current location is Resus (Resuscitation) | Integer (0 or 1) | 0 | 0.00 (0.02) | 0 | 0.05 (0.22) | 0 | 0.07 (0.26) |
| current_ED.MHS | Current location is MHS (Mental Health Stream) | Integer (0 or 1) | 0 | 0.00 (0) | 0 | 0.00 (0.06) | 0 | 0.04 (0.19) |
| current_ED.UTC | Current location is UTC (Urgent Treatment Centre) | Integer (0 or 1) | 0 | 0.00 (0.01) | 0 | 0.07 (0.25) | 0 | 0.09 (0.28) |
| current_ED.Waiting | Current location is Waiting | Integer (0 or 1) | 0 | 1.00 (0.06) | 0 | 0.54 (0.5) | 0 | 0.18 (0.38) |
| num_locations | Number of ED locations visited since arrival (multiple visits to the same location are counted separately) | Integer | 0 | 1.00 (0) | 0 | 2.03 (1.09) | 0 | 2.89 (1.09) |
| visited_ED.CDU | Patient has visited CDU (Clinical Decision Unit) | Boolean | NA | NA | 0 | 0.00 | 0 | 0.06 |
| visited_ED.DIAGNOSTICS | Patient has visited Diagnostics | Boolean | 0 | 0.00 | 0 | 0.16 | 0 | 0.21 |
| visited_ED.MAJORS | Patient has visited Majors | Boolean | 0 | 0.00 | 0 | 0.28 | 0 | 0.64 |
| visited_ED.RAT | Patient has visited RAT (Rapid Assessment and Triage) | Boolean | 0 | 0.00 | 0 | 0.27 | 0 | 0.44 |
| visited_ED.RESUS | Patient has visited Resus (Resuscitation) | Boolean | 0 | 0.00 | 0 | 0.06 | 0 | 0.11 |
| visited_ED.MHS | Patient has visited MHS (Mental Health Stream) | Boolean | 0 | 0.00 | 0 | 0.00 | 0 | 0.04 |
| visited_ED.UTC | Patient has visited UTC (Urgent Treatment Centre) | Boolean | 0 | 0.00 | 0 | 0.07 | 0 | 0.12 |
| visited_ED.Waiting | Patient has visited Waiting | Boolean | 0 | 1.00 | 0 | 1.00 | 0 | 1.00 |
| has_obs | Patient has observations recorded | Integer (0 or 1) | 0 | 0.01 (0.1) | 0 | 0.98 (0.13) | 0 | 0.99 (0.07) |
| latest_Heartrate | Latest heart rate | Integer (beats/min) | 108633 | 84.71 (22.33) | 18981 | 84.51 (16.58) | 2009 | 81.39 (16.3) |
| latest_ManchesterTriageScore_Blue | Latest Manchester Triage Score is blue (non-urgent) | Boolean | 0 | 0.00 (0.01) | 0 | 0.03 (0.17) | 0 | 0.01 (0.11) |
| latest_ManchesterTriageScore_Green | Latest Manchester Triage Score is green (standard) | Boolean | 0 | 0.00 (0.03) | 0 | 0.44 (0.5) | 0 | 0.26 (0.44) |
| latest_ManchesterTriageScore_Not.recorded | No Manchester Triage Score has been recorded | Boolean | 0 | 1.00 (0.04) | 0 | 0.08 (0.26) | 0 | 0.07 (0.25) |
| latest_ManchesterTriageScore_Orange | Latest Manchester Triage Score is orange (very urgent) | Boolean | 0 | 0.00 (0.02) | 0 | 0.14 (0.35) | 0 | 0.24 (0.43) |
| latest_ManchesterTriageScore_Red | Latest Manchester Triage Score is red (immediate) | Boolean | 0 | 0.00 (0.01) | 0 | 0.00 (0.05) | 0 | 0.00 (0.05) |
| latest_ManchesterTriageScore_Yellow | Latest Manchester Triage Score is yellow (urgent) | Boolean | 0 | 0.00 (0.02) | 0 | 0.31 (0.46) | 0 | 0.42 (0.49) |

| Feature / variable | Description | Data type (units) | T0 | | T90 | | T240 | |
|---|---|---|---|---|---|---|---|---|
| | | | Num missing | Mean (SD) | Num missing | Mean (SD) | Num missing | Mean (SD) |
| latest_NEWSscore | Latest NEWS (National Early Warning Score) value; ranges from 0 to 3 | Integer | 108808 | 2.18 (2.7) | 41875 | 1.12 (1.61) | 4479 | 1.07 (1.58) |
| latest_ObjectivePainScore_ Mild | Latest Objective Pain Score is mild | Boolean | 0 | 0.00 (0.02) | 0 | 0.15 (0.36) | 0 | 0.09 (0.29) |
| latest_ObjectivePainScore_ Moderate | Latest Objective Pain Score is moderate | Boolean | 0 | 0.00 (0.01) | 0 | 0.05 (0.22) | 0 | 0.05 (0.22) |
| latest_ObjectivePainScore_ Nil | Latest Objective Pain Score is nil | Boolean | 0 | 0.00 (0.02) | 0 | 0.08 (0.27) | 0 | 0.05 (0.22) |
| latest_ObjectivePainScore_ Not.recorded | Latest Objective Pain Score is not recorded | Boolean | 0 | 1.00 (0.03) | 0 | 0.71 (0.45) | 0 | 0.79 (0.41) |
| latest_ObjectivePainScore_ Severe | Latest Objective Pain Score is severe | Boolean | 0 | 0.00 (0) | 0 | 0.01 (0.07) | 0 | 0.01 (0.09) |
| latest_Respiratoryrate | Latest respiratory rate | Integer (breaths/min) | 108675 | 18.77 (4.12) | 26073 | 17.44 (3.43) | 2718 | 17.65 (2.9) |
| latest_TempinCelsius | Latest temperature | Float (degrees celcius) | 108697 | 36.69 (0.76) | 22281 | 36.75 (0.67) | 2065 | 36.77 (0.65) |
| num_ACVPU | Number of times the AVCPU score has been recorded. ACVPU is a scale used to assess and track a patient's neurological status and level of consciousness Values are alert, confusion, voice, pain, unresponsive | Integer | 0 | 0.01 (0.09) | 0 | 0.76 (0.84) | 0 | 2.27 (1.69) |
| num_BestMotorResponse | Number of times the patient's best motor response has been recorded | Integer | 0 | 0.00 (0.06) | 0 | 0.21 (0.65) | 0 | 0.65 (1.32) |
| num_BestVerbalResponse | Number of times the patient's best verbal response has been recorded | Integer | 0 | 0.00 (0.06) | 0 | 0.21 (0.66) | 0 | 0.65 (1.32) |
| num_Bloodpressure | Number of times blood pressure has been recorded | Integer | 0 | 0.01 (0.1) | 0 | 0.81 (0.91) | 0 | 2.39 (1.78) |
| num_Heartrate | Number of times heart rate has been recorded | Integer | 0 | 0.01 (0.1) | 0 | 1.10 (0.86) | 0 | 2.57 (1.71) |
| num_ManchesterTriageScor e | Number of times Manchester Triage Score has been recorded | Integer | 0 | 0.00 (0.04) | 0 | 0.97 (0.35) | 0 | 0.99 (0.37) |
| num_Oxygendeliverydevice | Number of times the use of an oxygen delivery device has been recorded | Integer | 0 | 0.00 (0.04) | 0 | 0.03 (0.28) | 0 | 0.15 (0.78) |
| num_Oxygentherapyflowrat e | Number of times oxygen therapy flow rate has been recorded | Integer | 0 | 0.00 (0.03) | 0 | 0.03 (0.26) | 0 | 0.14 (0.75) |
| num_Painscoreverbalatrest | Number of times verbal pain score at rest has been recorded | Integer | 0 | 0.00 (0.03) | 0 | 0.33 (0.61) | 0 | 0.96 (1.29) |
| num_Painscoreverbalonmov ement | Number of times verbal pain score on movement has been recorded | Integer | 0 | 0.00 (0.03) | 0 | 0.25 (0.51) | 0 | 0.74 (1.06) |
| num_RPupilReaction | Number of times a R pupil reaction has been recorded | Integer | 0 | 0.00 (0.06) | 0 | 0.10 (0.44) | 0 | 0.31 (0.91) |
| num_Roomairoroxygen | Number of times the use of room air or oxygen has been recorded | Integer | 0 | 0.01 (0.09) | 0 | 0.76 (0.84) | 0 | 2.25 (1.68) |
| num_TempinCelsius | Number of times the patient's temperature has been recorded | Integer | 0 | 0.01 (0.09) | 0 | 1.02 (0.82) | 0 | 2.48 (1.6) |

| Feature / variable | Description | Data type (units) | T0 | | T90 | | T240 | |
|---|---|---|---|---|---|---|---|---|
| | | | Num missing | Mean (SD) | Num missing | Mean (SD) | Num missing | Mean (SD) |
| num_allpressureareasobserved | Number of times someone has recorded that all pressure areas have been observed | Integer | 0 | 0.00 (0.02) | 0 | 0.00 (0.05) | 0 | 0.01 (0.11) |
| num_news_high | Number of times the NEWS score has been recorded as high | Integer | 0 | 0.00 (0.03) | 0 | 0.02 (0.21) | 0 | 0.08 (0.51) |
| num_obs | Number of observations that have been recorded, of any type, at any time | Integer | 0 | 0.10 (1.16) | 0 | 11.55 (10.09) | 0 | 28.83 (21.22) |
| num_obs_events | Number of timestamps at which observations have been recorded | Integer | 0 | 0.01 (0.12) | 0 | 2.20 (1.24) | 0 | 4.16 (2.26) |
| num_obs_type | Number of types of observations have been recorded | Integer | 0 | 0.08 (0.97) | 0 | 7.68 (4.9) | 0 | 11.12 (4.51) |
| latest_ALB | Latest lab result for albumin, a marker of nutritional status and liver function | Float (g/L) | 109453 | 39.08 (6.86) | 83255 | 43.94 (5.36) | 17221 | 43.77 (5.43) |
| latest_CREA | Latest lab result for creatinine, a marker of renal function | Float (umol/L) | 109452 | 82.85 (37.77) | 79004 | 88.38 (50.66) | 9808 | 88.60 (61.55) |
| latest_HCTU | Latest lab result for hematocrit, which may suggest dehydration if high, and significant crystalloid fluid replacement if low | Float (L/L) | 109458 | 0.31 (0.05) | 71818 | 0.40 (0.05) | 9022 | 0.39 (0.05) |
| latest_HTRT | Latest lab result for high sensitivity troponin t, where a high value indicates acute coronary syndrome, or other cardiac disease, sepsis, renal dysfunction | Float (ng/L) | 109464 | 37.00 | 87147 | 29.26 (125.74) | 23858 | 35.68 (220.48) |
| latest_K | Latest lab result for potassium, an important ion in blood. Abnormal values necessitate treatment | Float (mmol/L) | 109456 | 4.12 (0.47) | 81327 | 4.28 (0.47) | 13924 | 4.31 (0.49) |
| latest_NA | Latest lab result for sodium, where abonormal values suggest various issues including endocrine dysfunction | Float (mmol/L) | 109454 | 137.45 (5.13) | 84830 | 138.75 (4.05) | 19694 | 138.35 (4.34) |
| latest_WCC | Latest lab result for white cell count, where abnormal values indicate infection or haemotological disease | Float (10^9/L) | 109458 | 7.39 (4.48) | 71823 | 9.28 (5.28) | 9026 | 9.56 (7.17) |
| num_battery | Number of batteries of lab tests orders | Integer | 0 | 0.00 (0.06) | 0 | 1.46 (2.62) | 0 | 4.18 (3.17) |
| num_oor_high | Number of lab results that are out of target range and high | Integer | 0 | 0.00 (0.03) | 0 | 0.38 (1.17) | 0 | 1.84 (2.33) |
| num_oor_low | Number of lab results that are out of target range and low | Integer | 0 | 0.00 (0.03) | 0 | 0.28 (0.96) | 0 | 1.23 (1.87) |
| num_oor_low_HCTU | Number of lab results for hematocrit that are out of target range and low. Low values suggest resuscitation | Integer | 0 | 0.00 (0) | 0 | 0.03 (0.16) | 0 | 0.11 (0.31) |
| req_battery_BC | Request for blood culture battery, implies that sepsis is suspected | Boolean | 0 | 0.00 | 0 | 0.02 | 0 | 0.08 |
| req_battery_BON | Request for bone profile battery, implies that hyper/hypo calcaemia is suspected | Boolean | 0 | 0.00 | 0 | 0.01 | 0 | 0.05 |
| req_battery_CRP | Request for C-reactive protein battery, a non specific marker of inflammation, suggests general unwellness | Boolean | 0 | 0.00 | 0 | 0.24 | 0 | 0.66 |
| req_battery_CSNF | Request for blood clotting battery, suggests bleeding disorders | Boolean | 0 | 0.00 | 0 | 0.08 | 0 | 0.23 |
| req_battery_DDIT | Request for D-Dimer battery, implies signfiicant respiratory symptoms/chest pain/low oxygen | Boolean | 0 | 0.00 | 0 | 0.01 | 0 | 0.04 |

*Table S 3: Mean and standard deviations for features included in each dataset for Pre-Covid, Covid and Post-SDEC datasets for the T90 models only*
See Table S 2 for descriptions of what each feature represents. Features are included in this table if they had an importance score of > 0.01 in any of the three datasets for the T90 model.

| **Model N** | **Pre-Covid T90** | | **Covid T90** | | **Post-SDEC T90** | |
|---|---|---|---|---|---|---|
| | **Discharged** | **Admitted (prop of total visits)** | **Discharged** | **Admitted (prop of total visits)** | **Discharged** | **Admitted (prop of total visits)** |
| Number in model | 71092 | 16887 (0.19) | 59841 | 18049 (0.23) | 37496 | 9477 (0.20) |
| | **Pre-Covid T90** | | **Covid T90** | | **Post-SDEC T90** | |
| | **Discharged mean (SD)** | **Admitted mean (SD)** | **Discharged mean (SD)** | **Admitted mean (SD)** | **Discharged mean (SD)** | **Admitted mean (SD)** |
| age | 40.69 (17.38) | 58.82 (21.41) | 43.27 (17.58) | 58.19 (20.7) | 43.40 (17.81) | 58.46 (20.55) |
| age_gt70 | 0.08 | 0.38 | 0.10 | 0.35 | 0.10 | 0.35 |
| arrival_method_Ambnomedic | 0.04 (0.21) | 0.14 (0.35) | 0.03 (0.18) | 0.10 (0.3) | 0.03 (0.18) | 0.11 (0.31) |
| arrival_method_Ambulance | 0.12 (0.33) | 0.37 (0.48) | 0.15 (0.35) | 0.42 (0.49) | 0.13 (0.33) | 0.40 (0.49) |
| arrival_method_PublicTrans | 0.31 (0.46) | 0.17 (0.38) | 0.29 (0.45) | 0.17 (0.38) | 0.28 (0.45) | 0.16 (0.37) |
| arrival_method_Walkin | 0.49 (0.5) | 0.28 (0.45) | 0.48 (0.5) | 0.27 (0.44) | 0.51 (0.5) | 0.29 (0.45) |
| arrival_window_0600.1200 | 0.24 (0.42) | 0.20 (0.4) | 0.24 (0.43) | 0.20 (0.4) | 0.25 (0.43) | 0.19 (0.39) |
| arrival_window_1200.1600 | 0.26 (0.44) | 0.28 (0.45) | 0.25 (0.43) | 0.29 (0.45) | 0.24 (0.43) | 0.29 (0.46) |
| arrival_window_1600.2200 | 0.33 (0.47) | 0.33 (0.47) | 0.35 (0.48) | 0.34 (0.47) | 0.36 (0.48) | 0.35 (0.48) |
| arrival_window_2200.0600 | 0.17 (0.37) | 0.18 (0.39) | 0.16 (0.37) | 0.18 (0.38) | 0.15 (0.36) | 0.17 (0.38) |
| covid_surge | NA | NA | 0.40 | 0.48 | 0.37 | 0.49 |
| female | 0.53 | 0.50 | 0.52 | 0.50 | 0.53 | 0.50 |
| inpatient_prior_to_ED | 0.00 | 0.00 | 0.00 | 0.01 | 0.00 | 0.01 |
| num_consults | 0.06 (0.24) | 0.41 (0.52) | 0.16 (0.38) | 0.49 (0.54) | 0.19 (0.41) | 0.51 (0.54) |
| num_prior_ED_visits | 0.72 (4.14) | 0.80 (1.95) | 2.00 (7.42) | 2.35 (4.66) | 2.01 (6.58) | 2.63 (5.28) |
| num_prior_adm_after_ED | 0.08 (0.47) | 0.44 (1.15) | 0.30 (1.27) | 1.20 (2.8) | 0.33 (1.39) | 1.35 (3.22) |
| other_gender | 0.00 | 0.00 | 0.00 | 0.00 | 0.00 | 0.00 |
| quarter_of_year_2 | 0.19 (0.39) | 0.19 (0.39) | 0.41 (0.49) | 0.35 (0.48) | 0.48 (0.5) | 0.38 (0.49) |
| time_from_pres_to_ED_arrival | 4.02 (805.57) | 44.35 (1202.86) | 6.85 (496.84) | 149.12 (3257.41) | 8.19 (596.44) | 189.58 (3215.92) |
| weekend | 0.24 | 0.25 | 0.25 | 0.24 | 0.24 | 0.23 |
| current_ED.CDU | 0.00 (0.03) | 0.00 (0.03) | NA | NA | NA | NA |
| current_ED.MAJORS | 0.21 (0.41) | 0.53 (0.5) | 0.39 (0.49) | 0.67 (0.47) | 0.37 (0.48) | 0.63 (0.48) |
| current_ED.RAT | 0.04 (0.2) | 0.10 (0.3) | 0.07 (0.25) | 0.06 (0.24) | 0.06 (0.23) | 0.07 (0.26) |
| current_ED.RESUS | 0.02 (0.12) | 0.19 (0.39) | 0.02 (0.13) | 0.09 (0.28) | 0.02 (0.12) | 0.08 (0.27) |
| current_ED.MHS | 0.01 (0.07) | 0.00 (0.01) | 0.01 (0.08) | 0.00 (0.01) | 0.01 (0.09) | 0.00 (0.01) |
| current_ED.UTC | 0.08 (0.27) | 0.03 (0.16) | 0.33 (0.47) | 0.06 (0.24) | 0.37 (0.48) | 0.02 (0.14) |
| current_ED.Waiting | 0.64 (0.48) | 0.13 (0.34) | 0.11 (0.31) | 0.02 (0.12) | 0.05 (0.21) | 0.01 (0.1) |
| current_SDEC | NA | NA | 0.07 (0.25) | 0.10 (0.3) | 0.10 (0.31) | 0.19 (0.39) |
| num_locations | 1.87 (1.07) | 2.70 (0.88) | 2.39 (0.76) | 2.65 (0.68) | 2.46 (0.71) | 2.73 (0.68) |
| visited_ED.CDU | 0.00 | 0.00 | 0.00 | 0.00 | 0.00 | 0.00 |
| visited_ED.DIAGNOSTICS | 0.15 | 0.20 | 0.00 | 0.00 | 0.00 | 0.00 |
| visited_ED.MAJORS | 0.22 | 0.55 | 0.40 | 0.70 | 0.39 | 0.67 |
| visited_ED.RAT | 0.22 | 0.51 | 0.47 | 0.61 | 0.47 | 0.67 |
| visited_ED.RESUS | 0.02 | 0.22 | 0.03 | 0.10 | 0.02 | 0.10 |
| visited_ED.MHS | 0.01 | 0.00 | 0.01 | 0.00 | 0.01 | 0.00 |
| visited_ED.UTC | 0.08 | 0.03 | 0.35 | 0.08 | 0.40 | 0.03 |
| visited_SDEC | 0.00 | 0.00 | 0.07 | 0.10 | 0.11 | 0.19 |
| latest_Heartrate | 83.94 (15.47) | 86.47 (19.82) | 84.38 (15.21) | 87.39 (18.73) | 84.67 (15.04) | 87.84 (18.52) |
| latest_ManchesterTriageScore_Green | 0.51 (0.5) | 0.16 (0.36) | 0.44 (0.5) | 0.16 (0.37) | 0.45 (0.5) | 0.16 (0.37) |

| | | | | | | |
|---|---|---|---|---|---|---|
| latest_ManchesterTriageScore_Not.recorded | 0.06 (0.24) | 0.12 (0.33) | 0.03 (0.16) | 0.02 (0.14) | 0.04 (0.2) | 0.03 (0.16) |
| latest_ManchesterTriageScore_Orange | 0.09 (0.29) | 0.33 (0.47) | 0.13 (0.34) | 0.38 (0.49) | 0.13 (0.33) | 0.38 (0.49) |
| latest_ManchesterTriageScore_Yellow | 0.30 (0.46) | 0.37 (0.48) | 0.37 (0.48) | 0.41 (0.49) | 0.36 (0.48) | 0.41 (0.49) |
| latest_NEWSscore | 0.81 (1.14) | 1.82 (2.19) | 0.77 (1.11) | 2.01 (2.37) | 0.74 (1.08) | 2.01 (2.34) |
| latest_ObjectivePainScore_Moderate | 0.05 (0.23) | 0.05 (0.21) | 0.18 (0.39) | 0.20 (0.4) | 0.19 (0.39) | 0.21 (0.41) |
| latest_Respiratoryrate | 17.09 (3.24) | 18.51 (3.74) | 17.49 (2.51) | 19.19 (4.03) | 17.47 (2.48) | 19.11 (3.81) |
| latest_TempinCelsius | 36.71 (0.63) | 36.86 (0.78) | 36.68 (0.59) | 36.87 (0.77) | 36.66 (0.54) | 36.86 (0.81) |
| num_ACVPU | 0.61 (0.73) | 1.38 (1) | 0.82 (0.81) | 1.50 (0.89) | 0.77 (0.78) | 1.44 (0.88) |
| num_BestMotorResponse | 0.14 (0.51) | 0.52 (1.01) | 0.71 (1.01) | 1.39 (1.24) | 0.59 (0.9) | 1.18 (1.13) |
| num_Bloodpressure | 0.65 (0.79) | 1.46 (1.07) | 1.17 (1.16) | 2.08 (1.18) | 1.08 (1.12) | 2.01 (1.16) |
| num_Heartrate | 0.99 (0.76) | 1.59 (1.06) | 1.43 (0.89) | 1.91 (0.97) | 1.39 (0.85) | 1.84 (0.95) |
| num_ManchesterTriageScore | 0.98 (0.33) | 0.93 (0.41) | 0.98 (0.2) | 1.00 (0.19) | 0.97 (0.22) | 0.98 (0.19) |
| num_Oxygendeliverydevice | 0.00 (0.1) | 0.14 (0.58) | 0.01 (0.13) | 0.25 (0.75) | 0.01 (0.13) | 0.25 (0.74) |
| num_Oxygentherapyflowrate | 0.00 (0.1) | 0.14 (0.58) | 0.01 (0.13) | 0.25 (0.75) | 0.01 (0.13) | 0.25 (0.74) |
| num_Painscoreverbalatrest | 0.00 (0.1) | 0.13 (0.55) | 0.01 (0.13) | 0.22 (0.7) | 0.01 (0.12) | 0.22 (0.68) |
| num_Painscoreverbalonmovement | 0.29 (0.57) | 0.50 (0.76) | 0.49 (0.73) | 0.80 (0.91) | 0.45 (0.7) | 0.76 (0.87) |
| num_RPupilReaction | 0.21 (0.47) | 0.40 (0.64) | 0.40 (0.63) | 0.67 (0.78) | 0.37 (0.61) | 0.65 (0.75) |
| num_Roomairoroxygen | 0.07 (0.33) | 0.24 (0.73) | 0.10 (0.39) | 0.19 (0.56) | 0.08 (0.35) | 0.16 (0.5) |
| num_TempinCelsius | 0.91 (0.74) | 1.52 (0.95) | 2.03 (1.49) | 2.81 (1.77) | 1.47 (0.92) | 1.91 (1.08) |
| num_allpressureareasobserved | 0.00 (0.02) | 0.01 (0.1) | 0.00 (0.02) | 0.00 (0.06) | 0.00 (0.03) | 0.00 (0.06) |
| num_news_high | 0.00 (0.07) | 0.10 (0.44) | 0.00 (0.08) | 0.15 (0.55) | 0.00 (0.08) | 0.14 (0.53) |
| num_obs | 9.87 (8.26) | 18.65 (13.43) | 18.93 (12.41) | 28.29 (13.99) | 17.78 (11.41) | 26.20 (13.22) |
| num_obs_events | 2.03 (1.11) | 2.90 (1.49) | 2.64 (1.52) | 3.79 (1.65) | 2.53 (1.42) | 3.57 (1.57) |
| num_obs_type | 6.94 (4.74) | 10.76 (4.34) | 11.94 (5.58) | 15.07 (4.09) | 12.06 (5.51) | 15.14 (4.36) |
| latest_ALB | 46.02 (3.96) | 42.21 (5.75) | 45.06 (4.25) | 41.35 (5.95) | 44.83 (4.24) | 41.03 (5.91) |
| latest_CREA | 80.43 (26.48) | 97.26 (67.09) | 80.37 (27.56) | 96.39 (62.34) | 80.48 (29.43) | 95.14 (62.73) |
| latest_HCTU | 0.41 (0.04) | 0.39 (0.06) | 0.40 (0.05) | 0.38 (0.06) | 0.40 (0.05) | 0.39 (0.06) |
| latest_HTRT | 13.86 (89.45) | 55.72 (167.9) | 20.12 (175.09) | 41.67 (101.77) | 15.22 (67.32) | 45.59 (117.93) |
| latest_K | 4.28 (0.4) | 4.29 (0.54) | 4.22 (0.38) | 4.25 (0.54) | 4.22 (0.39) | 4.24 (0.54) |
| latest_Lac | NA | NA | 1.27 (0.76) | 1.68 (1.51) | 1.27 (0.76) | 1.68 (1.51) |
| latest_NA | 139.40 (3.15) | 138.16 (4.65) | 138.81 (3.01) | 137.16 (4.66) | 138.63 (3.03) | 136.83 (4.61) |
| latest_WCC | 8.59 (4.1) | 10.18 (6.4) | 8.17 (3.51) | 10.08 (6.5) | 8.08 (3.5) | 10.11 (7.13) |
| latest_pCO2 | NA | NA | 6.13 (0.9) | 5.92 (1.13) | 6.13 (0.9) | 5.92 (1.13) |
| latest_pH | NA | NA | 7.40 (0.04) | 7.40 (0.07) | 7.40 (0.04) | 7.40 (0.07) |
| num_battery | 0.95 (2.1) | 3.61 (3.38) | 1.74 (2.59) | 4.63 (3.74) | 1.65 (2.61) | 4.74 (4.06) |
| num_oor_high | 0.19 (0.76) | 1.16 (1.99) | 0.36 (1.02) | 1.54 (2.26) | 0.32 (0.97) | 1.45 (2.28) |
| num_oor_low | 0.14 (0.64) | 0.87 (1.63) | 0.30 (0.93) | 1.22 (1.92) | 0.28 (0.92) | 1.15 (1.91) |
| num_oor_low_HCTU | 0.01 (0.1) | 0.09 (0.29) | 0.02 (0.15) | 0.13 (0.34) | 0.02 (0.15) | 0.12 (0.33) |
| req_battery_BC | 0.01 | 0.06 | 0.01 | 0.11 | 0.01 | 0.12 |
| req_battery_BON | 0.00 | 0.06 | 0.02 | 0.11 | 0.02 | 0.11 |
| req_battery_CRP | 0.17 | 0.55 | 0.30 | 0.63 | 0.27 | 0.59 |
| req_battery_CSNF | 0.04 | 0.26 | 0.13 | 0.43 | 0.12 | 0.42 |
| req_battery_DDIT | 0.01 | 0.02 | 0.06 | 0.12 | 0.07 | 0.15 |
| req_battery_NCOV | 0.00 | 0.00 | 0.00 | 0.01 | 0.00 | 0.01 |
| req_battery_RFLU | 0.00 | 0.00 | 0.01 | 0.07 | 0.01 | 0.13 |
| req_battery_XCOV | 0.00 | 0.00 | 0.01 | 0.09 | 0.01 | 0.11 |

### Supplementary Note 3: Machine Learning

As described in the main paper, 12 datasets were used in the ML part of the analysis. (We also tested part of the pipeline using only 3 datasets, as described in Supplementary Note 7)

Figure S 5 shows how the class balance changes over the 12 datasets, ranging in the Pre-Covid dataset from 16% positive in T0, which is the moment of arrival for all patients (thus highly tilted toward the negative class) to balanced at T240, when approximately 50% of the patients that remain will be admitted, to tilted toward the positive class thereafter.

Although the datasets were imbalanced, the XGBoost documentation[2] advises against re-balancing the dataset using the scale_pos_weight parameter if (as in our case) you care about predicting the right probability – ie calibration is more important than classification. No adjustment was made for imbalanced datasets.

[2] https://xgboost.readthedocs.io/en/stable/tutorials/param_tuning.html

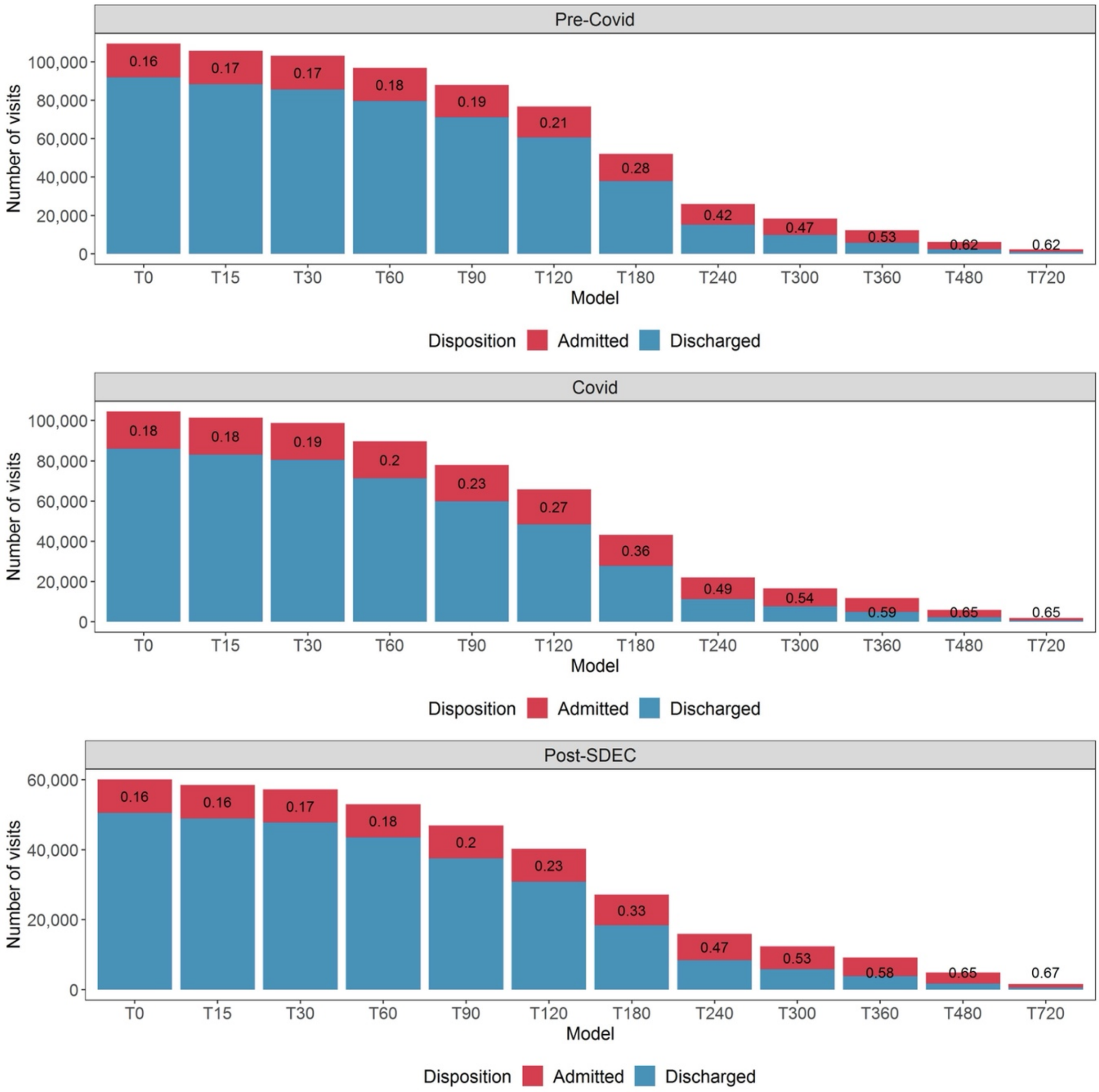


*Figure S 5: Class balance in each dataset*

We selected hyperparameters using a sequential tuning method, using mean log loss over the 10 cross-validation folds to decide which parameter was optimal. The tuning rounds for XGBoost were: number of estimators, tree-based hyperparameters (max depth and min child weight), samples and features (subsample and colsample by tree). The optimal parameter that improved on the previous round was selected for the next round. Early tuning attempts indicated that tuning gamma and the learning rate made no difference to performance. Since there was limited evidence of problems with over-fitting, gamma and learning rate were not included in tuning rounds. Figures S 6 to S 8 show model calibration across all three time periods. Relative to the Pre-Covid period, the Covid and Post-SDEC dataset are marginally weaker for the early models, and like the Pre-Covid period, their calibration deteriorates in the final model, where patients are long-staying and difficult to classify.

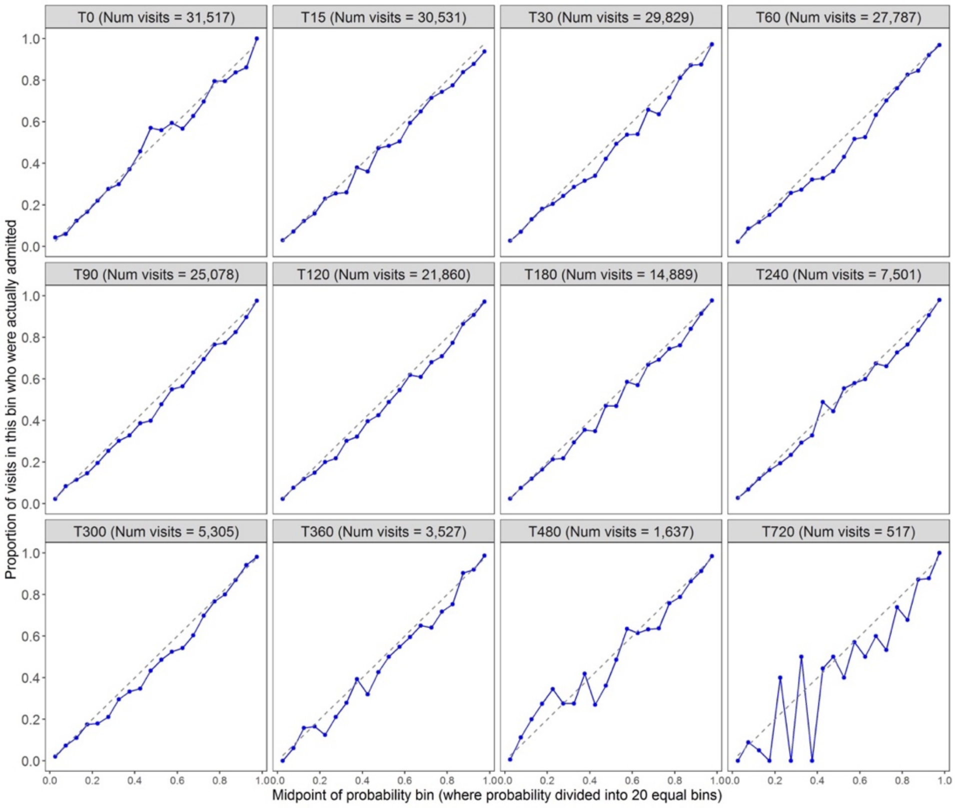


*Figure S 6: Calibration plots: Pre-Covid test set*

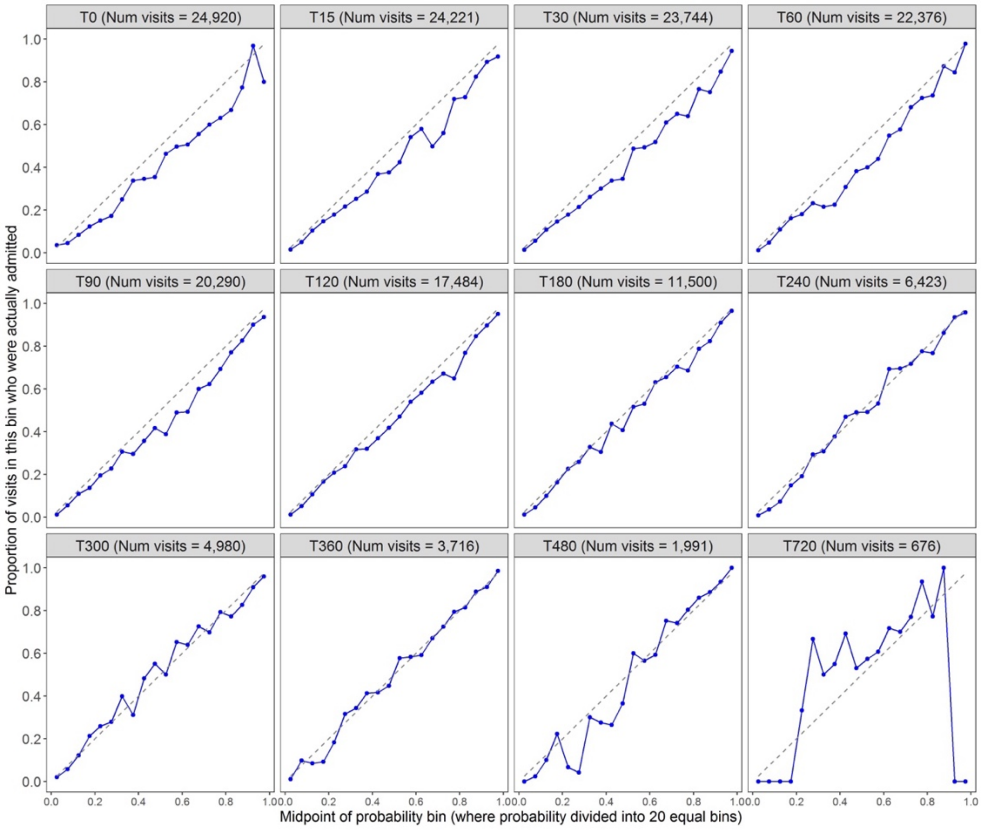


*Figure S 7: Calibration plots: Covid test set*

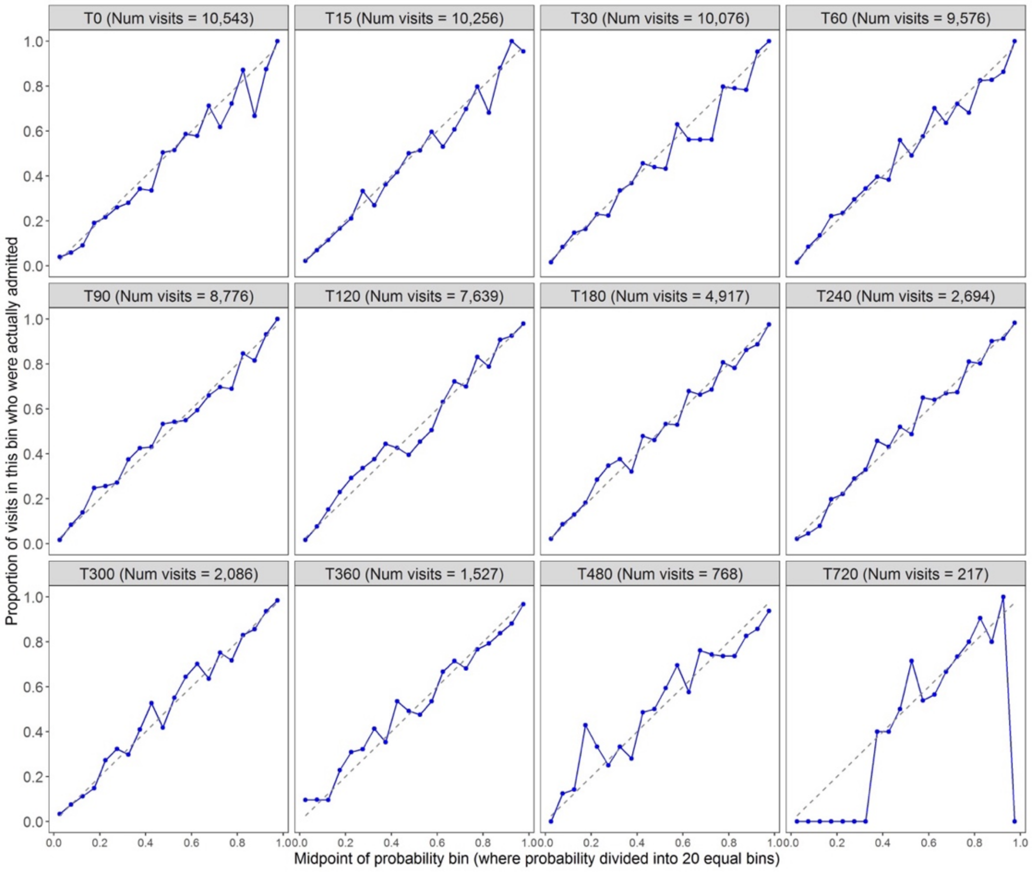


*Figure S 8: Calibration plots: Post-SDEC test set*

### Supplementary Note 4: Survival analysis

A dataset containing only admitted patients in the training set period was prepared. A Kaplan-Meier survival curve of 'survival times' (time from first arrival in an ED location to their departure to an inpatient ward) were calculated. A tail was added to the survival curve using an exponential distribution to allow for the possibility that an admitted patient in the future might remain in the ED for longer than any observations in the training set.

A Cox regression was fitted to the survival curve, with coefficients for factors known to be related to the time it would take patients to be admitted. These included: the patients' arrival window (between 06:00-12:00, 12:00-16:00, 16:00-22:00 and 22:00-06:00), the quarter of year (to reflect seasonal variations), whether patient presented on a weekday or weekend, and the number of other patients in various ED locations at the time each patient arrived. These were: num_Majors_Resus (the number of patients in Majors and Resus combined), num_SDEC (for post-SDEC dataset only, the number in SDEC), and num_elsewhere, the number in all other locations. Each variable was scaled prior to inclusion in the Cox regression. The coefficients are shown in Table S 4.

Each patient for whom prediction is made has an individually adjusted survival curve. If $X_i = (X_{i1}, \ldots, X_{ip})$ are the scaled values of the covariates for subject *i*, and $\beta = (\beta_1, \ldots, \beta_p)$ are the coefficients, and $\lambda_0(t)$ is the baseline hazard at time *t,* then the regression equation for patient *i* has the form

$$\lambda_0(t)\exp(X_i.\beta) \quad (1)$$

Factors associated with longer times to admission (from Table S 4), other things being equal, were a higher-than-average number of patients in Resus and Majors at the time a patient arrived, and a higher-than-average number of patients elsewhere. In the pre-Covid period, factors associated with shorter times to admission were arriving between 22:00 and 06:00 relative to other times of day, arriving at a weekend, and arriving in the third or fourth quarter of the year, rather than the first quarter, which was the reference category.

For a given patient $i$ in the ED, the analysis above yielded a survival function $Si(t)$, where $t = 0$ corresponds to that patient's arrival to the ED. At step 4 of the prediction pipeline, the conditional probability that patient $i$ is admitted by time $A+w$ *given* that they are to be admitted AND have not been admitted by time $A$ following their arrival was calculated as

$$(S_i(A) - S_i(A+w)) \,/\, S_i(A) \quad (2)$$

and multiplied by the probability of admission for patient $i$ estimated at Step 2 to give the probability for patient $i$ being admitted within the next interval $w$.

To evaluate the predicted survival curves, we used a method analogous to the QQ plots used elsewhere in this paper. At each report time *T* in the test set, for each patient in the ED, a survival curve for their probability of admission at a given moment *t* after their arrival was generated using the Cox regression equations, and converted to a cdf for time from *T* to admission. The observed time-to-event is each patient's time from *T* to their actual admission, and this observed time-to-event is associated with a quantile on their unique cdf. Figure S 10 shows the concordance between observations and the predicted distributions. From visual inspection, there is very good concurrence between the predicted distributions and observations for the Pre-Covid and Post-SDEC datasets. In the Covid dataset, more observed instances fall at the higher end of their distribution than would be expected by the predicted distributions. During the Covid test set period, patients were taking longer to be admitted than expected, with the result that fewer patients were admitted within the prediction window. See Supplementary Note 6 for discussion of the operational conditions leading to this slower processing of admissions during the Covid test set period.

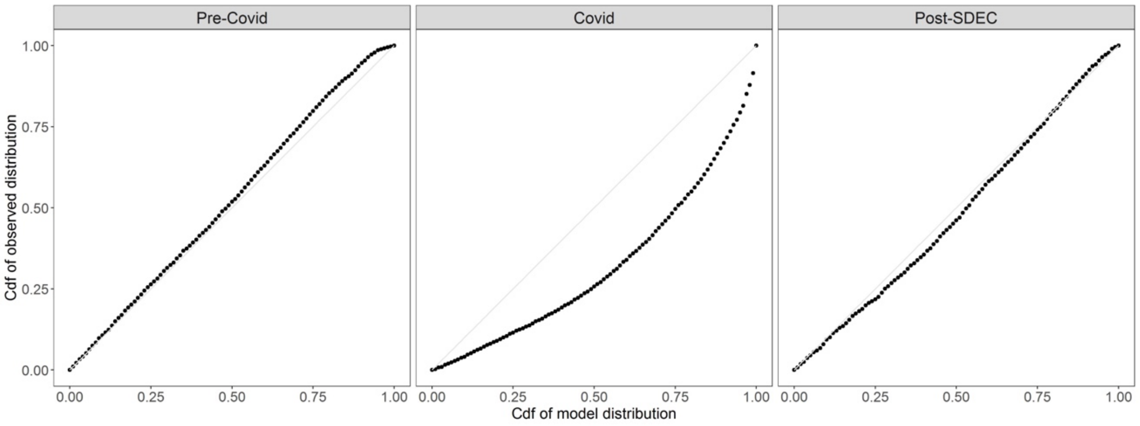


*Figure S 9: QQ plots evaluating survival curves used for predicting probability of admission within prediction windows, for Pre-Covid, Covid and Post-SDEC datasets*

*Table S 4: Regression coefficients for Cox regression equations*

| **Variables** | **Mean (SD) prior to scaling** | **Regression coefficients [95% CI]** |
|---|---|---|
| **Pre-Covid model** | | |
| quarter_3 | 0.46 (0.5) | 0.10 [ 0.08, 0.13] |
| quarter_4 | 0.25 (0.43) | 0.02 [ 0.00, 0.05] |
| weekend_1 | 0.25 (0.43) | 0.04 [ 0.02, 0.06] |
| arrival_time_06_12 | 0.2 (0.4) | 0.03 [ 0.00, 0.06] |
| arrival_time_12_16 | 0.26 (0.44) | 0.08 [ 0.05, 0.11] |
| arrival_time_16_22 | 0.36 (0.48) | 0.07 [ 0.04, 0.10] |
| num_MAJORS_RESUS_scaled | 23.1 (7.91) | -0.15 [-0.18, -0.13] |
| num_elsewhere_scaled | 33.26 (14.52) | -0.02 [-0.05, 0.01] |
| **Covid model** | | |
| quarter_2 | 0.19 (0.4) | 0.32 [ 0.29, 0.34] |
| quarter_3 | 0.27 (0.44) | 0.41 [ 0.39, 0.44] |
| quarter_4 | 0.29 (0.46) | 0.12 [ 0.10, 0.15] |
| weekend_1 | 0.24 (0.43) | 0.02 [ 0.00, 0.04] |
| arrival_time_06_12 | 0.19 (0.39) | 0.01 [-0.02, 0.03] |
| arrival_time_12_16 | 0.26 (0.44) | 0.12 [ 0.09, 0.14] |
| arrival_time_16_22 | 0.36 (0.48) | 0.12 [ 0.09, 0.15] |
| num_MAJORS_RESUS_scaled | 12.1 (5.81) | -0.12 [-0.14, -0.09] |
| num_elsewhere_scaled | 17.13 (11.06) | -0.16 [-0.19, -0.13] |
| **Post-SDEC model** | | |
| quarter_2 | 0.26 (0.44) | 0.12 [ 0.10, 0.13] |
| quarter_4 | 0.27 (0.44) | 0.21 [ 0.20, 0.23] |
| weekend_1 | 0.24 (0.43) | 0.05 [ 0.03, 0.06] |
| arrival_time_06_12 | 0.18 (0.39) | 0.01 [-0.01, 0.02] |
| arrival_time_12_16 | 0.27 (0.45) | -0.01 [-0.03, 0.00] |
| arrival_time_16_22 | 0.36 (0.48) | 0.12 [ 0.10, 0.13] |
| num_MAJORS_RESUS_scaled | 12.93 (5.67) | 0.13 [ 0.11, 0.15] |
| num_SDEC_scaled | 13.58 (6.85) | -0.26 [-0.27, -0.24] |
| num_elsewhere_scaled | 14.27 (8.26) | -0.12 [-0.14, -0.10] |

### Supplementary Note 5: Poisson regression

At each time of day (06:00, 12:00, 16:00 and 22:00) that predictions were to be made, a count of the number of patients who had not yet arrived, who were subsequently admitted within a prediction window of interest, was derived from each day in the training set. A Poisson regression was fitted to the count data, with coefficients for factors known to be related to the number of patients who presented at the ED. These included: the prediction time of day (06:00, 12:00, 16:00 and 22:00), quarter of year (to reflect seasonal variations), and weekday or weekend. The exponentiated coefficients are shown in Table S 5.

The estimated incident rate ratio (given by the exponentiated coefficient) for the Pre-Covid dataset for a report time of 12:00 was 4.24 (95% CI 3.36, 5.40) applying a four hour prediction window. Relative to the reference category of 06:00, the model estimates that 4.2 times the number of patients who have not yet arrived will be admitted within 4 hours of 12:00 than the number admitted within 4 hours of 06:00.

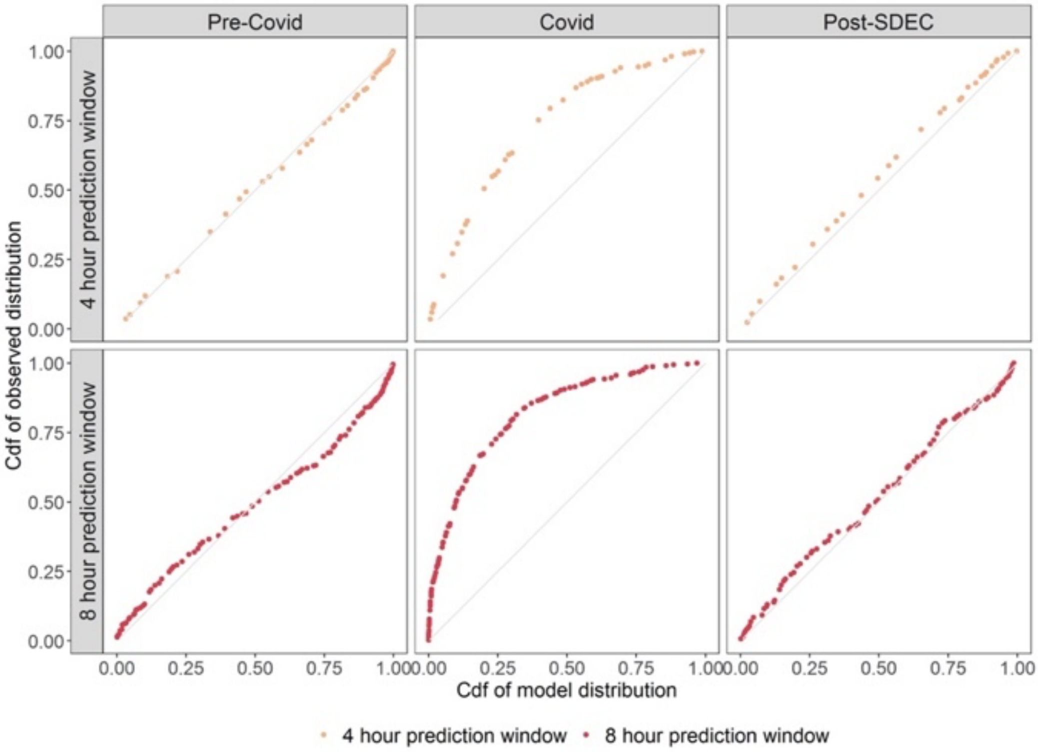


*Figure S 10: QQ plots evaluating Poisson distributions used for predicting numbers of patients yet-to-arrive for Pre-Covid, Covid and Post-SDEC datasets*

To evaluate the distributions derived from the Poisson regression, we used a method analogous to QQ plot but for discrete distributions. At each report time in the test set, a predicted distribution for the number of patients yet to arrive, who would be subsequently admitted within a prediction window, was generated from the Poisson equations. From visual inspection of the QQ plots shown in Figure S 1010, there is very good concurrence between the predicted distributions and observations for the Pre-Covid and Post-SDEC datasets. In the Covid dataset, more observed instances fall at the lower end of their distribution than would be expected by the predicted distributions. During the Covid test set period, patients were taking longer to be admitted than expected, with the result that fewer than expected yet-to-arrive patients were processed within the prediction window. See Supplementary Note 6 for further discussion of the operational conditions leading to this slower processing of admissions.

*Table S 5: Exponentiated coefficients from Poisson regression*

| **Variables** | **Exponentiated coefficients [95% CI]** | |
|---|---|---|
| | **4 hour prediction window** | **8 hour prediction window** |
| **Pre-Covid** | | |
| (Intercept) | 0.40 [0.31, 0.51] | 5.80 [5.38, 6.24] |
| quarter_2 | 0.93 [0.77, 1.11] | 0.95 [0.89, 1.02] |
| quarter_3 | 1.22 [1.04, 1.44] | 1.16 [1.09, 1.24] |
| weekend_1 | 0.93 [0.80, 1.07] | 0.92 [0.87, 0.97] |
| time_of_report_12 | 4.24 [3.36, 5.40] | 1.79 [1.67, 1.92] |
| time_of_report_16 | 3.89 [3.08, 4.97] | 1.58 [1.47, 1.70] |
| time_of_report_22 | 1.96 [1.52, 2.56] | 0.68 [0.62, 0.74] |
| **Covid** | | |
| (Intercept) | 0.22 [0.17, 0.28] | 2.29 [1.82, 2.89] |
| quarter_1 | 0.58 [0.47, 0.72] | 3.66 [3.42, 3.91] |
| quarter_2 | 1.57 [1.32, 1.86] | 0.65 [0.61, 0.70] |
| quarter_3 | 2.21 [1.89, 2.60] | 1.08 [1.02, 1.15] |
| weekend_1 | 0.87 [0.76, 0.99] | 1.50 [1.42, 1.59] |
| time_of_report_12 | 4.00 [3.24, 4.98] | 0.83 [0.79, 0.87] |
| time_of_report_16 | 3.99 [3.23, 4.97] | 2.05 [1.92, 2.19] |
| time_of_report_22 | 2.29 [1.82, 2.89] | 2.04 [1.91, 2.18] |
| **Post-SDEC** | | |
| (Intercept) | 0.11 [0.06, 0.17] | 1.82 [1.60, 2.08] |
| quarter_1 | 1.00 [0.74, 1.36] | 0.94 [0.86, 1.03] |
| quarter_2 | 0.91 [0.64, 1.30] | 0.87 [0.78, 0.97] |
| weekend_1 | 1.21 [0.92, 1.57] | 0.90 [0.83, 0.98] |
| time_of_report_12 | 3.95 [2.47, 6.63] | 3.16 [2.78, 3.60] |
| time_of_report_16 | 4.75 [3.00, 7.91] | 3.32 [2.92, 3.78] |
| time_of_report_22 | 2.70 [1.64, 4.62] | 1.37 [1.18, 1.59] |

### Supplementary Note 6: Comparing results across the Pre-Covid, Covid and Post-SDEC datasets

Figure S 11 shows the feature importance used by each ML model in each of the Pre-Covid, Covid and Post-SDEC datasets. (See Table S 2 for a description of each feature.) Patterns of feature importance are consistent across each dataset, confirming that the ML models are relatively unaffected by changes brought about by the pandemic. There are a few differences. Among the **location features** and earlier in the stay, being in a Waiting location is more important Pre-Covid. During that period, the ED was much busier, so waiting is indicative of lower likelihood of admission. During Covid and Post-SDEC, visiting the UTC (for minor illness and treatment) appears to substitute for waiting as an indicator of lower acuity.

Among the **observation features**, in the Covid dataset, features relating to oxygen delivery are important in models trained on visits lasting longer than 60 minutes; use of delivery devices is sparsely recorded in ED in general, so these indicators possibly reflect care for patients unwell with Covid who were later admitted. During Covid and Post-SDEC, the NEWS and Manchester triage scores are more important, again perhaps compensating for the relatively lower utility of waiting as an indicator of acuity.

Among the **pathology features,** orders of specific pathology tests and their latest values were more important in the Covid dataset than Pre-Covid, including NCOV and XVOC which are lab tests for the Covid disease. These Covid tests were not available early in the Covid training set; their first use was June 2020. This may explain why they have stronger importance in the Post-SDEC dataset when they were available throughout in the training set period. pCO2 is indicative of respiratory issues, possibly explaining its relatively higher importance in the Covid and Post-SDEC datasets.

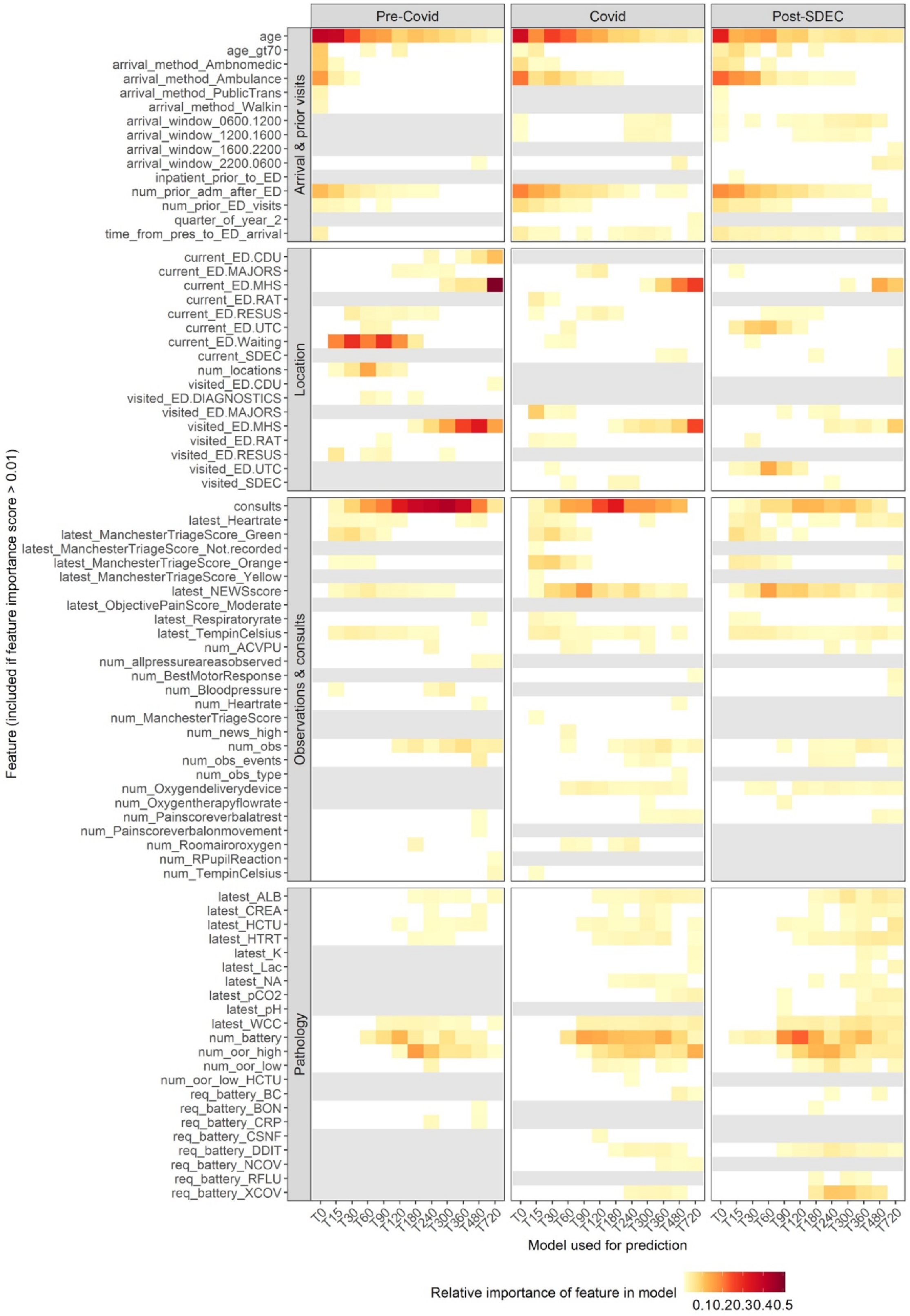


*Figure S 11: Feature importance and performance for each model on test set*

The colour intensity reflects the relative importance of each feature within each model. A feature is included if it had a raw importance of more than 0.01 in any model. The grey areas indicate features that were only important in either the pre-Covid or Covid periods; where greyed out they either never achieved an importance of greater than 0.1, or (as for example the CDU location which was not used during Covid, and the NCOV and XCOV tests which were not used pre-Covid) were not relevant at all in the period.

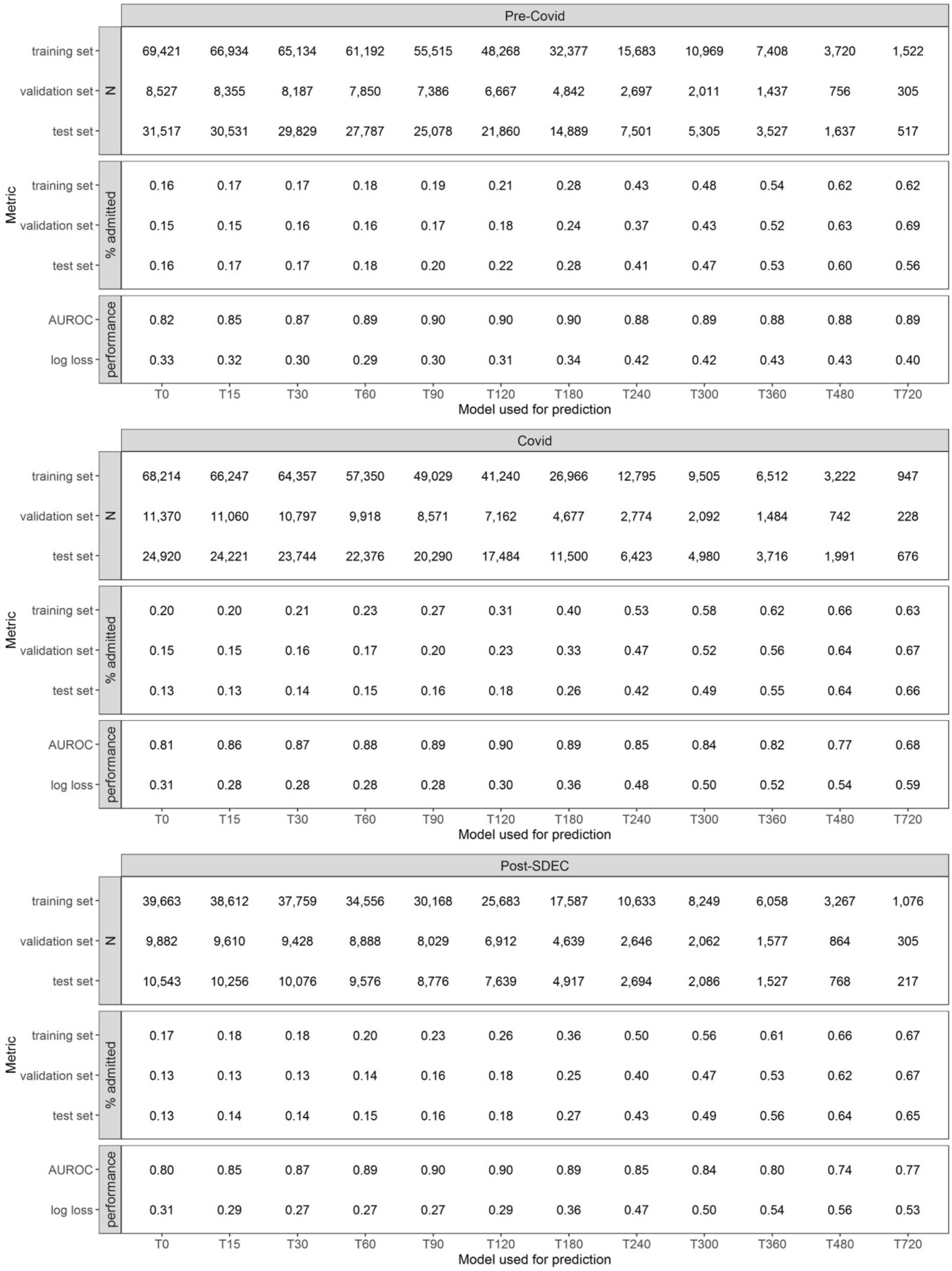


*Figure S 12: Performance for each model on test sets of Pre-Covid, Covid and Post-SDEC datasets*

Figure S 12 shows the performance of each model. As for Pre-Covid, the Covid and Post-SDEC models achieved their lowest log loss when presented with patients with elapsed times of between 30 and 120 minutes, and the best AUROC between 90 and 120 minutes. In the Covid and post-SDEC datasets, model fit (denoted by AUROC and log loss) deteriorates more than Pre-Covid in the later models, possibly because the summer of 2021 (the period of their test sets) saw greater heterogeneity in the pathology or the management of longer-staying patients.

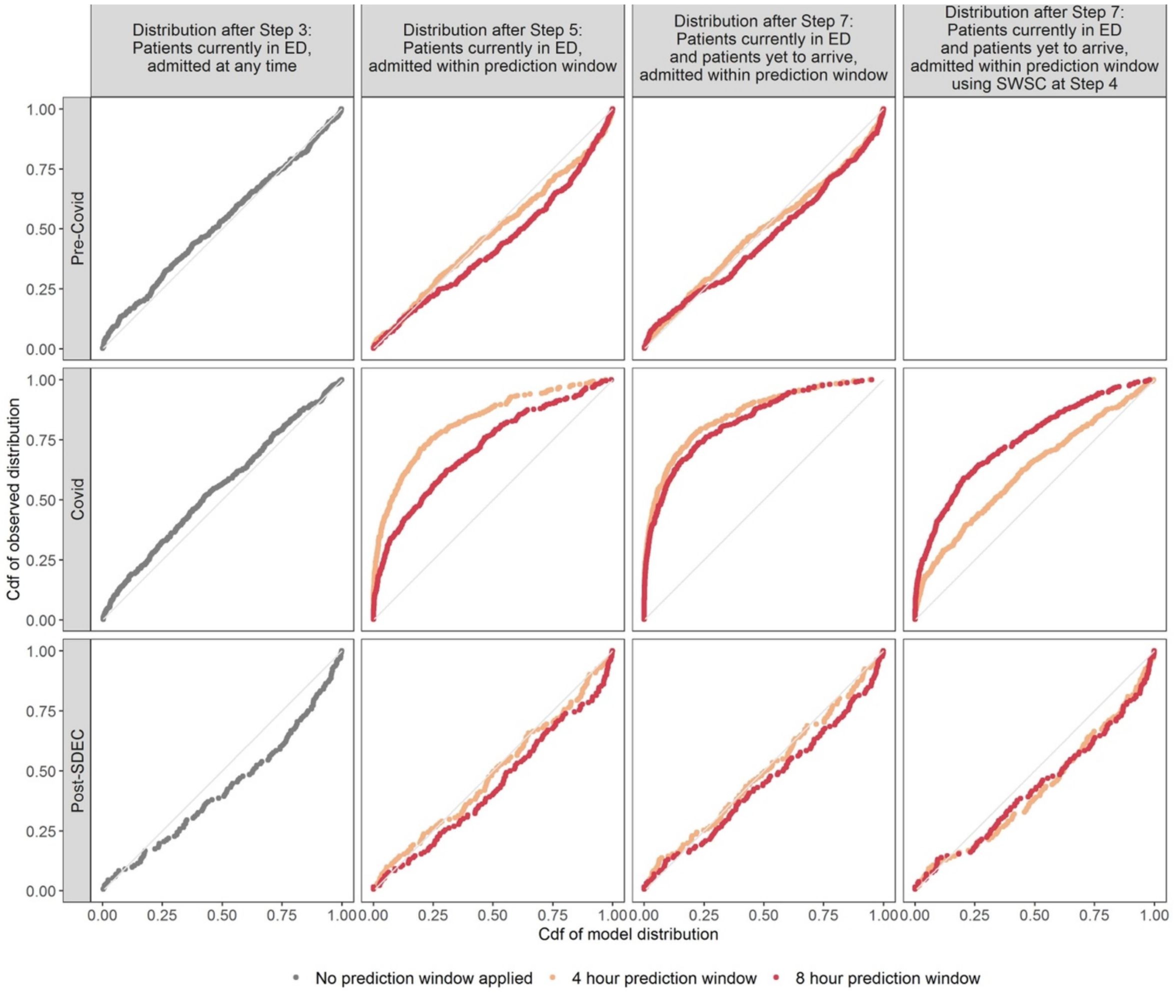


*Figure S 13: QQ plots evaluating distribution of models predicting number of beds needed for Pre-Covid, Covid and Post-SDEC datasets*

As noted in the main paper, there is good concurrence between the predicted distributions and observations in the **pre-Covid** period (top row Figure S 11). When no prediction window is applied, the **Covid** aggregate predictions are also well-balanced (middle row, first column). With a prediction window, the Covid data shows a wide deviation (second column, second row), with more observed instances falling at the lower end of their distribution than would be expected by the predicted distributions. During the Covid period, patients were taking longer to be admitted than expected. The large deviation at this step can be explained by changing operational conditions in the ED. The training set for the Covid dataset was one year from 19 March 2020. During that 12-month period, EDs were relatively empty and hospitals were anxious to minimise risk of infection so times spent in the ED before admission were relatively short. In the latter part of the year, numbers presenting had begun to increase and they were consistently higher during the test set period from May to July 2021, when EDs had returned to levels of busy-ness more like the pre-Covid period. This difference between training and test set periods is illustrated in the middle column Figure S 14. This difference between training and test set did not apply in the Post-SDEC test set.

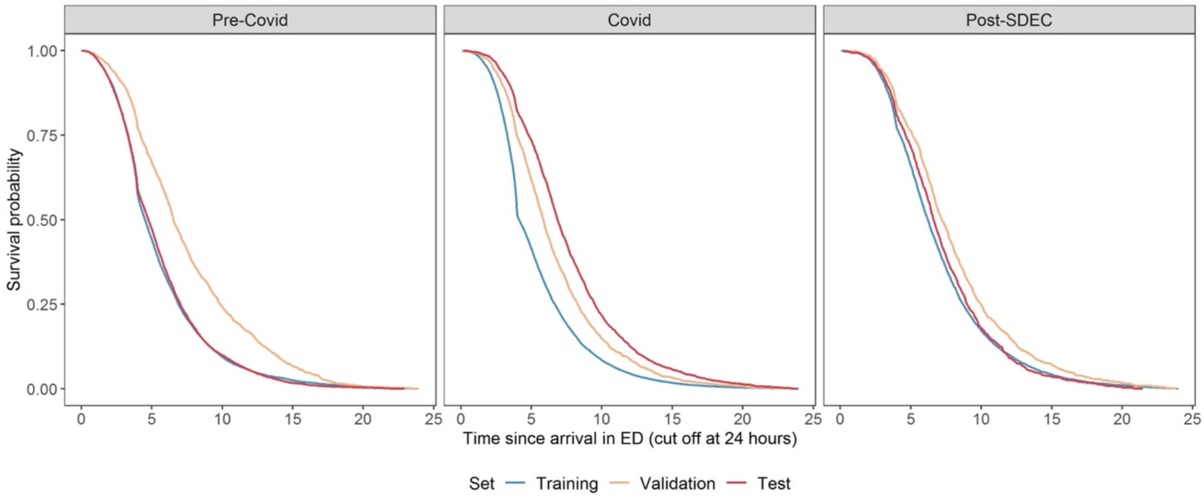


*Figure S 14: Survival curves for Pre-Covid, Covid and Post-SDEC training, validation and test sets*
Note that survival refers here to each patient's probability of still being in the ED at the elapsed time after arrival

Figure S 15 shows the performance of each against the benchmark (see main paper for explanation of the benchmark). Pre-Covid, the shaded band is mainly above the x axis; the model underestimated the number admitted in the prediction window. During the Covid period, when using the training set survival curve (second column in Figure S 15), the shaded band is mainly below the x axis. The model overestimated the number admitted in the prediction window. The six week benchmark outperformed the model during the Covid period (MAE for model of 6.5, compared with 5.0 for the benchmark). However, during the post-SDEC period (fourth column of Figure S 15), the model outperformed the benchmark (MAE for model of 3.3 compared with 5.5 for the benchmark).

In the third and fifth columns of Figure S 15, the survival curve and Cox regression coefficients used for Step 4 have been derived using a sliding window of the last six weeks' data on times to admission. This post-hoc adjustment shows that final predictions on the test set using a sliding window survival curve (SWSC) are improved, relative to using the training set survival curve and coefficients. With this, the Covid dataset outperforms the benchmark (MAE of 4.2 relative to 4.9 for the benchmark). Using the SWSC did not benefit the post-SDEC test set, because its training and test set survival curves are very similar as noted above.

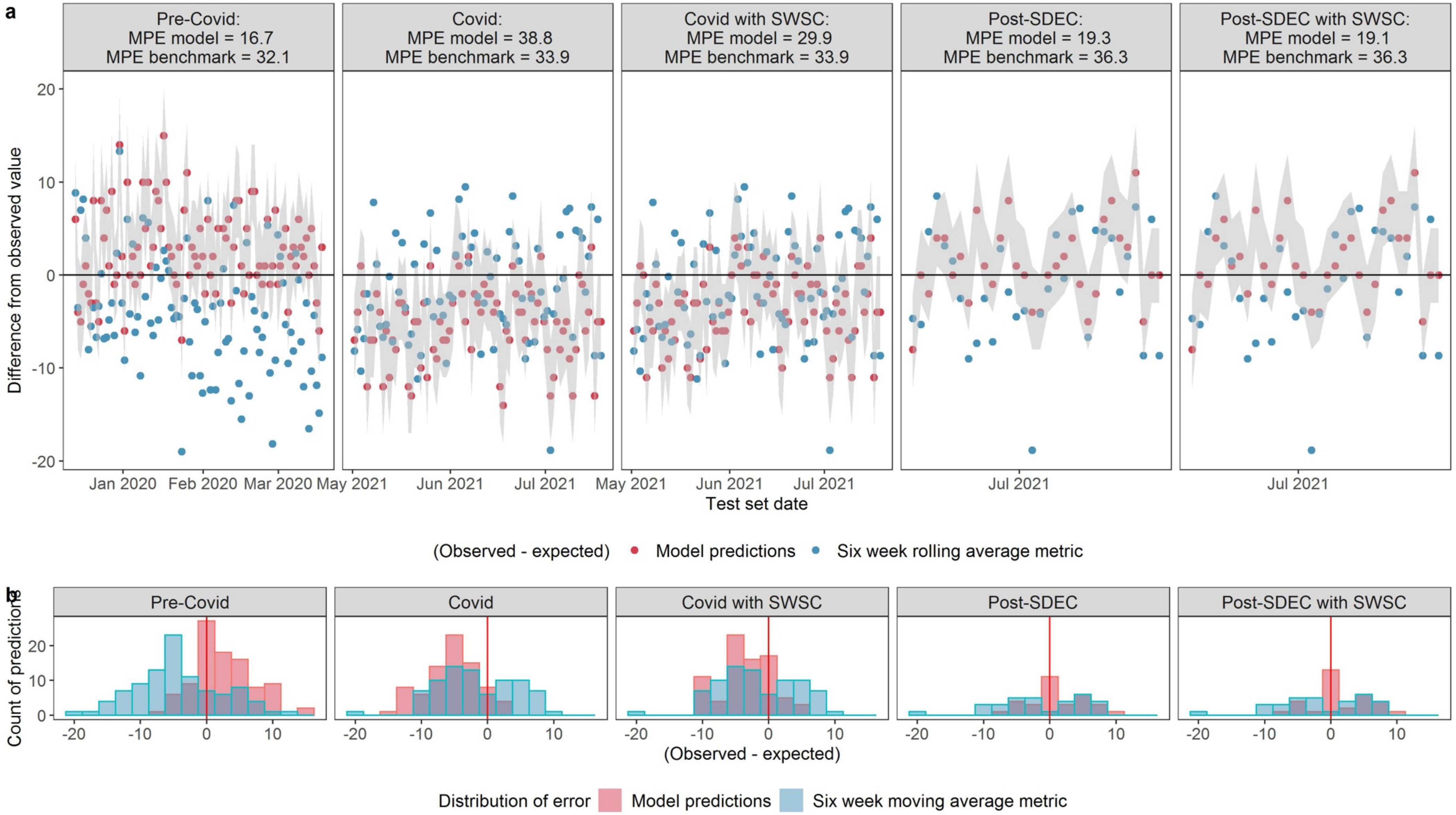


*Figure S 15: Comparing model predictions with six week rolling average benchmark for number of admissions 8 hours after 16:00, including patients who yet to arrive, for Pre-Covid, Covid and Post-SDEC datasets*

In the third and fifth columns a sliding window survival curve (SWSC) has been applied. **a** shows the difference between the observed number of admissions and the expected value from the probability distribution for the model predictions (the red dots) and between the observed number of admissions and expected value from the benchmark (the blue dots). Where the expected value equals the observed value, the dots fall on the x axis (y = 0). The grey shaded band represents the range of probability between the $10^{th}$ and the $90^{th}$ centile of the cumulative probability distribution of the model. **b** shows the distribution of errors (difference between expected and observed). The model predictions, although offset from 0, were in a tighter range than the benchmark in each case.

### Supplementary Note 7: Comparing 3 and 12 models

Our a priori reasoning for training 12 temporally framed models in Steps 1 and 2 of our pipeline was that a larger number of models could exploit evolving data completeness during a patient's visit to ED, with models able to infer different signals about missingness of data. The use of more models also made it possible to interrogate how other feature importance changed with a patient's elapsed time in the ED.

A previous paper looking at predicting ED admissions used 3 temporally framed models[3]. To assess the utility of additional models, we conducted a post-hoc analysis using only 3 of our 12 models to make predictions, using the T0 model to make predictions on patients with elapsed times in ED of less than 90 minutes, the T90 model for elapsed times less than 240 min and T240 for the remainder.

In Figure S 16, we show the calibration plots obtained for the Pre-Covid period for each of the 12 models compared to the calibration plot achieved among the same cohort using the T0, T90 or T240 model as appropriate. The tiles in the figure relating to T0, T90 and T240 are, by definition, identical. The tiles relating to other models show that using 3 models rather than 12 gives worse calibration, most notably for predictions at elapsed times of 15-89 minutes. Calibration fluctuates widely for the T480 and T720 models and when using the T240 model for predictions made at elapsed times > 480 minutes.

Figure S 17 compares the QQ plots obtained after Step 3 of our pipeline using all 12 or just the 3 patient-level models for each of the Pre-Covid, Covid and Post-SDEC periods. The performance is worse when using just 3 models for Pre-Covid and Post-SDEC and similar for the Covid period.

[3] Barak-Corren, Y., Israelit, S. H. & Reis, B. Y. Progressive prediction of hospitalisation in the emergency department: Uncovering hidden patterns to improve patient flow. Emerg. Med. J. 34, 308–314 (2017).

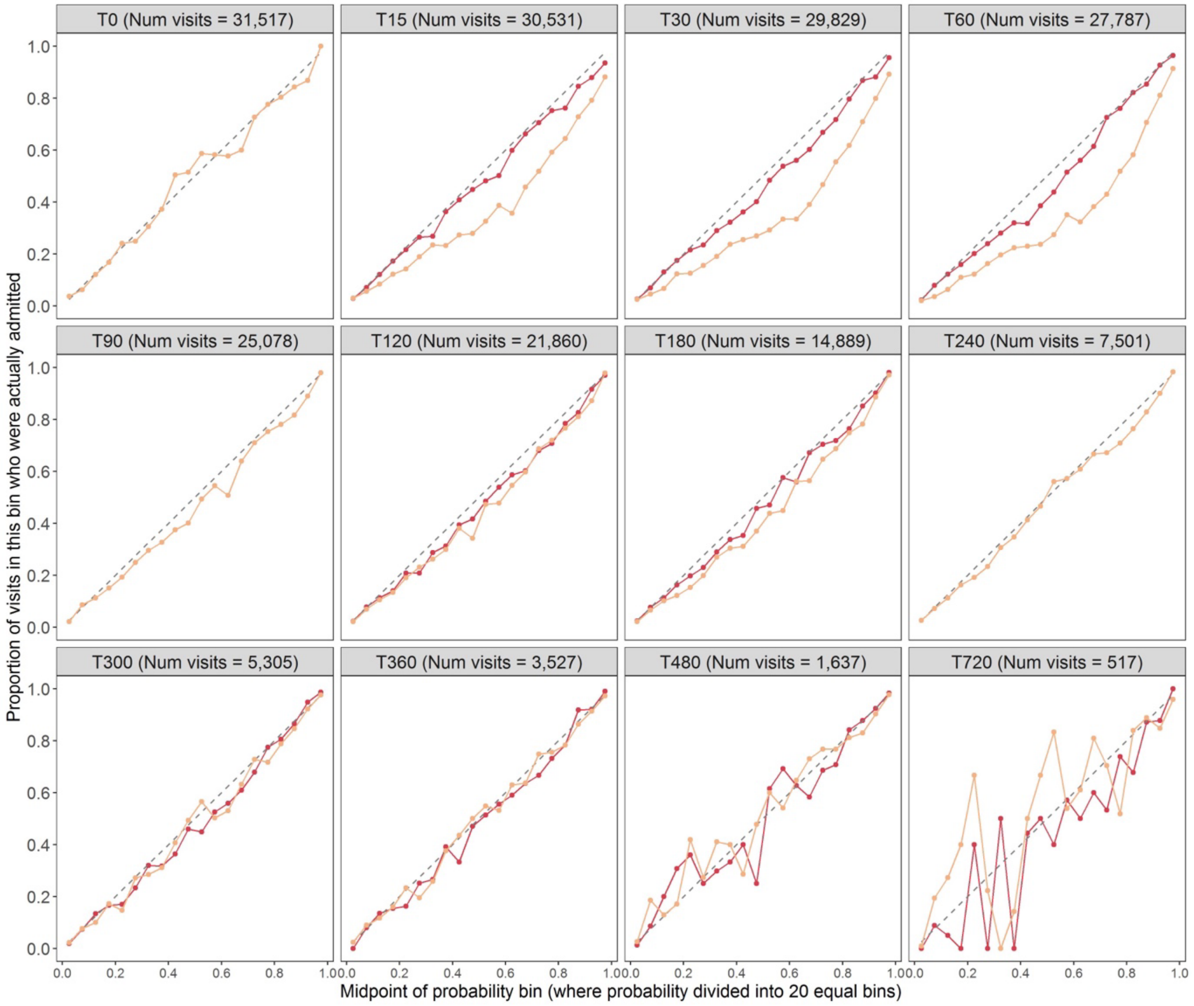


*Figure S 16: Calibration plots when using 3 models at Step 2*

The darker, red lines show the calibration plot obtained for the 12 models T0-T720 for the Pre-Covid period. The lighter, yellow lines show the calibration plot obtained applying the T0 model for elapsed times up to 89 minutes, the T90 model for elapsed times of 90- 239 minutes and the T240 model for elapsed times of 240 minutes or longer.

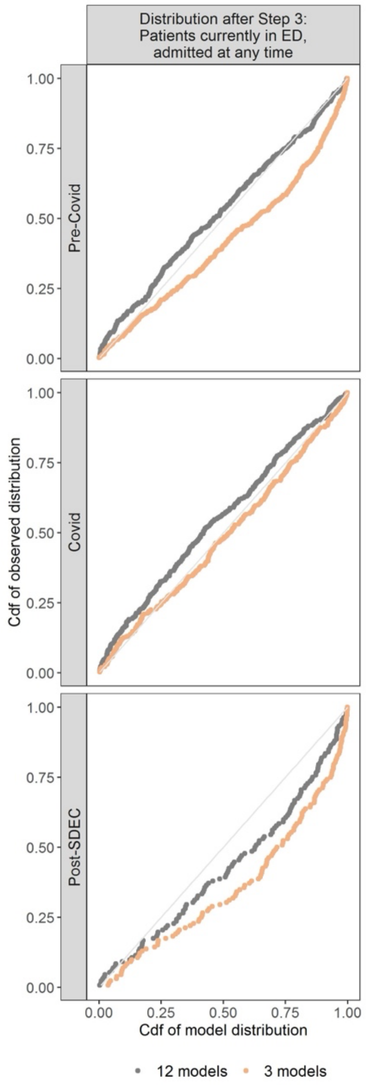


*Figure S 17: Comparing 3 and 12 models at Step 3*
QQ plots showing the performance of the predictions of aggregate admission among all patients in ED at each report time in the test set of each model period. This column of charts is the equivalent to the left hand column of Figure S 13, showing the same grey line for 12 patient-level models, and an additional yellow line for predictions derived from 3 patient-level models only.

.

### Supplementary Note 8: Examining alternatives to XGBoost

We conducted additional analyses at the ML stage (Steps 1 and 2 of our pipeline) to see if other algorithms showed comparable performance and similar important variables. We compared Random Forest (RF) and Logistic Regression (LR) with XGBoost across the 12 models used in Steps 1 and 2 on the Pre-Covid dataset. Each started with the same feature set as XGBoost (range of 209 to 294 features); missing values were imputed at their median then any features with a uniform value after imputation were removed. This left a range of 156 to 274 features for Random Forest.

As there is an implementation cost associated with making variables available for analysis, we investigated the smallest subset of features sufficient for making a good prediction, and the performance implications of reducing the model to this subset. For this, we implemented Logistic Regression with Lasso regularisation using the *Lasso and Elastic-Net Regularized Generalized Linear Models* algorithm [4] and the associated R package GLMNET [5]. The algorithm aims to identity the optimal value for the regularisation penalty, referred to as lambda, by fitting models over a vector of penalty values and using cross-validation to determine the mean log loss at each value of lambda. Lasso regularisation will find a sparser solution, ie a regression equation with more coefficients set to zero, the greater the penalty value.

The Logistic Regression was given the same feature set as for Random Forest, with missing values imputed at the median. We used GLMNET on the Pre-Covid training set to derive, for each of our 12 models T0 to T720, the best performing LR model and 3 LR models that were more heavily penalised for having a larger number of coefficients (by constraining GLMNET to solutions with a mean loss 2, 3 or 4 standard errors away from that of the lambda.min model, referred to as the lambda.2se, lambda.3se and lambda.4se models respectively). These regression equations were applied to the test set to generate the performance scores show in Figure 4 in the main paper. Figure S 18 shows the number of non-zero coefficients in each LR model, illustrating how many non-zero coefficients remain as regularisation increases.

[4] For details of the algorithm, please see https://hastie.su.domains/Papers/glmnet.pdf and https://www.jstatsoft.org/article/view/v033i01
[5] See page 5 of https://cran.r-project.org/web/packages/glmnet/vignettes/glmnet.pdf

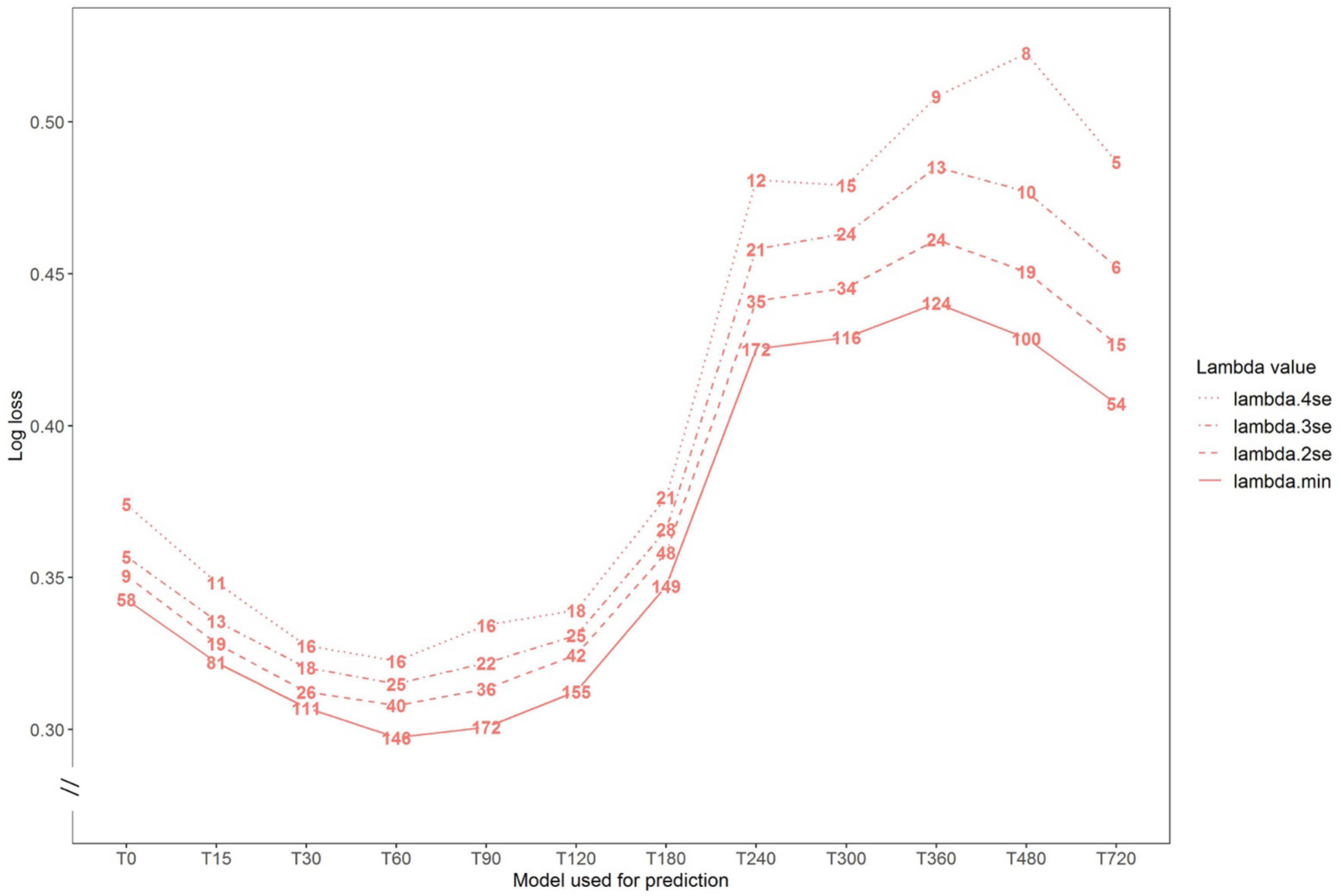


*Figure S 18: Number of non-zero coefficients (excluding the intercept) in each regularised LR model*

Below, we include figures showing the importance scores returned by the Random Forest algorithm (Figure S 19), and the values of non-zero coefficients in each of the regularised LR equations (Figure S 20 to Figure S 23).

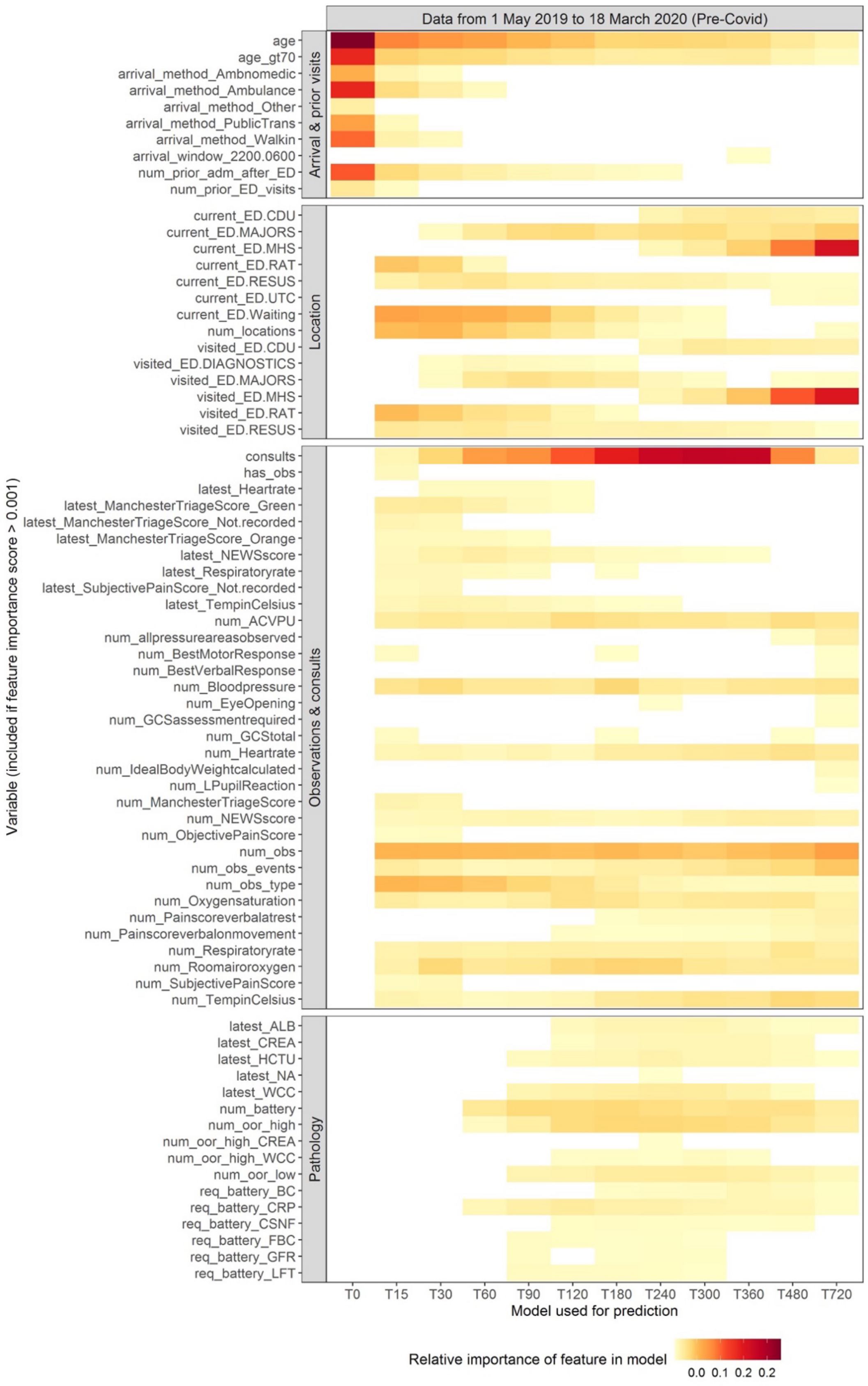


*Figure S 19: Feature importance for each model on test set using Random Forest*
The colour intensity reflects the relative importance of each feature within each model. For simplicity of presentation, a feature is excluded from the figure if it had a raw importance of less than 0.001 in all models.

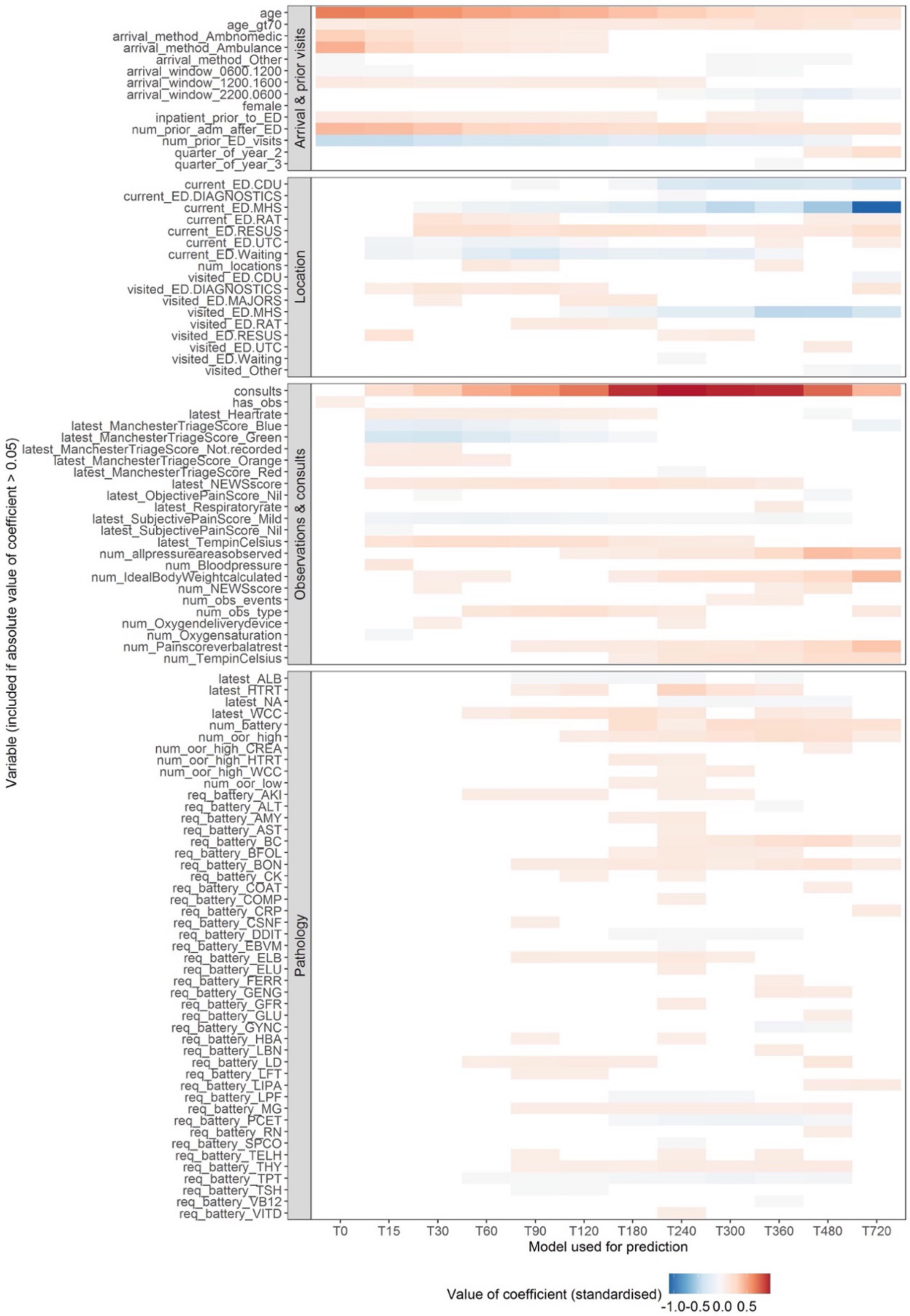


*Figure S 20: Logistic regression with optimal penalty value, lambda.min: standardised regression coefficients with absolute value > 0.05*

The colour reflects the size and sign of each coefficient, where negative values are blue and positive values are red. Note that, for the purpose of visual presentation, coefficients have only been included if their absolute value is greater than 0.05. Therefore, for any model, the number of coloured cells in this table will not match the number of variables marked in Figure S 18 above. For example, T0 has 59 non-zero coefficients but only 11 of them are > 0.05 in absolute value.

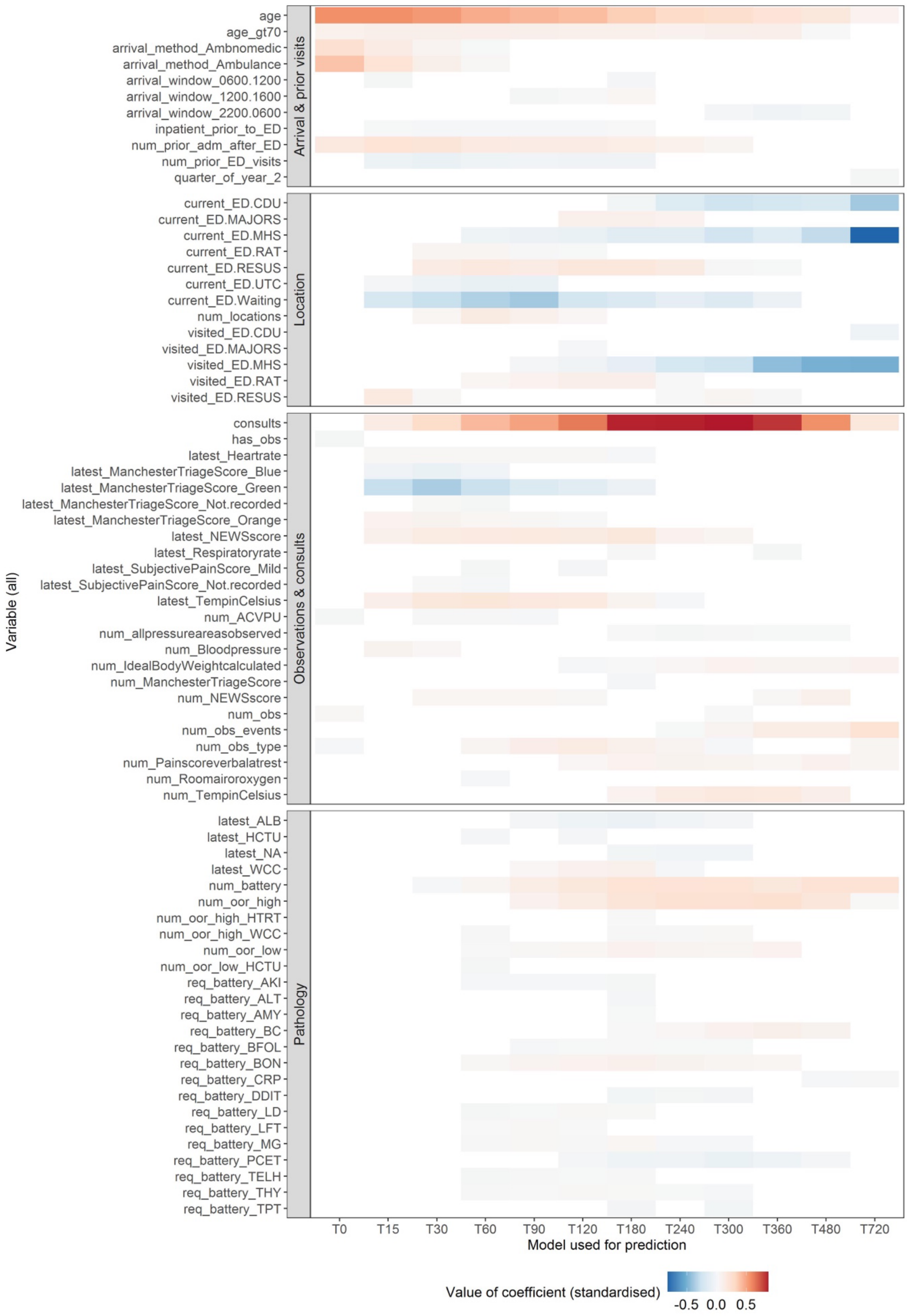


*Figure S 21: Logistic regression with penalty value lambda.2se: standardised regression coefficients (all)*
The colour reflects the size and sign of each coefficient, where negative values are blue and positive values are red.

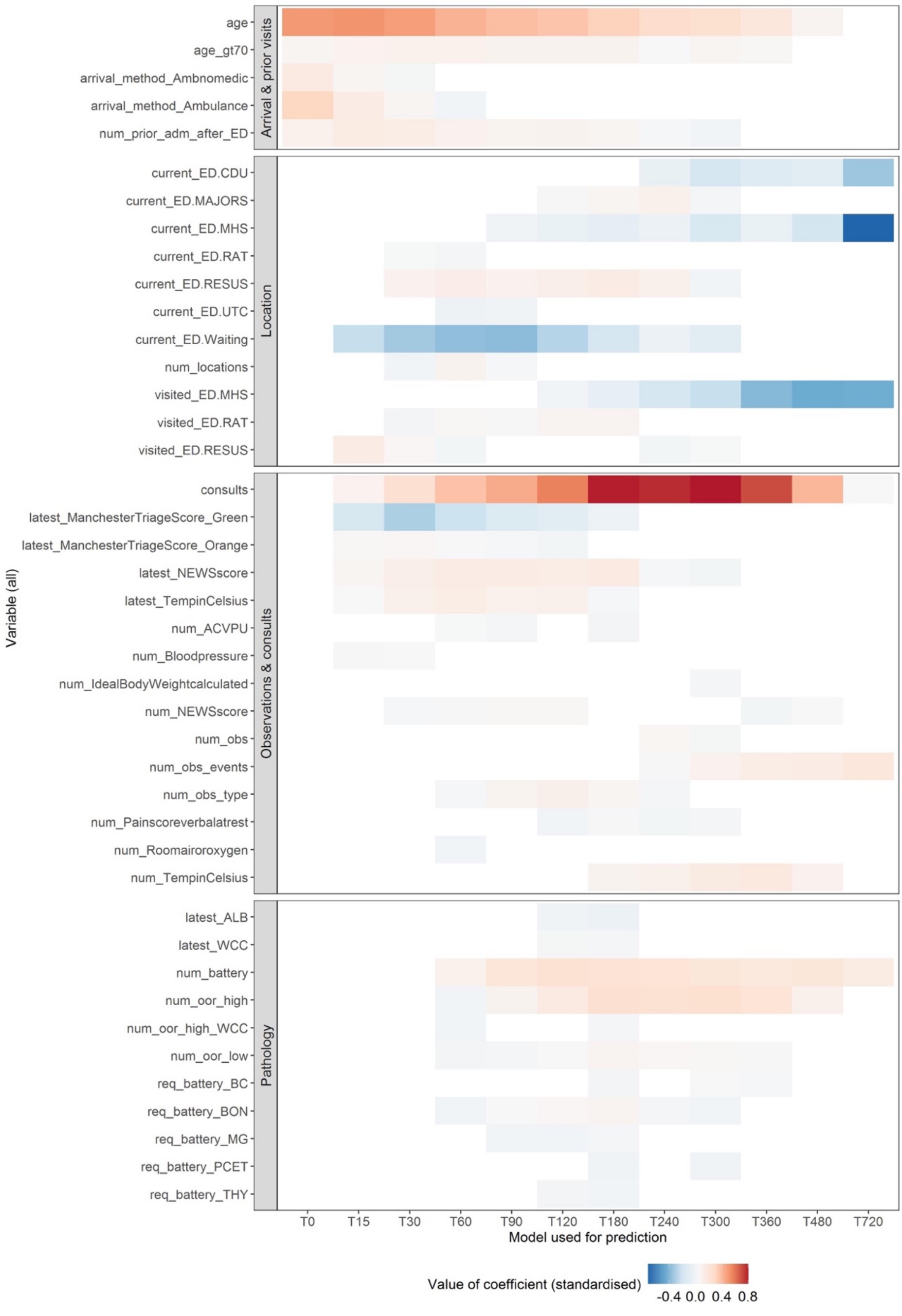


*Figure S 22: Logistic regression with penalty value lambda.3se: standardised regression coefficients (all)*
The colour reflects the size and sign of each coefficient, where negative values are blue and positive values are red.

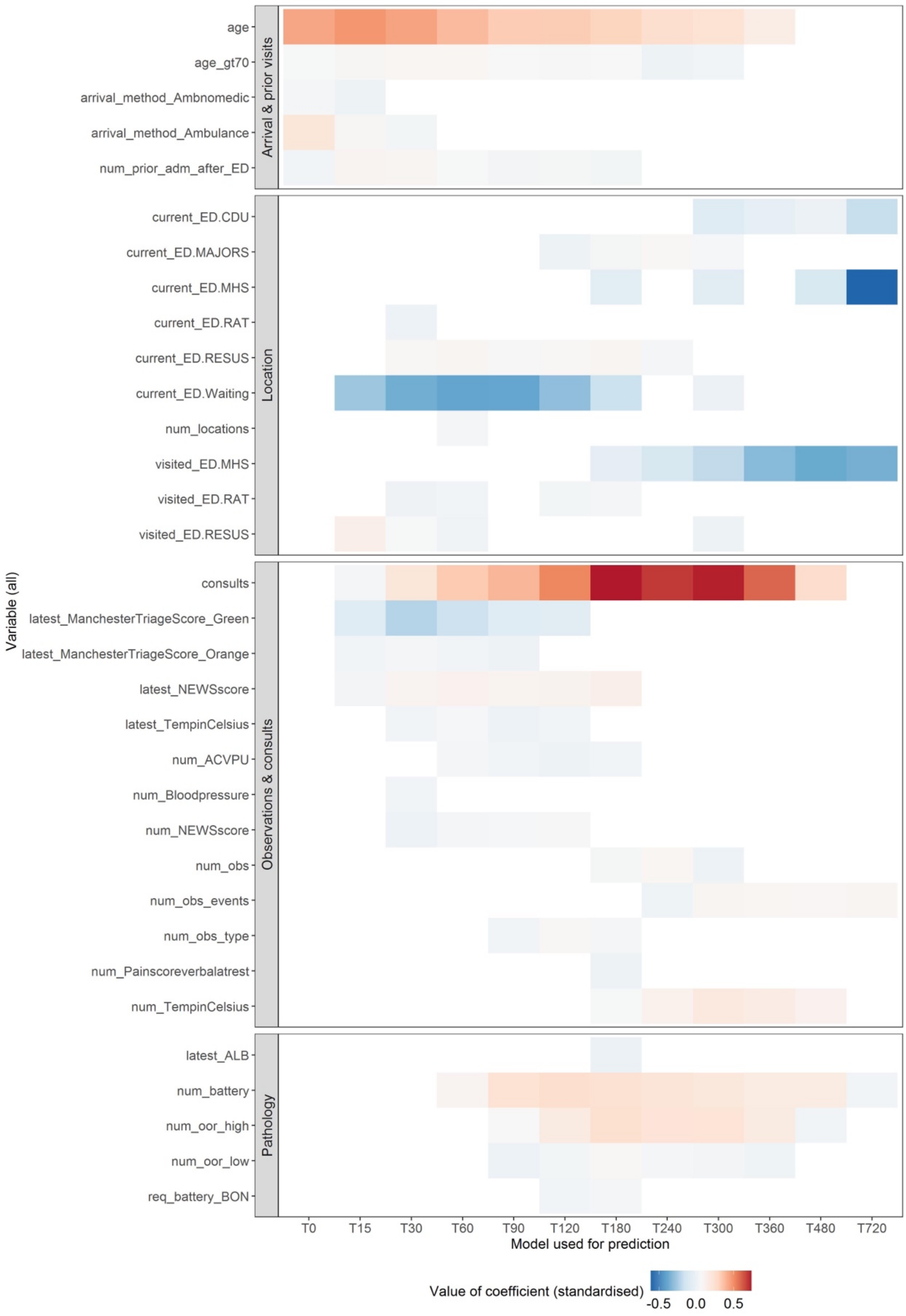


*Figure S 23: Logistic regression with penalty value lambda.4se: standardised regression coefficients (all)*
The colour reflects the size and sign of each coefficient, where negative values are blue and positive values are red.

### Supplementary Note 9: Details of the application in use

The existing workflow is that four times per day, one of the hospital's bed managers manually prepares and emails an Excel spreadsheet reporting on current and predicted hospital capacity between the current time and midnight, as well as ED performance and OPEL status[6]. These reports are intended to stimulate discharge activity when required. As noted in the main paper, these predictions work on a midnight-to-midnight basis. Predicted admissions (a single number) in each midnight-to-midnight period is derived from a reference day, which is either the previous day or the same day last week.

Our application runs in a Docker container on a security-enhanced RedHat Linux machine within the hospital network. It draws data from a PostgresSQL database that is updated in real-time from the EHR, generates predictions from saved models, and formats these into an email which is sent to two members of the bed management team. The application is subject to the clinical risk management processes enshrined in the DCB0160 standard[7] and to NHSx Digital Technology Acceptance Criteria[8]. At the time of writing, the application is at the user acceptance stage, so has not been fully integrated into the whole team's workflow. Screen shots of an example email are shown in Figure S 24.

The benefits of the application, which are difficult to isolate and evaluate, are assumed to flow from enriching the information available to bed managers for predicting bed capacity requirements. For example, at 22:00, the current midnight-to-midnight projections used give no insight into overnight demand. In contrast, in the example below, generated at 22:00, our application gives an indication of how many beds will be needed during the night for patients currently in the ED, enabling planning to free up capacity in the morning.

---

[6] Operational Pressures Escalation Levels https://www.england.nhs.uk/publication/operational-pressures-escalation-levels-framework/

[7] https://digital.nhs.uk/data-and-information/information-standards/information-standards-and-data-collections-including-extractions/publications-and-notifications/standards-and-collections/dcb0160-clinical-risk-management-its-application-in-the-deployment-and-use-of-health-it-systems

[8] https://www.nhsx.nhs.uk/key-tools-and-info/digital-technology-assessment-criteria-dtac/

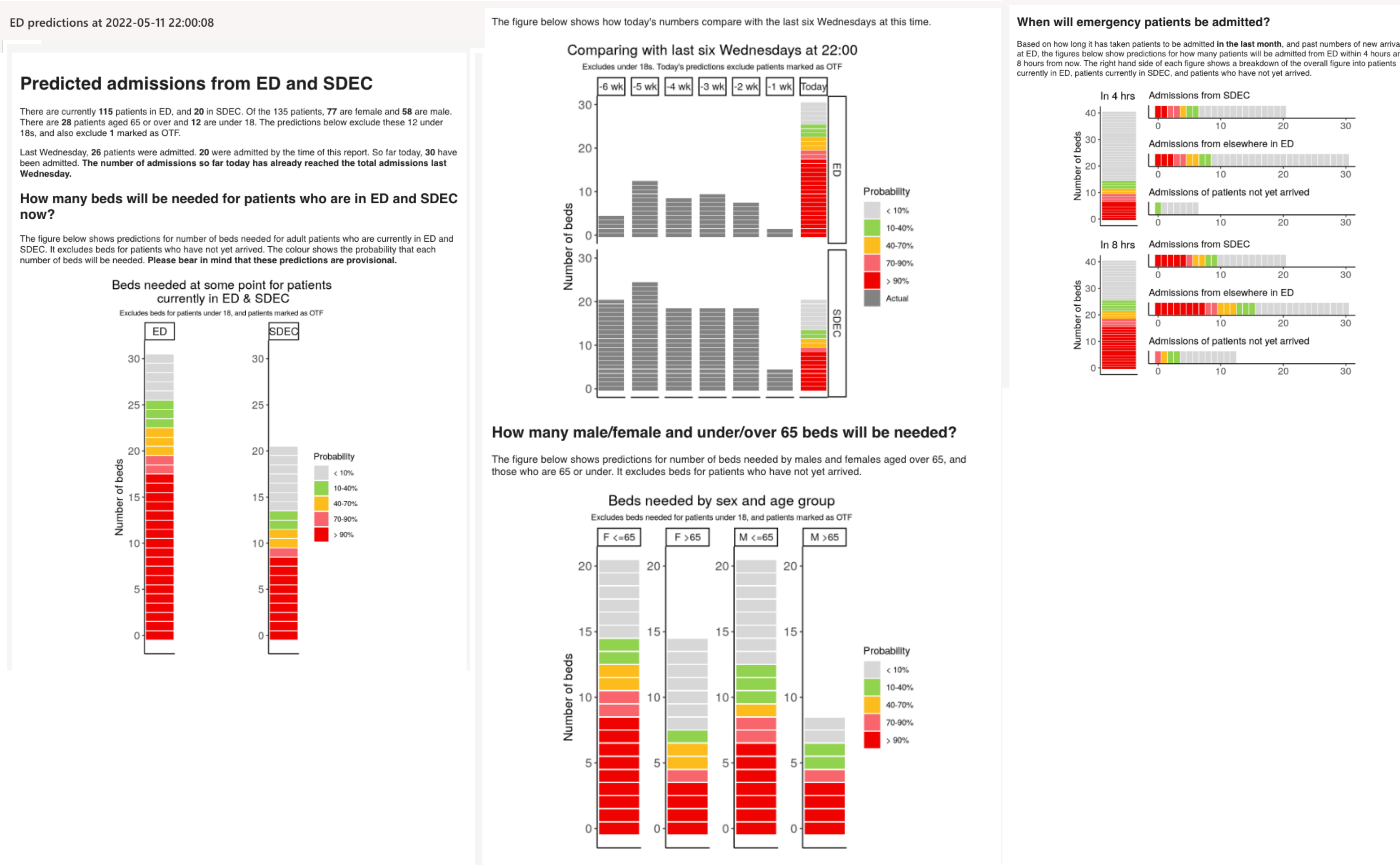


*Figure S 24: Screenshots of the application in use*

### Supplementary Note 10: TRIPOD Checklist for Prediction Model Development and Validation

Transparent reporting of a multivariable prediction model for individual prognosis or diagnosis (TRIPOD) is a checklist for reporting of studies developing, validating, or updating a prediction model, whether for diagnostic or prognostic purposes. [9]

| Section/Topic | Item | | Checklist Item | Page |
|---|---|---|---|---|
| **Title and abstract** | | | | |
| Title | 1 | D;V | Identify the study as developing and/or validating a multivariable prediction model, the target population, and the outcome to be predicted. | 1 |
| Abstract | 2 | D;V | Provide a summary of objectives, study design, setting, participants, sample size, predictors, outcome, statistical analysis, results, and conclusions. | 2 |
| **Introduction** | | | | |
| Background and objectives | 3a | D;V | Explain the medical context (including whether diagnostic or prognostic) and rationale for developing or validating the multivariable prediction model, including references to existing models. | 3-6 |
| | 3b | D;V | Specify the objectives, including whether the study describes the development or validation of the model or both. | 5 |
| **Methods** | | | | |
| Source of data | 4a | D;V | Describe the study design or source of data (e.g., randomized trial, cohort, or registry data), separately for the development and validation data sets, if applicable. | 20-22 |
| | 4b | D;V | Specify the key study dates, including start of accrual; end of accrual; and, if applicable, end of follow-up. | 21 |
| Participants | 5a | D;V | Specify key elements of the study setting (e.g., primary care, secondary care, general population) including number and location of centres. | 20, Supp note 1 |
| | 5b | D;V | Describe eligibility criteria for participants. | N/A |
| | 5c | D;V | Give details of treatments received, if relevant. | N/A |
| Outcome | 6a | D;V | Clearly define the outcome that is predicted by the prediction model, including how and when assessed. | SN2 |
| | 6b | D;V | Report any actions to blind assessment of the outcome to be predicted. | N/A |
| Predictors | 7a | D;V | Clearly define all predictors used in developing or validating the multivariable prediction model, including how and when they were measured. | Supp notes, 2,3 |
| | 7b | D;V | Report any actions to blind assessment of predictors for the outcome and other predictors. | N/A |
| Sample size | 8 | D;V | Explain how the study size was arrived at. | 20 |
| Missing data | 9 | D;V | Describe how missing data were handled (e.g., complete-case analysis, single imputation, multiple imputation) with details of any imputation method. | 25 |
| Statistical analysis methods | 10a | D | Describe how predictors were handled in the analyses. | 23-26 |
| | 10b | D | Specify type of model, all model-building procedures (including any predictor selection), and method for internal validation. | Table S1 |
| | 10c | V | For validation, describe how the predictions were calculated. | Table S1 |
| | 10d | D;V | Specify all measures used to assess model performance and, if relevant, to compare multiple models. | 24-27 |
| | 10e | V | Describe any model updating (e.g., recalibration) arising from the validation, if done. | Supp note 6 |
| Risk groups | 11 | D;V | Provide details on how risk groups were created, if done. | N/A |
| Development vs. validation | 12 | V | For validation, identify any differences from the development data in setting, eligibility criteria, outcome, and predictors. | Supp note 6 |
| **Results** | | | | |
| Participants | 13a | D;V | Describe the flow of participants through the study, including the number of participants with and without the outcome and, if applicable, a summary of the follow-up time. A diagram may be helpful. | N/A |
| | 13b | D;V | Describe the characteristics of the participants (basic demographics, clinical features, available predictors), including the number of participants with missing data for predictors and outcome. | Supp notes 2,3 |
| | 13c | V | For validation, show a comparison with the development data of the distribution of important variables (demographics, predictors and outcome). | x |
| Model development | 14a | D | Specify the number of participants and outcome events in each analysis. | Fig S5 |

[9] Collins GS, Reitsma JB, Altman DG, Moons KG. Transparent reporting of a multivariable prediction model for individual prognosis or diagnosis (TRIPOD): The TRIPOD statement. Downloaded from https://www.equator-network.org/reporting-guidelines/tripod-statement/

| | | | | |
|---|---|---|---|---|
| | 14b | D | If done, report the unadjusted association between each candidate predictor and outcome. | N/A |
| Model specification | 15a | D | Present the full prediction model to allow predictions for individuals (i.e., all regression coefficients, and model intercept or baseline survival at a given time point). | x |
| | 15b | D | Explain how to the use the prediction model. | Table S1 |
| Model performance | 16 | D;V | Report performance measures (with CIs) for the prediction model. | Figs 2-5, S12, S13, S15 |
| Model-updating | 17 | V | If done, report the results from any model updating (i.e., model specification, model performance). | Fig S15 |
| **Discussion** | | | | |
| Limitations | 18 | D;V | Discuss any limitations of the study (such as nonrepresentative sample, few events per predictor, missing data). | 18 |
| Interpretation | 19a | V | For validation, discuss the results with reference to performance in the development data, and any other validation data. | 16-17 |
| | 19b | D;V | Give an overall interpretation of the results, considering objectives, limitations, results from similar studies, and other relevant evidence. | 16-19 |
| Implications | 20 | D;V | Discuss the potential clinical use of the model and implications for future research. | 19 |
| **Other information** | | | | |
| Supplementary information | 21 | D;V | Provide information about the availability of supplementary resources, such as study protocol, Web calculator, and data sets. | Supp Mat |
| Funding | 22 | D;V | Give the source of funding and the role of the funders for the present study. | 27 |
